# Development and external validation of a multivariable regression model for bacteraemia in adults presenting to emergency departments

**DOI:** 10.64898/2026.07.17.26358264

**Authors:** Thomas HA Samuels, Robbie Forrest-Hammond, Chloe Stockford, Steve Harris, David W Eyre, Rishi K Gupta, Mahdad Noursadeghi

## Abstract

**Background:** Bacteraemia is associated with poor outcomes but the diagnostic gold standard, peripheral blood culture, takes up to 24 hours to become clinically actionable, hampering early management decisions in suspected infection. Single predictors and existing sepsis risk scores discriminate poorly, and few multivariable bacteraemia models have been adequately validated in UK populations.

**Methods:** We developed a logistic regression model, using backwards AIC-based selection of pre- defined candidate predictors routinely available within hours of hospital attendance, in a retrospective cohort of 33,874 hospital encounters at University College London Hospitals (UCLH) between 2019 and 2024. Continuous predictors were modelled using restricted cubic splines and missing data handled using multiple imputation. Model performance was assessed via internal-external cross-validation and prediction instability analysis, before temporal validation in held-out 2024 UCLH data and external validation in 53,669 hospital encounters from the Infections in Oxfordshire Research Database (IORD).

**Results:** Bacteraemia occurred in 5.2% of UCLH and 8.9% of IORD encounters, respectively. Twenty predictors were retained, spanning demographics, comorbidities, vital signs and blood tests. Discrimination was stable across development time periods (pooled c-statistic 0.82, 95%CI 0.81–0.84) and was maintained in temporal (0.83, 0.79–0.87) and external validation (0.83, 0.82–0.83), with excellent calibration in external validation (calibration slope 1.08 (1.05-1.11); calibration-in-the-large 0.01 (−0.02-0.04)). The model outperformed single predictors, established risk scores, and a reconstructed comparator model, and showed superior net benefit in decision curve analysis. Performance was consistent across age, sex, ethnicity and socioeconomic subgroups but degraded when blood cultures were sampled more than six hours after attendance and varied by likely infection site.

**Conclusions:** This model accurately predicts bacteraemia using routinely collected data available within hours of hospital attendance, with performance maintained in a large, independent external validation cohort. It offers a generalisable, clinically interpretable tool to support early decision-making in suspected infection, pending further work to establish optimal implementation thresholds.

## Introduction

Infectious diseases are one of the commonest causes of Emergency Department (ED) attendance in the United Kingdom (UK).^1^ The presence of bacteria in the bloodstream during infection, or bacteraemia, is associated with excess mortality, and often changes clinical management.^2^ Peripheral blood culture is the gold-standard diagnostic investigation for bacteraemia. Effective and timely blood culture testing has been shown to both improve patient outcomes^3^ and reduce hospital costs.^4^ However, despite modern equipment and techniques, it can take more than 24 hours to produce a clinically actionable positive result, and up to 5 days to give a definitive negative result.^5^ This may contribute to sub-optimal clinical decision-making in the initial management of infection, such as the decision to admit, and the choice, dose and route of antibiotics.

Bacteraemia does not occur uniformly in cases of suspected infection; rates are known to differ depending on the severity of infection,^6^ patient demographics,^7^ immune status,^8^ and the site of infection.^9^ This uneven distribution of risk raises the possibility of stratifying risk of bacteraemia early in the healthcare episode, before microbiological results are known. Predicting those patients most likely to be bacteraemic at the outset could support management decisions before results become available. In addition, it may also support targeted blood culture testing to improve diagnostic yield.

Single predictors (e.g., fever or elevated white cell count) are insufficiently discriminative for bacteraemia for this task.^9^ Commonly used sepsis risk scores, such as qSOFA, are also only weakly predictive.^10^ Multivariable predictive models, by contrast, have shown some promise. A recent systematic review identified twenty models developed between 2008 and 2023,^11^ some of good methodological quality and with reasonable discriminatory performance. However, many of these models were at high risk of bias, were developed on small sample sizes, used variables that are not universally available,^12^ or lacked external validation in other cohorts. To date, only one has been externally validated in UK data, with a very modest sample size.^13,14^

In the current study, we aimed to develop and validate a diagnostic model to predict bacteraemia in two large cohorts of adult patients with suspected infection in the UK, using only predictors routinely available within a few hours of attendance at hospital.

## Methods

### Study design, data sources and participants

For the model development cohort, we formed a retrospective observational cohort of adult patients who presented to University College London Hospitals NHS Foundation Trust (UCLH) between the 1^st^ of May 2019 and the 30^th^ of April 2024 using routinely collected electronic healthcare record (EHR) data. All patients ≥18 years old from whom at least one blood culture sample was taken within 24 hours of an unplanned attendance to ED were eligible for inclusion. Only patients who had previously indicated a preference against having their EHR data used in research via the National Health Service Data Opt-Out service^15^ were excluded.

A retrospective observational cohort formed from the Infections in Oxfordshire Research Database (IORD) was used for external validation cohort.^16^ This is a deidentified electronic database of patients attending or having diagnostic tests performed by Oxford University Hospitals NHS Foundation Trust, an organisation made up of four hospitals in Oxfordshire, UK. Microbiological data is linked to primary and secondary EHR data to match relevant demographic and clinical details. Adult individuals in the IORD database who attended participating hospitals between January 2016 and March 2025 and who fulfilled the same inclusion criteria as the development cohort were eligible to form the external validation cohort.

All analyses were conducted in R version 4.5.2 (2025-10-31) and reported in accordance with TRIPOD+AI guidance for the reporting of predictive model development; a completed TRIPOD+AI checklist is present in the supplementary material.^17^ The code used for model development and analysis is available in a publicly available GitHub repository at the following URL: [TBC].

### Ethics

This study was approved by the University College London/University College London Hospitals NHS Foundation Trust Joint Research Office and UCLH Data Trust Committee using the Data Access Process for Research (DAP-R) pathway. This approval pathway enables the use of routinely collected and anonymised electronic healthcare record data for defined research purposes without the need for individual participant consent. Individuals may opt out of having their data used under this pathway using the National Health Service data opt-out, and these participants were excluded from the current study. IORD has ethical approvals from the National Research Ethics Service South Central Oxford C Research Ethics Committee (19/SC/0403) and the National Confidentiality Advisory Group (19/CAG/0144).

### Outcomes and predictors

We sought to predict clinically significant bacteraemia, hence bacteraemia was defined a priori as the isolation of any organism considered to be pathogenic from at least one blood culture set, or the repeated isolation (≥3 blood culture sets) of an organism considered to be a contaminant, in the first 24 hours of that patient encounter. The isolation of a contaminant organism on ≤2 occasions in the encounter was considered a negative blood culture. The range of organisms considered to be pathogenic was defined broadly, to maximise model generalisability. Organisms considered to be pathogenic (versus contaminant) are listed in the supplement.

Candidate predictors for the model were predefined, based on their availability in routinely collected EHR data within 24 hours of hospital attendance, and based on prior knowledge of association with bacteraemia including use in previously published models. We only considered candidate predictors that were available in ≥60% of participants, to prioritise routinely measured variables. The first occurrence of an individual vital sign or blood test recorded in the EHR were used. Antibiotic prescription data and candidate predictors unavailable within 24 hours of hospital attendance were not considered for inclusion.

Clinical data were extracted directly from the EHR/database for both cohorts. Ethnicity was self-reported and assigned using standard NHS coding, and socioeconomic status was established by matching patient postcodes to the 2019 Index of Multiple Deprivation using the 2011 UK Local Layer Super Output Areas, where available.^18,19^ Comorbidity diagnoses were assigned using available ICD-10 codes for that patient encounter, matching diagnoses to the ICD-10 UK 5^th^ edition as defined by the UK National Clinical Coding Standards and categorised using code sets from Health Data Research UK.^20^ Site of infection was established by using the same methodology on the primary encounter ICD-10 code, but was not eligible for inclusion as a candidate predictor into the model since it reflected discharge diagnosis and was not available at the point of hospital presentation. Comorbidity and site of infection ICD-10 code sets are presented in the supplementary appendix.

### Model development and validation

We developed a logistic regression model to predict bacteraemia using backwards selection of predefined candidate predictors. This selection was based on Akaike Information Criterion (AIC), a statistical property that balances the model’s capacity to explain the variation in the underlying data, whilst penalising for each additional variable retained in the model.^21^ Candidate predictors were sequentially removed from the model and the AIC calculated; predictors in the model with lowest AIC were retained. Missing data were handled using multiple imputation (MI) using the aRegImpute package in R, to allow compatibility with restricted cubic splines for continuous variables, as previously.^22^ All predictors and the outcome were included in the MI models. Variable selection was performed in each of the 10 MI datasets. Predictors selected in >50% MI datasets were retained in the final model. All continuous predictors were modelled using restricted cubic splines with 3 knots to capture non-linear outcome associations.

We then evaluated the model with internal-external cross-validation (IECV), as previously.^22,23^ This method examines the potential generalisability of the model by examining heterogeneity in performance between, in this case, time period. The development data was split by quarter-year, and each time period was iteratively left out of the model development data and used for validation by quantifying discrimination and calibration. Discrimination measures how well a model determines between those who do and do not meet the outcome and is measured using the c-statistic (analogous to the area under a Receiver Operator Characteristic curve). Calibration measures how well model predictions match observed outcomes. The calibration slope assesses if the spread of predictions is accurate (slopes <1 suggest overfitting, whereas slopes >1 indicate predictions are too conservative), while calibration-in-the-large evaluates systematic over/under prediction of the outcome (<0 indicates predictions are too high overall, whereas >0 indicates systematic underprediction). Calibration is also visualised using calibration plots. Performance was analysed in each MI dataset, before being pooled using Rubin’s rules. We then used random effects metanalysis to estimate pooled measures of discrimination and calibration across time periods in the development data.

We also evaluated the model for prediction instability. This uses bootstrapping with replacement to evaluate a range of possible alternative models that could have been trained on a different sample from the same population, using the predictions made by those models to estimate the uncertainty (or instability) in the original model’s predictions.^24^ Briefly, the model development process described above was repeated on 1000 bootstrapped datasets derived from the original development data, generating 1000 bootstrapped models. New predictions were then calculated for each individual in the development dataset using each of these bootstrapped models. These bootstrapped predictions were plotted against the predictions made by the original model in a prediction instability plot. Exemplar patients had their bootstrapped predictions plotted as a distribution against their original predicted risk with 95% bootstrapped prediction intervals, to demonstrate model instability over an example range of predicted risks.

The final model was then trained on the full development dataset (2019-2023), before being validated temporally in held-out UCLH validation data (January 2024 onwards), and externally in the Oxford IORD dataset. Discrimination and calibration were also^17^ reported stratified by age, sex, ethnicity and socioeconomic status, to analyse model fairness.^17^ We benchmarked model performance against the single best candidate predictor, other commonly used risk scores for severe infection and sepsis, and other models for bacteraemia for which data was available in ≥60% patients and for which model reconstruction was possible from the original manuscript.^25–28^ We also conducted decision curve analysis in both the temporal and external validation datasets to quantify overall net benefit of implementing the model to inform clinical decision-making compared with test all, test none, and other benchmark approaches.^29^ Decision curve analysis calculates net benefit as the proportion of true cases identified by the model, minus the proportion of false positives, weighted by the number of unnecessary tests a clinician would accept for each true case correctly identified at a given threshold (analogous to a number-needed-to-test). This was then evaluated across a plausible range of thresholds for bacteraemia. A stratified analysis of model performance by time to blood culture sampling and by site of infection was performed in the whole UCLH and external validation cohorts, the former to ensure adequate sample size and event rates.

### Sample size

Based on the sample size of our development dataset, we calculated that 224 parameters could be considered for inclusion during model development based on a predicted c-statistic of 0.8 and an outcome prevalence of 0.05 using the pmsampsize package in R, ^30^ far in excess of the number of pre-defined candidate predictors we included.

## Results

We included 33,874 hospital encounters from 25,195 unique patients in the UCLH dataset, of which 1,760 encounters (5.2%) fulfilled the primary outcome of bacteraemia. The most common organisms isolated in these blood cultures were *Escherichia coli* (27.4%), *Staphylococcus aureus* (13.3%) and *Klebsiella* species (10.2%); others are presented in Figure S2. 31,379 hospital encounters (all encounters between the start of the study period and the end of 2023) were included in model development, and 2,495 encounters from 2024 were held out for temporal validation; bacteraemia occurred in 5.2% and 5.6% of these encounters, respectively. Participant characteristics by outcome are presented in Table 1. Participant flow diagrams are shown in the supplement Figure S1 and participant characteristics by development/validation cohort in Table S1.

**Table 1:**
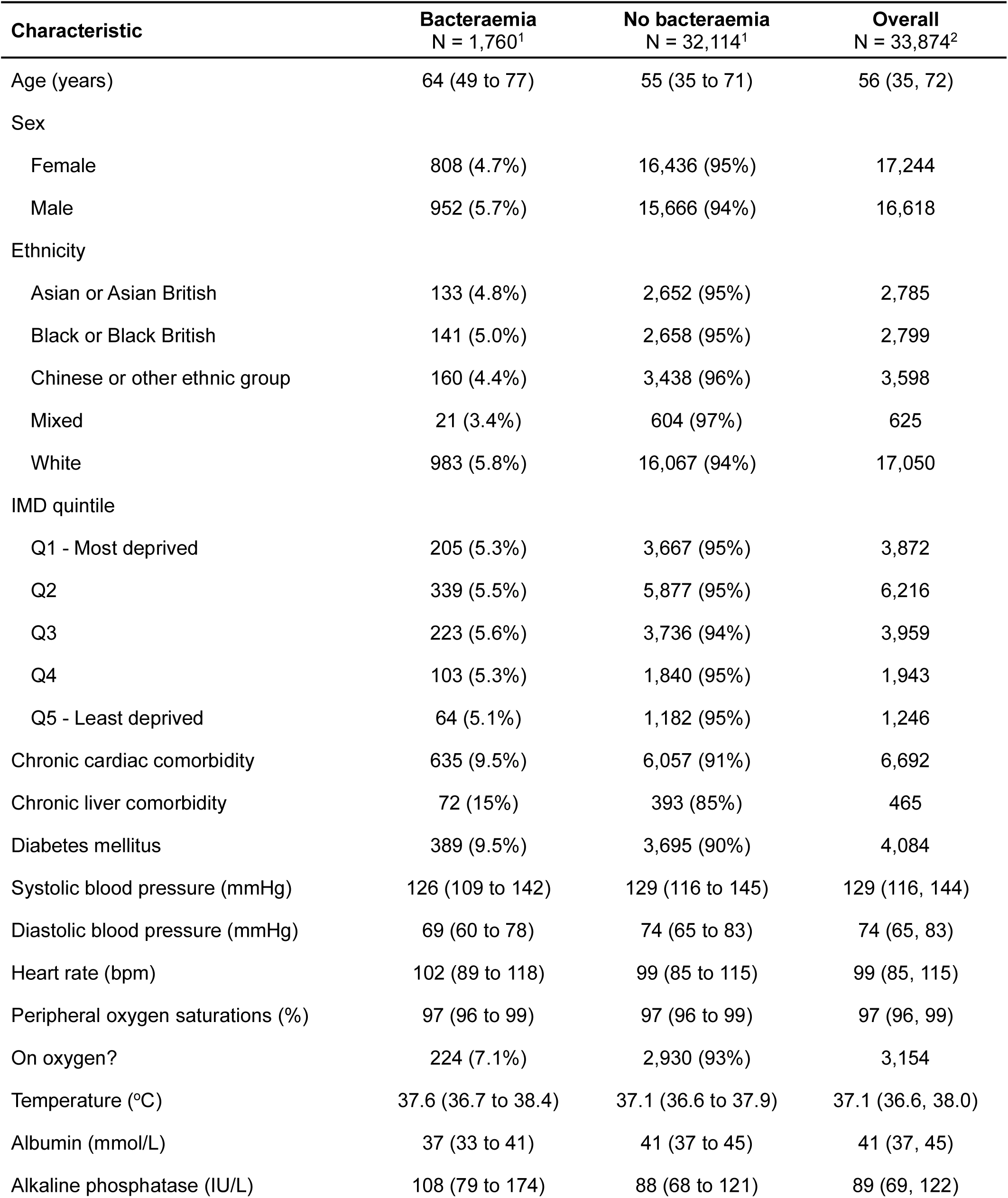
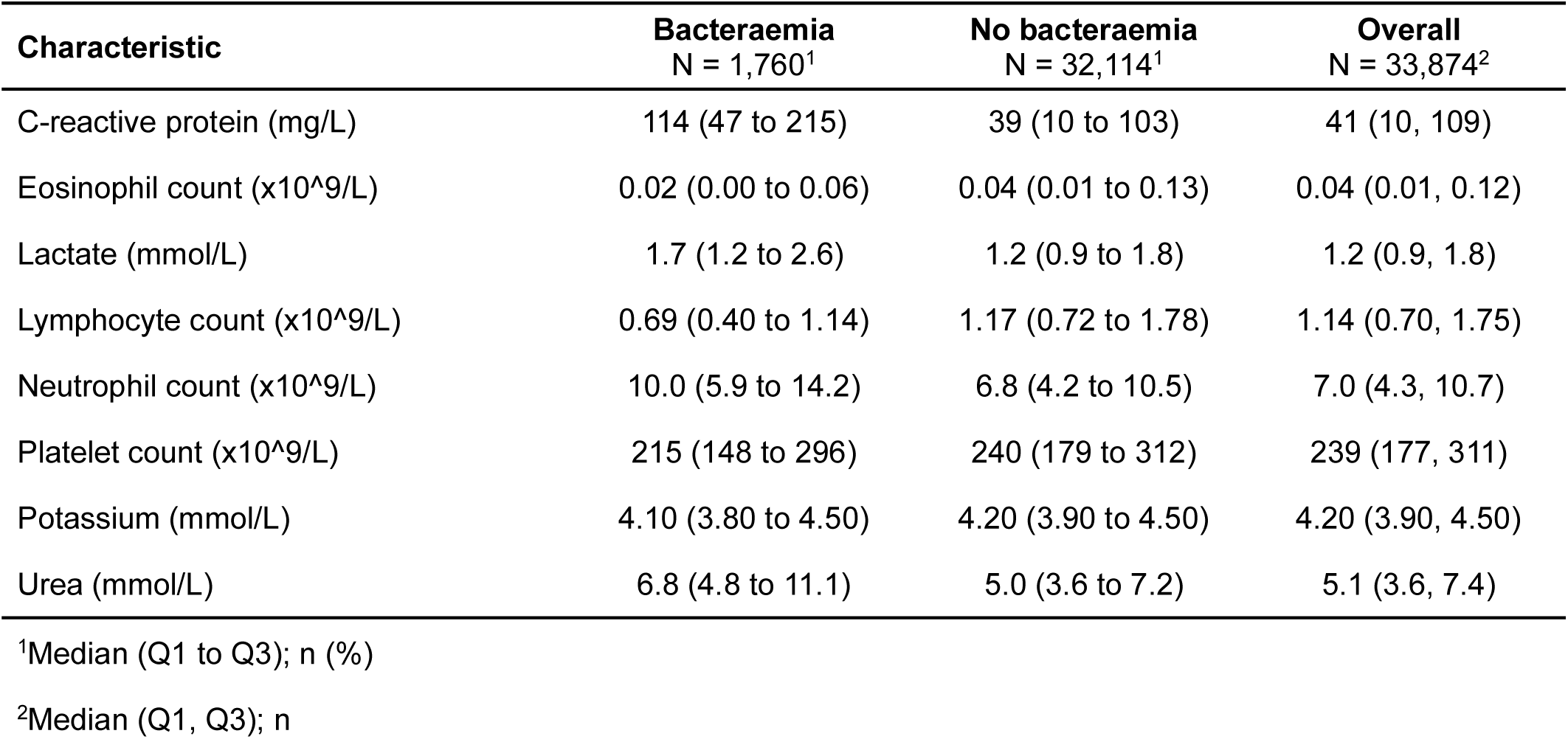
Baseline characteristics of the UCLH cohort, stratified by outcome. Continuous variables are shown as median (interquartile range) and categorical variables as n (%). Data missingness is presented in Table S2. IMD = Index of Multiple Deprivation.

| Characteristic | Bacteraemia<br>N = 1,760<sup>1</sup> | No bacteraemia<br>N = 32,114<sup>1</sup> | Overall<br>N = 33,874<sup>2</sup> |
| --- | --- | --- | --- |
| Age (years) | 64 (49 to 77) | 55 (35 to 71) | 56 (35, 72) |
| Sex |  |  |  |
| Female | 808 (4.7%) | 16,436 (95%) | 17,244 |
| Male | 952 (5.7%) | 15,666 (94%) | 16,618 |
| Ethnicity |  |  |  |
| Asian or Asian British | 133 (4.8%) | 2,652 (95%) | 2,785 |
| Black or Black British | 141 (5.0%) | 2,658 (95%) | 2,799 |
| Chinese or other ethnic group | 160 (4.4%) | 3,438 (96%) | 3,598 |
| Mixed | 21 (3.4%) | 604 (97%) | 625 |
| White | 983 (5.8%) | 16,067 (94%) | 17,050 |
| IMD quintile |  |  |  |
| Q1 - Most deprived | 205 (5.3%) | 3,667 (95%) | 3,872 |
| Q2 | 339 (5.5%) | 5,877 (95%) | 6,216 |
| Q3 | 223 (5.6%) | 3,736 (94%) | 3,959 |
| Q4 | 103 (5.3%) | 1,840 (95%) | 1,943 |
| Q5 - Least deprived | 64 (5.1%) | 1,182 (95%) | 1,246 |
| Chronic cardiac comorbidity | 635 (9.5%) | 6,057 (91%) | 6,692 |
| Chronic liver comorbidity | 72 (15%) | 393 (85%) | 465 |
| Diabetes mellitus | 389 (9.5%) | 3,695 (90%) | 4,084 |
| Systolic blood pressure (mmHg) | 126 (109 to 142) | 129 (116 to 145) | 129 (116, 144) |
| Diastolic blood pressure (mmHg) | 69 (60 to 78) | 74 (65 to 83) | 74 (65, 83) |
| Heart rate (bpm) | 102 (89 to 118) | 99 (85 to 115) | 99 (85, 115) |
| Peripheral oxygen saturations (%) | 97 (96 to 99) | 97 (96 to 99) | 97 (96, 99) |
| On oxygen? | 224 (7.1%) | 2,930 (93%) | 3,154 |
| Temperature (°C) | 37.6 (36.7 to 38.4) | 37.1 (36.6 to 37.9) | 37.1 (36.6, 38.0) |
| Albumin (mmol/L) | 37 (33 to 41) | 41 (37 to 45) | 41 (37, 45) |
| Alkaline phosphatase (IU/L) | 108 (79 to 174) | 88 (68 to 121) | 89 (69, 122) |

| **Characteristic** | **Bacteraemia**<br>**N = 1,760<sup>1</sup>** | **No bacteraemia**<br>**N = 32,114<sup>1</sup>** | **Overall**<br>**N = 33,874<sup>2</sup>** |
| --- | --- | --- | --- |
| C-reactive protein (mg/L) | 114 (47 to 215) | 39 (10 to 103) | 41 (10, 109) |
| Eosinophil count (x10<sup>9</sup>/L) | 0.02 (0.00 to 0.06) | 0.04 (0.01 to 0.13) | 0.04 (0.01, 0.12) |
| Lactate (mmol/L) | 1.7 (1.2 to 2.6) | 1.2 (0.9 to 1.8) | 1.2 (0.9, 1.8) |
| Lymphocyte count (x10<sup>9</sup>/L) | 0.69 (0.40 to 1.14) | 1.17 (0.72 to 1.78) | 1.14 (0.70, 1.75) |
| Neutrophil count (x10<sup>9</sup>/L) | 10.0 (5.9 to 14.2) | 6.8 (4.2 to 10.5) | 7.0 (4.3, 10.7) |
| Platelet count (x10<sup>9</sup>/L) | 215 (148 to 296) | 240 (179 to 312) | 239 (177, 311) |
| Potassium (mmol/L) | 4.10 (3.80 to 4.50) | 4.20 (3.90 to 4.50) | 4.20 (3.90, 4.50) |
| Urea (mmol/L) | 6.8 (4.8 to 11.1) | 5.0 (3.6 to 7.2) | 5.1 (3.6, 7.4) |

<sup>1</sup>Median (Q1 to Q3); n (%)
<sup>2</sup>Median (Q1, Q3); n

Twenty predictors were retained in ≥50% of MI datasets during model development (Table S7). These included one demographic predictor (age); three comorbidity predictors (chronic liver disease, diabetes mellitus, chronic cardiac comorbidity); six vital sign predictors (systolic blood pressure, diastolic blood pressure, heart rate, temperature, peripheral oxygen saturations and whether these were taken on supplemental oxygen); four blood haematological predictors (neutrophil, lymphocyte, eosinophil and platelet counts) and six blood biochemical predictors (albumin, alkaline phosphatase, c-reactive protein, lactate, potassium and urea). Predictor-outcome associations are presented in Figure 1. Of the vital sign and blood predictors, 94.4% were recorded within 6 hours of attendance to hospital when present (median 0.8 hours, IQR 0.3 to 1.6; Table S8, Figure S3). Model coefficients are shown in Table S6.

**Figure 1:**
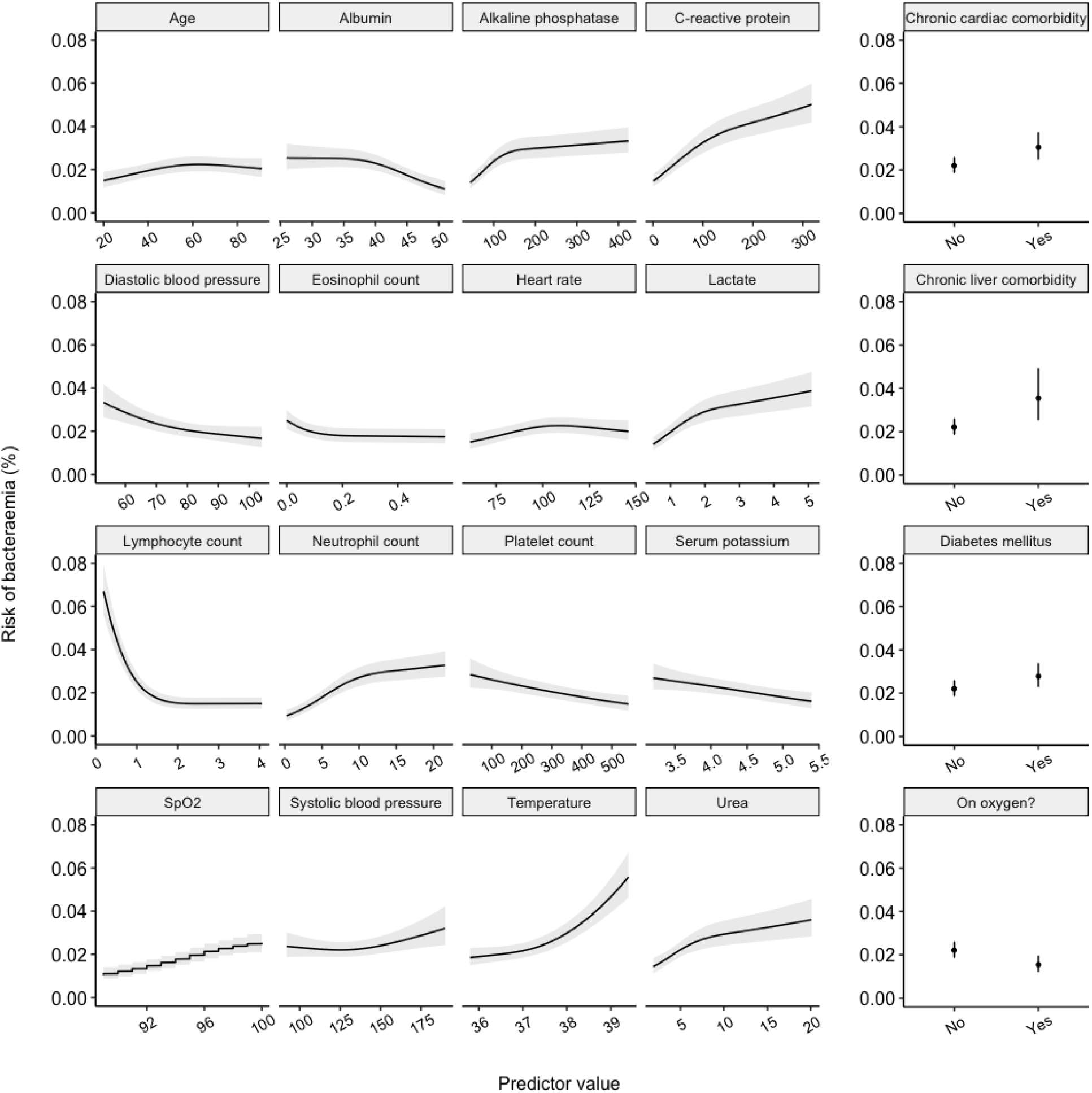
Multivariable associations between selected predictors and bacteraemia in the development cohort. Continuous variables were modeled using restricted cubic splines. The final model parameters are pooled across multiply imputed datasets (total sample size for model development = 31379 participants in the UCLH cohort). For continuous variables, black lines represent point estimates and grey shaded regions represent 95% confidence intervals. For categorical variables, black dots represent point estimates and black lines represent 95% confidence intervals. To plot risk for each variable, all other variables were held at their median (continuous) or mode (categorical) value. 95% confidence intervals were calculated from the regression model.

Model discrimination in the development data was consistent over time (pooled c-statistic 0.82 [95%CI 0.81 to 0.84]; Table 2 and Figure S4). Calibration, in the form of slope and calibration-in-the-large (CITL), was also relatively consistent (pooled slope 0.98 [95%CI 0.93 to 1.03]; pooled CITL 0.00 [95%CI −0.08 to 0.09], although variation in CITL suggested some underprediction of risk during winter periods and during the initial waves of the COVID-19 pandemic in the UK. The model showed good overall prediction stability, with a mean absolute prediction error of 0.9% (IQR 0.1% to 0.9%; Figure S6a). Predictions were most stable at low predicted bacteraemia risks (at predicted risk of 2%, 95% prediction interval 1.7% to 2.6%) but more variable at higher predicted risks (at predicted risk of 10%, 95% prediction interval 7.1% to 16.4%), where data were sparser (Figure S6b).

**Table 2:** Model performance. The table shows the model performance by cohort: pooled metrics (by random effects metanalysis) in UCLH development data during internal-external cross-validation by time-period (IECV REMA); in held-out temporal validation in UCLH; and in external validation in the IORD Oxford dataset. Brackets contain 95% confidence intervals. CITL = calibration-in-the-large.

| Cohort | C-statistic | Calibration slope | CITL | Observed events | Expected events | Sample size |
| --- | --- | --- | --- | --- | --- | --- |
| UCLH development (IECV REMA pooled) | 0.82 (0.81 - 0.84) | 0.98 (0.93 - 1.03) | 0 (-0.08 - 0.09) | 1621 | 1621 | 31379 |
| UCLH held-out temporal validation | 0.83 (0.79 - 0.87) | 1.05 (0.89 - 1.21) | 0.03 (-0.15 - 0.21) | 139 | 135 | 2495 |
| Oxford IORD external validation | 0.83 (0.82 - 0.83) | 1.08 (1.05 - 1.11) | 0.01 (-0.02 - 0.04) | 4765 | 4736 | 53669 |

In the held out temporal validation cohort, the median predicted risk in bacteraemic patients was 12.5% (IQR 6.8% to 25.6%), and in non-bacteraemic patients 2.4% (IQR 1.1% to 5.5%; Figure 2A). Discrimination performance was similar to internal-external cross-validation (c-statistic 0.83, 95%CI 0.79 to 0.87; Table 2). In calibration predicted risk closely tracked observed risk (Figure 2B), although confidence intervals widened significantly at higher risks where data were sparse.

**Figure 2:**
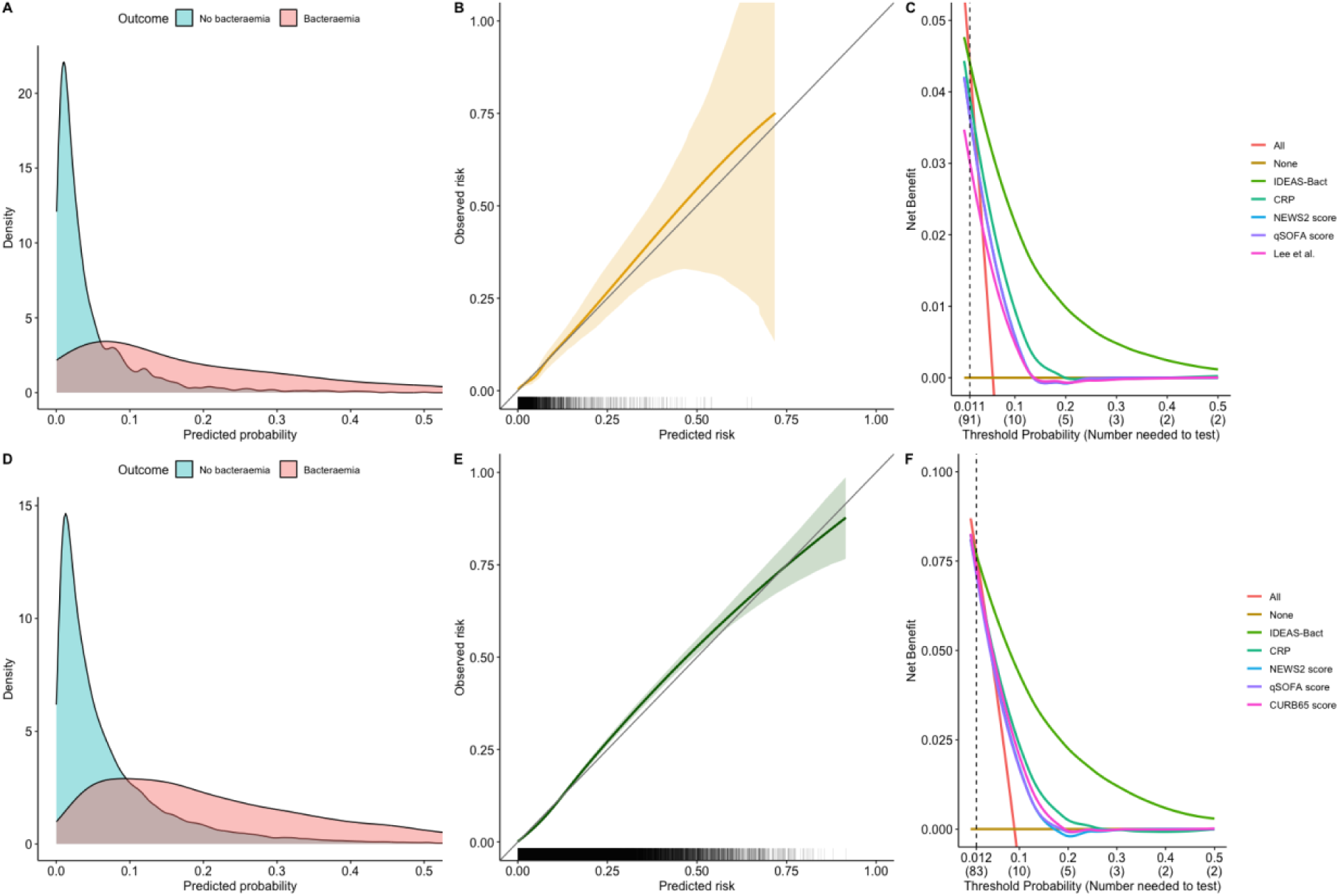
Held-out and External validation. Panels (A–C) show performance of the IDEAS-Bacteraemia model in the held-out temporal validation cohort at UCLH. Panels (D–F) show performance in the Oxford IORD external validation cohort. Panel (A) shows a density plot of the distribution of predicted model risk, stratified by outcome. Panel (B) shows calibration using a loess smoother; shaded areas represent 95% confidence intervals generated by fitting a loess smoother to 500 bootstrapped samples, and rug plots on the x-axis show the distribution of predicted risk. Panel (C) shows decision curve analysis quantifying net benefit versus alternative approaches: the single best univariate predictor (C-reactive protein), commonly used risk scores (NEWS2, qSOFA), other predictive models that could be reconstructed in the data (Lee et al.), test-all (All), and test-none (None). Panels (D–F) show equivalent plots for the Oxford IORD cohort. Vertical dashed lines in Panels (C) and (F) represent the number needed to test above which test-all becomes a superior strategy to the IDEAS-Bacteraemia model.

The IORD external validation dataset included 53,669 encounters from 39,332 unique patients. Participant characteristics are shown in Table S14. The distribution of model predictions was higher than in the UCLH dataset (Figure 2D; Table S15). This was driven by more extreme predictor values and was reflected in a higher cohort bacteraemia rate of 8.9%. Overall, model performance was in keeping with both the development and temporal validation cohorts, with a c-statistic of 0.83 (95%CI 0.82 to 0.83), calibration slope of 1.08 (1.05 to 1.11) and CITL 0.01 (−0.02 to 0.04). Calibration was excellent across the range of predicted risk (Figure 2E). Decision curve analysis showed similar results to the temporal validation cohort (Figure 2F).

Model discrimination varied by age and sex in the temporal validation cohort, trending towards inferiority in males (c-statistic 0.80 (95%CI 0.74 to 0.85) vs 0.86 (95%CI 0.81 to 0.91) for females, difference 0.06 (95%CI −0.14 to 0.01), p=0.09) and in the bottom quartile of age (18-36 years old, c-statistic 0.72 (95%CI 0.58 to 0.85); c-statistic 0.81 to 0.87 for other age groups, difference vs all others 0.12 (95%CI −0.26 to 0.02), p=0.1), although as overall event numbers were low, confidence intervals were wide. There was no evidence that performance varied by socioeconomic status or ethnicity (Table S9). There was no evidence of a difference in performance by age, sex, ethnicity or socioeconomic deprivation quintile in the larger external validation cohort (Table S17).

Model performance varied by blood culture sample timing and infection site. Delayed blood culture sampling, more than 6 hours from the time of hospital attendance, led to degraded model discrimination and calibration when analysed in the whole UCLH cohort (Figure 3A), with optimal performance seen when blood cultures were taken within 6 hours of hospital attendance. These findings were replicated in the external validation cohort (Figure 3B). Discrimination differed by site of infection but differences in CITL were more marked. The model systematically underpredicted risk for urinary, hepatobiliary and neurological infections (CITL point estimate 0.41, 0.34 and 0.65, respectively), whilst systematically overpredicting risk for lower respiratory, upper respiratory and skin and soft tissue infections (CITL point estimate −1.28, −1.01 and −0.62, respectively); this likely represents the differential risk of bacteraemia from infections at these sites, which the model does not capture in its predictions. We could not analyse performance by infection site in the external validation cohort.

**Figure 3:**
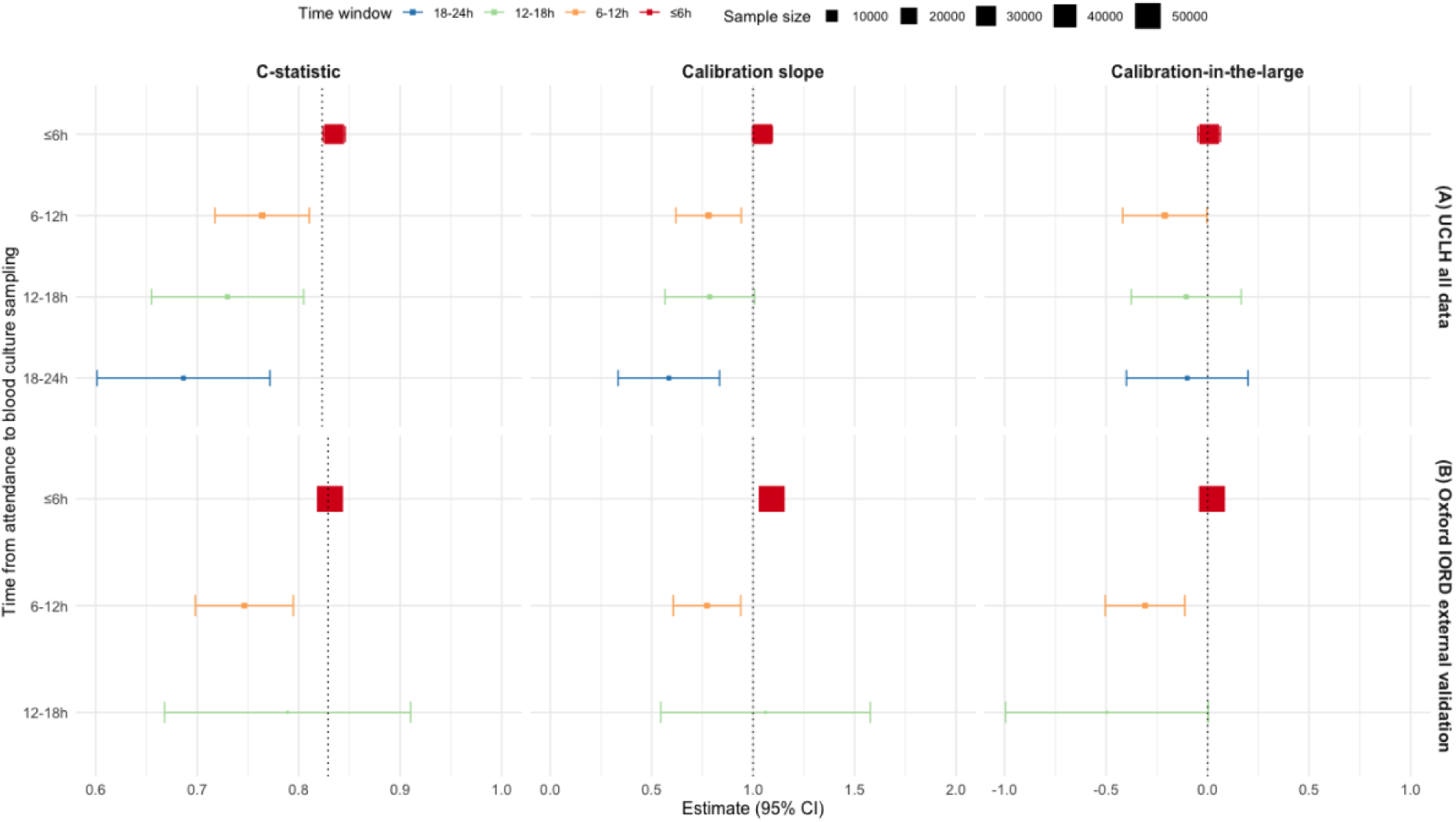
Model performance by time from hospital attendance to blood culture sampling. Panel (A) shows performance for the model in all UCLH data. Panel (B) shows performance in external validation in Oxford IORD data. Time from attendance to blood culture sampling is categorised into 6-hour windows; 18-24 is not shown for the external validation data due to small sample size. Squares show point estimates and are sized proportional to group size. Bars show 95% confidence intervals. Vertical dashed grey reference lines indicate overall model performance for c-statistic, and perfect calibration for the calibration slope and calibration-in-the-large.

**Figure 4:**
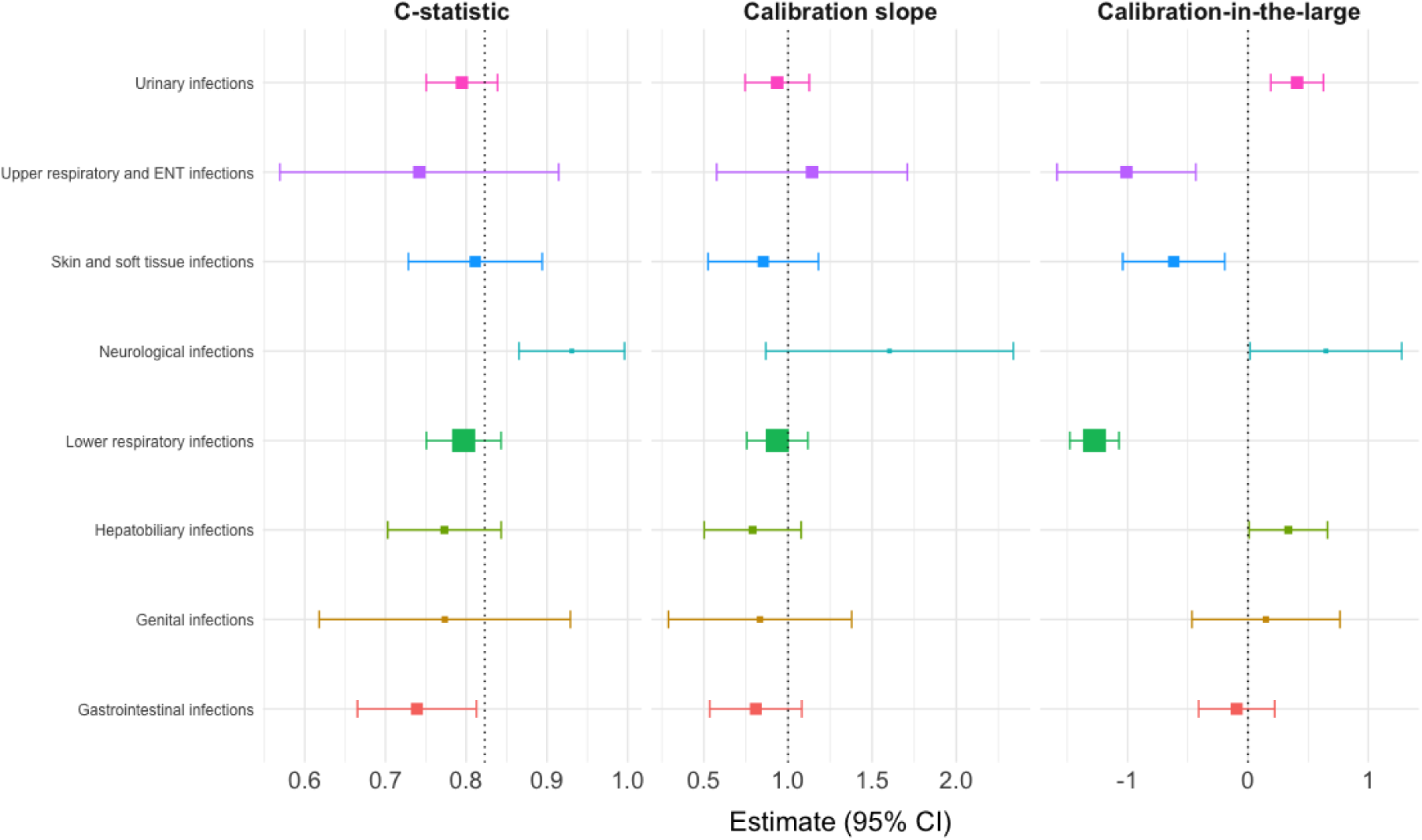
Model performance by infection source in UCLH data. Infection source is shown on the y axis and was assigned using primary discharge ICD-10 codes. Squares show point estimates and are sized proportional to group size. Bars show 95% confidence intervals. Vertical dashed grey reference lines indicate overall model performance for c-statistic, and perfect calibration for the calibration slope and calibration-in-the-large.

Of the model variables, C-reactive protein was the strongest univariate predictor (c-statistic 0.70, 95%CI 0.65 to 0.74; Table S10) but its discrimination was inferior to the model itself. This was also seen with commonly used risk scores (Table S11). Of the identified pre-existing models for bacteraemia, only the model authored by Lee et al. could be reconstructed in the available data.^28^ Discrimination in the temporal validation dataset, calculated using their point score, was inferior to our model for both respiratory infections (for which the original model was trained), and in the overall cohort (Table S13). To report calibration, the model intercept was calibrated to our validation data as no intercept was reported in the original paper. Calibration assessment suggested predictions were too extreme, with a slope of 0.67 (95%CI 0.03 to 1.31).

In decision curve analysis, shown in Figure 2C, our model showed superior net benefit to alternative strategies over a wide range of number-needed-to-test (NNT), being inferior only to a test all strategy at a NNT of 91 (threshold probability of 0.011). When the validation dataset was restricted to respiratory patients only, the model proved superior to Lee et al.’s model over a NNT of 11 (Figure S7).

## Discussion

With sufficient sample volume and when taken at an early stage, before systemic antibiotics, blood cultures remain the gold standard for the detection of bacteraemia. However, even the most advanced culture systems can take up to 24 hours’ incubation to produce a positive result.^5^ We present a logistic regression model that accurately predicts risk of bacteraemia in adult patients with suspected infection within hours of their presentation to hospital. Alongside our collaborator’s reciprocal work,^31^ it has been developed and validated on the largest combined cohort of patients yet reported in the literature. The model accurately predicted bacteraemia during external validation, demonstrating excellent calibration and surpassing the net benefit of alternative approaches that we were able to replicate in our data during decision curve analysis. These results suggest significant potential clinical utility and generalisability across UK and similar populations internationally.

The model produces an individualised probability output that the patient under assessment currently has a bacteraemia. These predictions could help inform clinical decision-making in patients with suspected infection prior to their culture results becoming available, supporting decisions on admission vs discharge, selection of antibiotic class and route of administration, and seeking further advice from infection specialists, including the potential for targeting pathogen metagenomic testing to those at high risk. Designed using a frequentist regression approach, the model is transparent and interpretable, enabling implementation and portability between institutions. It will also facilitate any ongoing prospective assessment of its performance, such as recalibration in response to calibration drift over time.^32^ The model could be embedded directly into EHR systems, enabling automated predictions to be made in real time, which in turn could be presented as a decision-assist tool to clinicians using pop-ups or prompts.

There were plausible predictor-outcome associations across included model variables, many of which are already known to be associated with severe infection (which is, in turn, associated with bacteraemia).^9^ Some, including rising age, the presence of diabetes mellitus and chronic liver disease are known to contribute to both the risk of infection and infection severity.^33–35^ Other predictors are known to correlate with severe infection or mortality, such as rising lactate, C-reactive protein, and urea, and falling lymphocyte count.^11,36,37^ By contrast, the association of lower peripheral oxygen saturation and supplemental oxygen requirement with lower bacteraemia risk initially seems counterintuitive. However, in the absence of a model predictor capturing the source of infection, these variables may be acting as a proxy for the presence of lower respiratory tract infection, which is known to have a lower bacteraemia risk than infection at other sites.^9^

Model performance was strong across key demographic subgroups in temporal and external validation, including age, sex, socioeconomic status and ethnicity. Although discrimination in the youngest age bracket appeared weaker in temporal validation, this pattern was not seen in external validation. A similar but less pronounced phenomenon was observed in some other subgroups, e.g. sex. This likely represents the impact of small subgroup sample sizes in the temporal validation cohort. Overall, the model’s performance in the external validation subgroup analysis suggests it is likely to generate fair predictions across the target adult population.

However, performance did vary significantly according to the timing of blood culture sampling. Discrimination and calibration degraded significantly in both validation cohorts if the blood culture sampling occurred more than six hours after presentation to hospital. There are several plausible explanations for this. Firstly, blood culture sampling is known to be highly insensitive if conducted following the initiation of antibiotics, as bacterial numbers rapidly fall beneath the limit of culture detection in response to therapy.^38^ In many emergency presentations with suspected infection, antibiotics are started within hours as part of a package of interventions designed to reduce mortality from sepsis.^39^ Delayed blood culture sampling may therefore lead to an apparently negative result, even if bacteraemia might initially have been present. Secondly, our model was trained on predictor variables that were measured within hours of hospital attendance, reflecting the clinical state of the patient at that time point. As the clinical status of the patient subsequently evolves (including the potential impact of therapeutic interventions), initial clinical measurements become less reflective of the patient’s contemporaneous clinical status. Our model is best utilised to predict risk of bacteraemia in a contemporary sample, where its predictive performance is strongest.

Performance also varied considerably according to the likely source of infection. Although this was evident for discrimination, it was most stark for calibration-in-the-large. CITL in effect measures whether the model systematically over- or under-predicts risk. We observed that for sources that typically result in bacteraemia only rarely, such a lower respiratory infection or skin and soft tissue infection,^9^ CITL was strongly negative, indicating the model was systematically over-predicting risk in these patients. For sources known to associate more frequently with bacteraemia, such as urinary, hepatobiliary and neurological infections,^9^ CITL was strongly positive, suggesting systematic under-prediction of risk. These findings suggest source of infection is a key determinant of bacteraemia risk, and one which is not captured in our model by design. To an extent, this is unavoidable; in many cases the definitive source of infection is not known at the point of hospital attendance and only becomes apparent later in the patient’s admission after further investigation.^40^ However, future work could consider extensions that capture the working diagnosis at the initial patient assessment to potentially address this.

Strengths of our study include using best practice model development techniques following TRIPOD+AI guidance, using multiple imputation to handle missing data, and retaining continuous predictors modelled with restricted cubic splines to avoid loss of information from dichotomisation and model non-linear associations. We also followed best practice sample size guidance along with pre-defined candidate predictor selection and a large model development dataset to minimise the risk of model overfitting, which is reflected in its well-calibrated performance in external validation. The model demonstrated strong performance in a large independent external validation dataset, supporting its potential generalisability and clinical utility in other populations.

Our study also has limitations. The nature of our bacteraemia definition, whilst pragmatic, is not infallible. However, given the way we defined pathogens and contaminants, we are more likely to have incorrectly defined pathogens as contaminants than vice versa, which would bias our results towards the null and is unlikely to have inflated our reported model performance. Although comorbidity diagnoses were recorded in retrospect (post-discharge), they were highly likely to have been known at the point of admission. Whilst we have shown our model predictions are discriminative and well-calibrated, it is not yet clear what threshold of risk should prompt specific clinical actions in response, nor whether these predictions lead to improved patient outcomes. Further trial development work is required to prepare for a future clinical trial establishing this evidence base. This implementation gap remains a key issue in wider prediction modelling research, with very few developed models translated into interventions with meaningful clinical benefit.^41^ Furthermore, the suspected source of infection at the point of admission is not commonly or accurately captured in structured EHR data, and so could not reliably be used as a predictor in model development. The advent of large language models makes large scale extraction and summarisation of unstructured EHR data from clinical notes a possible avenue to address this shortcoming. Future work could focus on incorporating suspected source in model predictions using this method. In the meantime, clinicians using our model should be aware of the impact infection source can have on bacteraemia risk and, potentially, model calibration. This is most apparent for lower respiratory tract infections, for which the model is likely to significantly overpredict risk; we caution against use in this group.

In summary, we developed a diagnostic predictive model for bacteraemia in adults with suspected infection using routinely collected and commonly available predictors. It showed consistently strong performance over time in our centre, and excellent performance in a large external validation cohort. Future work should focus on generating evidence for optimal implementation and demonstration of impact in clinical practice.

## Data Availability

All data used in the present study are not shareable

## Acknowledgements

This study was supported by the SAFEHR team with the National Institute for Health and Care Research University College London Hospitals Biomedical Research Centre

This work uses data provided by patients and collected by the UK’s National Health Service as part of their care and support. We thank all the people of Oxfordshire who contribute to the Infections in Oxfordshire Research Database.

Research Database Team: L Butcher, H Boseley, C Crichton, DW Crook, J Davies, D Eyre, R Harrington, G Hayward, K Jeffery, F Kemp, E Morris, TEA Peto, D Prieto-Alhambra, TP Quan, R Shackell, B Shine, AS Walker, K Woods. Patient and Public Panel: M Ahmed, G Blower, J Hopkins, R Mandunya, S Markham.

## Bibliography

1. Hospital Accident & Emergency Activity, 2022-23 - GOV.UK [Internet]. [cited 2026 Jan 22]. Available from: https://www.gov.uk/government/statistics/hospital-accident-emergency-activity-2022-23

2. Verway M et al. Prevalence and Mortality Associated with Bloodstream Organisms: a Population-Wide Retrospective Cohort Study. J Clin Microbiol [Internet]. 2022 Apr 1 [cited 2025 Nov 2];60(4). Available from: /doi/pdf/10.1128/jcm.02429-21?download=true

3. Huang AM et al. Impact of rapid organism identification via matrix-assisted laser desorption/ionization time-of-flight combined with antimicrobial stewardship team intervention in adult patients with bacteremia and candidemia. Clin Infect Dis. 2013 Nov 1;57(9):1237–45.

4. Perez KK et al. Integrating rapid pathogen identification and antimicrobial stewardship significantly decreases hospital costs. Arch Pathol Lab Med. 2013;137(9):1247–54.

5. Lo CKF et al. Assessing impact of MALDI mass spectroscopy on reducing directed antibiotic coverage time for Gram-negative organisms. PLoS One [Internet]. 2020 [cited 2026 Feb 2];15(2):e0228935. Available from: https://pmc.ncbi.nlm.nih.gov/articles/PMC7043764/

6. Komori A et al. Characteristics and outcomes of bacteremia among ICU-admitted patients with severe sepsis. Scientific Reports 2020 10:1. 2020 Feb 19;10(1):1–8.

7. Fukui S et al. Bacteraemia predictive factors among general medical inpatients: a retrospective cross-sectional survey in a Japanese university hospital. BMJ Open. 2016 Jul 1;6(7):e010527.

8. Peri AM et al. Bloodstream infections in neutropenic and non-neutropenic patients with haematological malignancies: epidemiological trends and clinical outcomes in Queensland, Australia over the last 20 years. Clin Exp Med. 2023 Dec 1;23(8):4563.

9. Coburn B et al. Does this adult patient with suspected bacteremia require blood cultures? JAMA. 2012;308(5):502–11.

10. Cheng MP et al. qSOFA does not predict bacteremia in patients with severe manifestations of sepsis. Journal of the Association of Medical Microbiology and Infectious Disease Canada [Internet]. 2022 Dec 1 [cited 2026 Jan 23];7(4):364. Available from: https://pmc.ncbi.nlm.nih.gov/articles/PMC10312224/

11. Julián-Jiménez A et al. Models to predict bacteremia in the emergency department: a systematic review. Emergencias [Internet]. 2024 Jan 1 [cited 2026 Jan 22];36(1):48–62. Available from: https://pubmed.ncbi.nlm.nih.gov/38318742/

12. Julián-Jiménez A et al. Predicting bacteremia in patients attended for infections in an emergency department: the 5MPB-Toledo model - PubMed. Emergencias [Internet]. 2020 [cited 2026 Jan 22];32(2):81–9. Available from: https://pubmed.ncbi.nlm.nih.gov/32125106/

13. Hodgson LE et al. An external validation study of a clinical prediction rule for medical patients with suspected bacteraemia. Emergency Medicine Journal [Internet]. 2016 Feb 1 [cited 2026 Jan 22];33(2):124–9. Available from: https://emj.bmj.com/content/33/2/124

14. Shapiro NI et al. Who Needs a Blood Culture? A Prospectively Derived and Validated Prediction Rule. Journal of Emergency Medicine [Internet]. 2008 Oct [cited 2026 Jan 22];35(3):255–64. Available from: https://pubmed.ncbi.nlm.nih.gov/18486413/

15. National Data Opt-Out - NHS England Digital [Internet]. [cited 2026 Jun 21]. Available from: https://digital.nhs.uk/services/national-data-opt-out

16. IORD | NIHR Biomedical Research Centre: Oxford [Internet]. [cited 2026 Jun 21]. Available from: https://oxfordbrc.nihr.ac.uk/research-themes/modernising-medical-microbiology-and-big-infection-diagnostics/iord-about/

17. Collins GS et al. TRIPOD+AI statement: updated guidance for reporting clinical prediction models that use regression or machine learning methods. BMJ [Internet]. 2024 Apr 16 [cited 2024 May 7];385:q902. Available from: https://www.bmj.com/content/385/bmj-2023-078378

18. Postcode to OA (2011) to LSOA to MSOA to LAD with 2011 Classifications (May 2022) Best Fit Lookup in the UK | Open Geography Portal [Internet]. [cited 2026 May 7]. Available from: https://geoportal.statistics.gov.uk/datasets/b3c89316bba2410aa882ab073bedb523/about

19. English indices of deprivation 2019 - GOV.UK [Internet]. [cited 2026 May 7]. Available from: https://www.gov.uk/government/statistics/english-indices-of-deprivation-2019

20. Home - HDR UK [Internet]. [cited 2026 Jun 21]. Available from: https://www.hdruk.ac.uk/

21. Akaike H. A New Look at the Statistical Model Identification. IEEE Trans Automat Contr [Internet]. 1974 [cited 2026 Jan 22];19(6):716–23. Available from: https://ieeexplore.ieee.org/document/1100705

22. Samuels THA et al. Personalised risk-prediction tools for cryptococcal meningitis mortality to guide treatment stratification in sub-Saharan Africa: a prognostic modelling study based on pooled analysis of two randomised controlled trials. Lancet Glob Health [Internet]. 2025 May 1 [cited 2025 Jun 11];13(5):e920–30. Available from: https://www.thelancet.com/action/showFullText?pii=S2214109X25000105

23. Steyerberg EW et al. Prediction models need appropriate internal, internal-external, and external validation HHS Public Access. J Clin Epidemiol. 2016;69:245–7.

24. Riley RD et al. Stability of clinical prediction models developed using statistical or machine learning methods. Biometrical Journal. 2023 Dec 1;65(8).

25. Lim WS et al. Defining community acquired pneumonia severity on presentation to hospital: an international derivation and validation study. Thorax [Internet]. 2003 May 1 [cited 2024 Mar 28];58(5):377. Available from:/pmc/articles/PMC1746657/?report=abstract

26. Royal College of Physicians. National Early Warning Score (NEWS) 2: Standardising the assessment of acute-illness severity in the NHS. Vol. 17, Updated report of a working party. 2017.

27. Seymour CW et al. Assessment of Clinical Criteria for Sepsis: For the Third International Consensus Definitions for Sepsis and Septic Shock (Sepsis-3). JAMA [Internet]. 2016 Feb 23 [cited 2024 Mar 28];315(8):762–74. Available from: https://jamanetwork.com/journals/jama/fullarticle/2492875

28. Lee J et al. Bacteremia prediction model using a common clinical test in patients with community-acquired pneumonia. Am J Emerg Med [Internet]. 2014 Jul 1 [cited 2026 Jan 22];32(7):700–4. Available from: https://www-sciencedirect-com.libproxy.ucl.ac.uk/science/article/pii/S073567571400254X

29. Vickers AJ et al. A simple, step-by-step guide to interpreting decision curve analysis. Diagn Progn Res [Internet]. 2019;3(1). Available from: 10.1186/s41512-019-0064-7

30. Riley RD et al. Calculating the sample size required for developing a clinical prediction model. The BMJ. 2020 Mar 18;368.

31. Forrest-Hammond RW et al. Machine learning models to improve targeting of blood culture testing. medRxiv [Internet]. 2026 Jul 20 [cited 2026 Jul 21];2026.07.17.26358320. Available from: https://www.medrxiv.org/content/10.64898/2026.07.17.26358320v1

32. Riley RD et al. Evaluation of clinical prediction models (part 2): how to undertake an external validation study. BMJ [Internet]. 2024 Jan 15 [cited 2026 Jun 21];384. Available from: https://www.bmj.com/content/384/bmj-2023-074820

33. Albillos A et al. Cirrhosis-associated immune dysfunction. Nature Reviews Gastroenterology & Hepatology 2021 19:2 [Internet]. 2021 Oct 26 [cited 2026 Jun 21];19(2):112–34. Available from: https://www.nature.com/articles/s41575-021-00520-7

34. Liu Z et al. Immunosenescence: molecular mechanisms and diseases. Signal Transduction and Targeted Therapy 2023 8:1 [Internet]. 2023 May 13 [cited 2026 Jun 21];8(1):200-. Available from: https://www.nature.com/articles/s41392-023-01451-2

35. Hansen MLU et al. Diabetes increases the risk of disease and death due to Staphylococcus aureus bacteremia. A matched case-control and cohort study. Infect Dis [Internet]. 2017 Sep 2 [cited 2026 Jun 21];49(9):689–97. Available from: https://scholar.google.com/scholar_url?url=https://www.tandfonline.com/doi/pdf/10.1080/23744235.2017.1331463%3Fcasa_token%3DK-Bz5TTpLlUAAAAA:kQsN2DJFfziJhCiv89_VUc3AJ4ybduPLvpxLMPbP0qiPXREa4zBAaepr2fMwh-7rD_Qt5vi0v90tbw&hl=fr&sa=T&oi=ucasa&ct=ucasa&ei=RPI3aonDE5LwieoP_se1-A4&scisig=ANDmEU4LGhlrHJksLrcgq8K0WG79

36. de Jager CPC et al. Lymphocytopenia and neutrophil-lymphocyte count ratio predict bacteremia better than conventional infection markers in an emergency care unit. Critical Care 2010 14:5 [Internet]. 2010 Oct 29 [cited 2026 Jun 21];14(5):R192-. Available from: https://link.springer.com/article/10.1186/cc9309

37. Mikkelsen ME et al. Serum lactate is associated with mortality in severe sepsis independent of organ failure and shock. Crit Care Med. 2009;37(5):1670–7.

38. Cheng MP et al. Blood Culture Results Before and After Antimicrobial Administration in Patients With Severe Manifestations of Sepsis. 10.7326/M19-1696 [Internet]. 2019 Sep 17 [cited 2026 Jun 21];171(8):547–54. Available from: /doi/pdf/10.7326/M19-1696?download=true

39. Daniels R et al. The sepsis six and the severe sepsis resuscitation bundle: a prospective observational cohort study. Emergency Medicine Journal [Internet]. 2011 Jun 1 [cited 2026 Jun 21];28(6):507–12. Available from: https://emj.bmj.com/content/28/6/507

40. Dregmans E et al. Analysis of Variation Between Diagnosis at Admission vs Discharge and Clinical Outcomes Among Adults With Possible Bacteremia. JAMA Netw Open [Internet]. 2022 Jun 1 [cited 2026 Jun 21];5(6):e2218172–e2218172. Available from: https://jamanetwork.com/journals/jamanetworkopen/fullarticle/2793511

41. Meehan AJ et al. Clinical prediction models in psychiatry: a systematic review of two decades of progress and challenges. Molecular Psychiatry 2022 27:6 [Internet]. 2022 Apr 1 [cited 2026 Jun 21];27(6):2700–8. Available from: https://www.nature.com/articles/s41380-022-01528-4

