## Supplementary Appendix for "Development and external validation of a multivariable regression model for bacteraemia in adults presenting to emergency departments"

#### S1: Study population

###### Supplementary Figure S1: Study participant flow diagram

Panel (A) shows the study participant flow diagram for the UCLH development and held-out temporal validation cohort. Panel (B) shows the study participant flow diagram for the Oxford IORD external validation cohort.

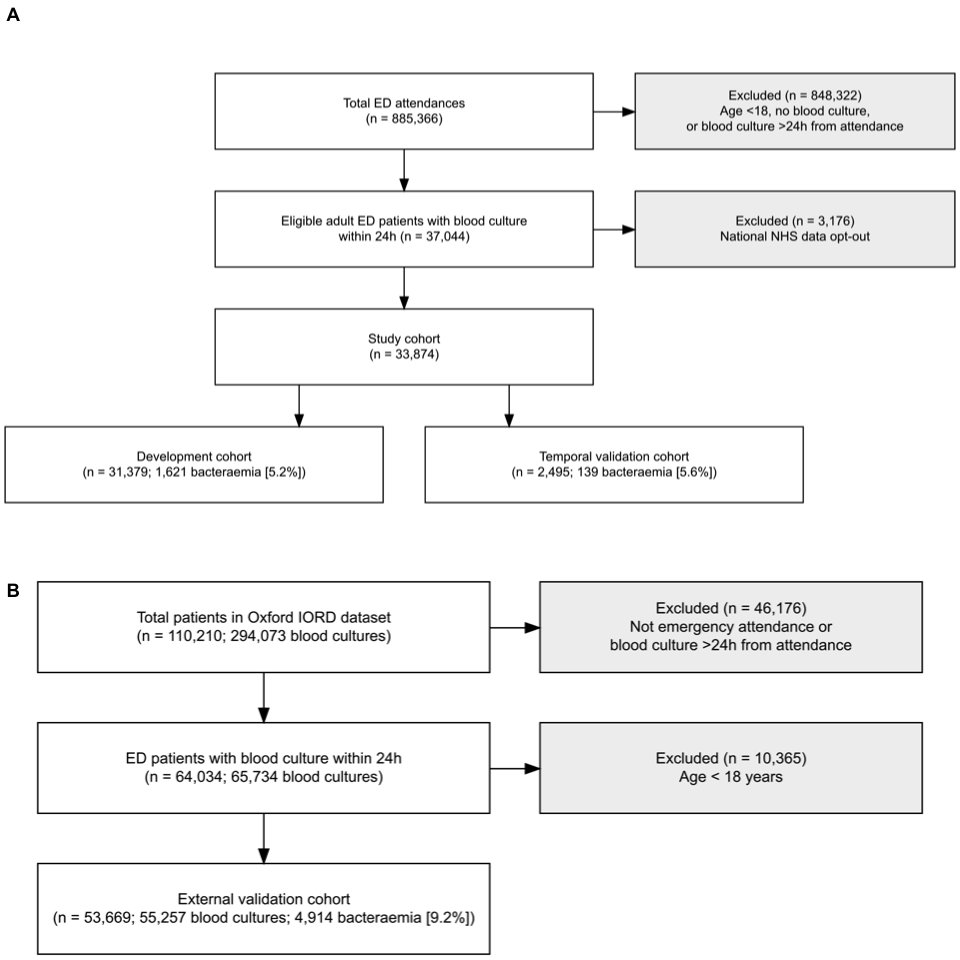

###### Supplementary Table S1: Participant characteristics stratified by development and validation cohort

| **Characteristic** | **Development**  N = 31,379^1^ | **Validation**  N = 2,495^1^ | **Overall**  N = 33,874^1^ |
| --- | --- | --- | --- |
| Bacteraemia | 1,621 (5.2%) | 139 (5.6%) | 1,760 (5.2%) |
| Missing, n (%) | 0 | 0 | 0 |
| Age | 55 (35, 72) | 57 (37, 72) | 56 (35, 72) |
| Missing, n (%) | 0 | 0 | 0 |
| Chronic cardiac comorbidity | 6,173 (23%) | 519 (26%) | 6,692 (23%) |
| Missing, n (%) | 4,689 | 500 | 5,189 |
| Chronic liver comorbidity | 425 (1.6%) | 40 (2.0%) | 465 (1.6%) |
| Missing, n (%) | 4,689 | 500 | 5,189 |
| Diabetes mellitus | 3,808 (14%) | 276 (14%) | 4,084 (14%) |
| Missing, n (%) | 4,689 | 500 | 5,189 |
| Systolic blood pressure | 129 (116, 144) | 129 (116, 145) | 129 (116, 144) |
| Missing, n (%) | 353 | 34 | 387 |
| Diastolic blood pressure | 74 (65, 83) | 73 (65, 82) | 74 (65, 83) |
| Missing, n (%) | 368 | 34 | 402 |
| Heart rate | 99 (85, 115) | 99 (85, 115) | 99 (85, 115) |
| Missing, n (%) | 118 | 7 | 125 |
| Temperature | 37.10 (36.60, 38.00) | 36.90 (36.60, 37.80) | 37.10 (36.60, 38.00) |
| Missing, n (%) | 114 | 7 | 121 |
| Peripheral oxygen saturation | 97.00 (96.00, 99.00) | 97.00 (96.00, 99.00) | 97.00 (96.00, 99.00) |
| Missing, n (%) | 148 | 7 | 155 |
| On oxygen? | 2,998 (9.7%) | 156 (6.3%) | 3,154 (9.4%) |
| Missing, n (%) | 322 | 14 | 336 |
| Albumin | 41 (37, 45) | 40 (36, 43) | 41 (37, 45) |
| Missing, n (%) | 3,171 | 291 | 3,462 |
| alp | 89 (69, 122) | 94 (72, 131) | 89 (69, 122) |
| Missing, n (%) | 4,659 | 435 | 5,094 |
| C-reactive protein | 40 (10, 109) | 49 (12, 114) | 41 (10, 109) |
| Missing, n (%) | 1,214 | 98 | 1,312 |
| Eosinophil count | 0.04 (0.01, 0.12) | 0.04 (0.01, 0.12) | 0.04 (0.01, 0.12) |
| Missing, n (%) | 886 | 43 | 929 |
| Lactate | 1.30 (0.90, 1.90) | 1.20 (0.80, 1.80) | 1.24 (0.90, 1.80) |
| Missing, n (%) | 13,161 | 285 | 13,446 |
| Lymphocyte count | 1.14 (0.70, 1.75) | 1.13 (0.69, 1.73) | 1.14 (0.70, 1.75) |
| Missing, n (%) | 873 | 42 | 915 |
| Neutrophil count | 7.0 (4.3, 10.6) | 7.2 (4.4, 11.0) | 7.0 (4.3, 10.7) |
| Missing, n (%) | 871 | 42 | 913 |
| Platelet count | 239 (177, 311) | 236 (173, 314) | 239 (177, 311) |
| Missing, n (%) | 840 | 40 | 880 |
| Potassium | 4.10 (3.90, 4.50) | 4.20 (3.90, 4.50) | 4.20 (3.90, 4.50) |
| Missing, n (%) | 4,412 | 394 | 4,806 |
| Urea | 5.1 (3.6, 7.4) | 5.5 (3.8, 7.8) | 5.1 (3.6, 7.4) |
| Missing, n (%) | 14,260 | 1,181 | 15,441 |
| ^1^n (%); Median (Q1, Q3) | | | |

###### Supplementary Table S2: Missingness of model variables in the UCLH cohort

Missingness is shown for all model predictor variables and key demographic variables, overall and stratified by the primary outcome. The primary outcome (bacteraemia) has no missing values as it is derived directly from the blood culture record. IMD = Index of Multiple Deprivation.

|  | **Missing: N (%)** | | |
| --- | --- | --- | --- |
| Variable | Overall | Bacteraemia | No bacteraemia |
| **Model predictors** | | | |
| Age | 0 (0%) | 0 (0%) | 0 (0%) |
| Chronic cardiac comorbidity | 5189 (15.3%) | 125 (7.1%) | 5064 (15.8%) |
| Systolic blood pressure | 387 (1.1%) | 13 (0.7%) | 374 (1.2%) |
| Diastolic blood pressure | 402 (1.2%) | 15 (0.9%) | 387 (1.2%) |
| Heart rate | 125 (0.4%) | 7 (0.4%) | 118 (0.4%) |
| Peripheral oxygen saturation | 155 (0.5%) | 9 (0.5%) | 146 (0.5%) |
| On oxygen | 336 (1%) | 13 (0.7%) | 323 (1%) |
| Temperature | 121 (0.4%) | 7 (0.4%) | 114 (0.4%) |
| Albumin | 3462 (10.2%) | 103 (5.9%) | 3359 (10.5%) |
| Alkaline phosphatase | 5094 (15%) | 194 (11%) | 4900 (15.3%) |
| C-reactive protein | 1312 (3.9%) | 51 (2.9%) | 1261 (3.9%) |
| Eosinophil count | 929 (2.7%) | 59 (3.4%) | 870 (2.7%) |
| Lactate | 13446 (39.7%) | 667 (37.9%) | 12779 (39.8%) |
| Lymphocyte count | 915 (2.7%) | 59 (3.4%) | 856 (2.7%) |
| Neutrophil count | 913 (2.7%) | 59 (3.4%) | 854 (2.7%) |
| Platelet count | 880 (2.6%) | 43 (2.4%) | 837 (2.6%) |
| Potassium | 4806 (14.2%) | 262 (14.9%) | 4544 (14.1%) |
| Urea | 15441 (45.6%) | 675 (38.4%) | 14766 (46%) |
| **Demographic variables** | | | |
| Sex | 12 (0%) | 0 (0%) | 12 (0%) |
| Ethnicity | 7017 (20.7%) | 322 (18.3%) | 6695 (20.8%) |
| IMD quintile | 6618 (19.5%) | 189 (10.7%) | 6429 (20%) |

###### Supplementary Table S3: Infection diagnoses in UCLH cohort

Infection diagnoses were assigned based on the primary ICD-10 code for that participant hospital encounter. Participants discharged directly from the Emergency Department had no codes assigned (discharge diagnosis = unknown). Site of infection was assigned based on the ICD-10 schema provided in the supplementary files.

| **Characteristic** | **Bacteraemia**  N = 1,760^1^ | **No bacteraemia**  N = 32,114^1^ |
| --- | --- | --- |
| Discharge diagnosis |  |  |
| Infection | 1,031 (7.6%) | 12,529 (92%) |
| Not infection | 604 (4.0%) | 14,521 (96%) |
| Unknown | 125 | 5,064 |
| Site of infection |  |  |
| Bone and Joint infections | 5 (6.8%) | 68 (93%) |
| Cardiovascular infections | 15 (54%) | 13 (46%) |
| Gastrointestinal infections | 44 (4.4%) | 963 (96%) |
| Genital infections | 12 (5.7%) | 200 (94%) |
| Hepatobiliary infections | 53 (15%) | 289 (85%) |
| Lower respiratory infections | 98 (1.7%) | 5,602 (98%) |
| Multisystem or other infectious disease | 204 (14%) | 1,296 (86%) |
| Neurological infections | 13 (7.6%) | 158 (92%) |
| Ocular infections | 0 (0%) | 25 (100%) |
| Sepsis | 448 (31%) | 985 (69%) |
| Skin and soft tissue infections | 24 (2.8%) | 835 (97%) |
| Upper respiratory and ENT infections | 12 (1.1%) | 1,126 (99%) |
| Urinary infections | 104 (8.9%) | 1,062 (91%) |
| Unknown | 728 | 19,492 |
| ^1^n (%) | | |

###### Supplementary Table S4: Patient outcomes stratified by bacteraemia status

| **Characteristic** | **Bacteraemia**  N = 1,760^1^ | **No bacteraemia**  N = 32,114^1^ |
| --- | --- | --- |
| Hospital length of stay (days) | 8 (4, 16) | 3 (0, 7) |
| Intensive care admission | 266 (16%) | 2,267 (8.4%) |
| Intensive care length of stay (days) | 4 (2, 8) | 4 (2, 8) |
| Days since hospital admission to ICU admission | 0.5 (0.2, 1.9) | 0.4 (0.2, 2.2) |
| Death during inpatient stay | 160 (9.1%) | 1,432 (4.5%) |
| Death within 30 days | 179 (10%) | 1,814 (5.7%) |
| Days since hospital admission to death | 9 (2, 17) | 13 (5, 23) |
| ^1^Median (Q1, Q3); n (%) | | |

###### Supplementary Table S5: Blood culture characteristics

| **Characteristic** | **Negative**  N = 33,625^1^ | **Contaminant**  N = 1,663^1^ | **Positive**  N = 2,044^1^ |
| --- | --- | --- | --- |
| Time from attendance to blood culture (hours) | 0.9 (0.4 to 3.7) | 0.8 (0.3 to 2.4) | 0.8 (0.3 to 4.0) |
| Total blood culture (per encounter) |  |  |  |
| 1 | 26,564 (93%) | 1,121 (3.9%) | 1,017 (3.5%) |
| 2 | 4,709 (85%) | 314 (5.7%) | 523 (9.4%) |
| 3 | 1,445 (78%) | 118 (6.3%) | 299 (16%) |
| 4+ | 907 (74%) | 110 (9.0%) | 205 (17%) |
| ^1^Median (Q1 to Q3); n (%) | | | |

###### Supplementary Figure S2: Pathogenic organisms isolated from blood cultures

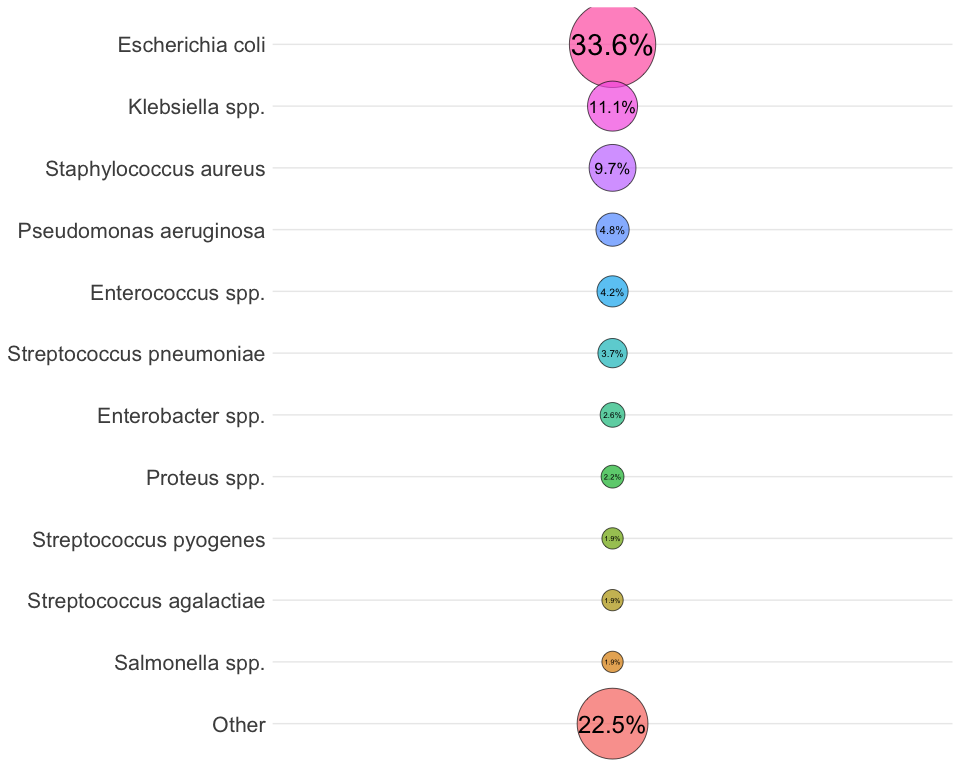

#### S2: Model development

###### Supplementary Table S6: Model coefficients

| Variable | Estimate | 95% Confidence Interval |
| --- | --- | --- |
| Intercept | -18.29 | -24.07 to -12.5 |
| Age | 0.01 | 0.01 to 0.02 |
| Age (spline 1) | -0.01 | -0.02 to 0 |
| Chronic cardiac comorbidity: Yes | 0.33 | 0.19 to 0.48 |
| Chronic liver comorbidity: Yes | 0.49 | 0.18 to 0.79 |
| Diabetes mellitus: Yes | 0.24 | 0.1 to 0.38 |
| Systolic BP | 0.00 | -0.01 to 0 |
| Systolic BP (spline 1) | 0.01 | 0 to 0.02 |
| Diastolic BP | -0.02 | -0.03 to -0.01 |
| Diastolic BP (spline 1) | 0.01 | 0 to 0.02 |
| Pulse | 0.01 | 0.01 to 0.02 |
| Pulse (spline 1) | -0.01 | -0.02 to 0 |
| Temperature | 0.11 | -0.02 to 0.24 |
| Temperature (spline 1) | 0.36 | 0.17 to 0.56 |
| SpO2 | 0.10 | 0.07 to 0.13 |
| SpO2 (spline 1) | -0.04 | -0.1 to 0.01 |
| On oxygen = Yes | -0.36 | -0.53 to -0.19 |
| Albumin | 0.00 | -0.02 to 0.01 |
| Albumin (spline 1) | -0.05 | -0.07 to -0.02 |
| Alkaline phosphatase | 0.01 | 0.01 to 0.01 |
| Alkaline phosphatase (spline 1) | -0.01 | -0.02 to -0.01 |
| C-reactive protein | 0.01 | 0.01 to 0.01 |
| C-reactive protein (spline 1) | -0.02 | -0.02 to -0.01 |
| Eosinophil count | -3.35 | -5.09 to -1.6 |
| Eosinophil count (spline 1) | 7.09 | 3.2 to 10.97 |
| Lactate | 0.69 | 0.45 to 0.92 |
| Lactate (spline 1) | -0.84 | -1.18 to -0.5 |
| Lymphocyte count | -1.34 | -1.5 to -1.17 |
| Lymphocyte count (spline 1) | 1.26 | 1.11 to 1.41 |
| Neutrophil count | 0.14 | 0.11 to 0.18 |
| Neutrophil count (spline 1) | -0.12 | -0.15 to -0.09 |
| Platelet count | 0.00 | 0 to 0 |
| Platelet count (spline 1) | 0.00 | 0 to 0 |
| Potassium | -0.19 | -0.41 to 0.02 |
| Potassium (spline 1) | -0.05 | -0.28 to 0.17 |
| Urea | 0.15 | 0.08 to 0.22 |
| Urea (spline 1) | -0.16 | -0.25 to -0.07 |
| Restricted cubic spline knot positions are: | | |
| - Age = 25, 55, 82; | | |
| - Systolic BP = 105, 129, 165; | | |
| - Diastolic BP = 58, 74, 91; | | |
| - Pulse = 73, 99, 130; | | |
| - Temperature = 36.3, 37.1, 38.7. | | |
| - SpO2 = 94, 97, 100. | | |
| - Albumin = 32, 41, 48; | | |
| - ALP = 55, 89, 189; | | |
| - CRP = 2.6, 40.5, 202. | | |
| - Eosinophils = 0, 0, 0.3; | | |
| - Lactate = 0.7, 1.2, 2.8; | | |
| - Lymphocyte count = 0.4, 1.1, 2.5. | | |
| - Neutrophil count = 2.3, 6.9, 15. | | |
| - Platelet count = 112, 239, 401. | | |
| - Potassium = 3.6, 4.2, 4.8; | | |
| - Urea = 2.7, 5, 11. | | |

###### Supplementary Table S7: Variable selection frequency across multiply imputed datasets

Variables selected in ≥5 datasets were included in the final model.

| Variable | Number of MI datasets selected |
| --- | --- |
| Age | 9 |
| Blood albumin | 10 |
| Blood alkaline phosphatase | 10 |
| Chronic cardiac comorbidity | 10 |
| Blood C-reactive protein | 10 |
| Diastolic blood pressure | 10 |
| Diabetes mellitus | 10 |
| Blood eosinophil count | 10 |
| Blood lactate | 10 |
| Chronic liver comorbidity | 10 |
| Blood lymphocyte count | 10 |
| Blood neutrophil count | 10 |
| On oxygen? (at time of Spo2 measurement) | 8 |
| Blood platelet count | 10 |
| Blood potassium | 10 |
| Heart rate | 10 |
| Respiratory rate | 3 |
| Peripheral oxygen saturation | 10 |
| Systolic blood pressure | 8 |
| Temperature | 10 |
| Blood urea | 10 |

###### Supplementary Table S8: Timing of predictor measurement in the UCLH cohort

Time from hospital attendance to measurement of each model predictor. Predictors are shown individually, by type (vital signs and blood tests), and in aggregate. Systolic blood pressure and diastolic blood pressure are shown together as Blood pressure, as these values were invariably measured together. IQR = interquartile range.

| Predictor | Time from attendance (hrs)*^1^* | Measured within 6 hours (%) |
| --- | --- | --- |
| Albumin | 1.3 (0.8 to 2.3) | 89.4% |
| Alkaline phosphatase | 1.3 (0.8 to 2.3) | 88.9% |
| Blood pressure | 0.3 (0.1 to 0.8) | 99% |
| C-reactive protein | 1.2 (0.8 to 2) | 92.2% |
| Eosinophils | 1.1 (0.7 to 2.1) | 92.3% |
| Heart rate | 0 (-0.1 to 0.4) | 99.5% |
| Lactate | 0.8 (0.4 to 1.5) | 93.6% |
| Lymphocytes | 1.1 (0.7 to 2) | 92.6% |
| Neutrophils | 1.1 (0.7 to 2) | 92.6% |
| On oxygen? | 0.1 (0 to 0.5) | 98.9% |
| Platelet count | 1.1 (0.7 to 2) | 92.6% |
| Potassium | 1.3 (0.8 to 2.2) | 90.7% |
| SpO2 | 0 (0 to 0.4) | 99.5% |
| Temperature | 0 (0 to 0.4) | 99.4% |
| Urea | 1.2 (0.8 to 2.1) | 88.1% |
| Blood tests | 1.2 (0.7 to 2.1) | 91.4% |
| Vital signs | 0.1 (0 to 0.5) | 99.3% |
| All predictors | 0.8 (0.3 to 1.6) | 94.4% |
| *^1^*Median (IQR) | | |

###### Supplementary Figure S3: Distribution of time from hospital attendance to predictor measurement in the UCLH cohort

Density plots are shown for each model predictor, truncated at 6 hours. Systolic blood pressure and diastolic blood pressure are shown together as Blood pressure, as these values were invariably measured together.

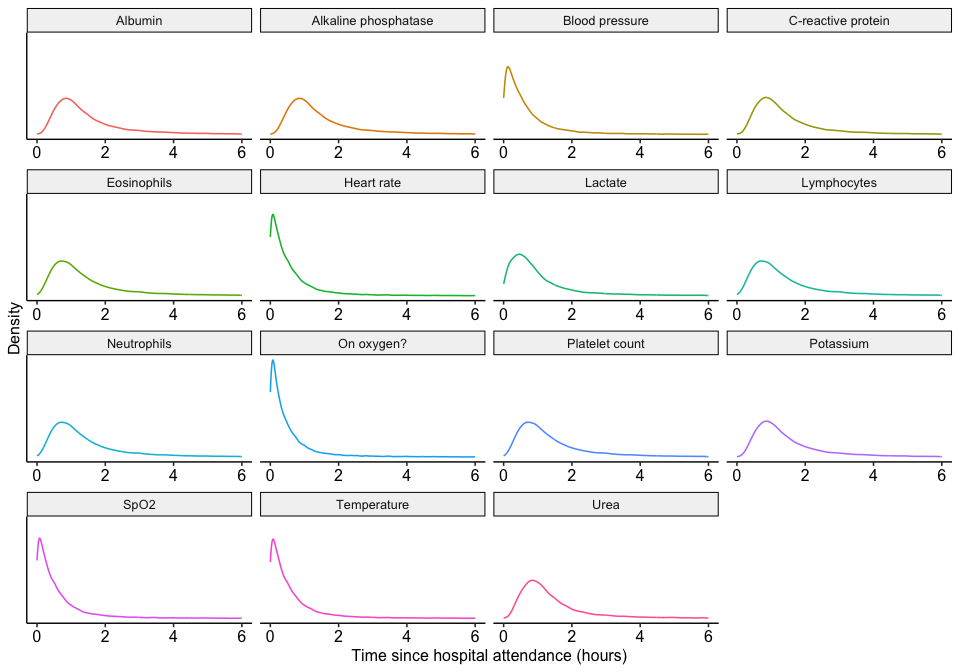

#### S3: Internal validation

###### Supplementary Figure S4: Internal-external cross-validation (IECV) meta-analysis, by time of attendance

Pooled estimates are calculated through random-effects meta-analysis (total sample size = 31,379 participants in the UCLH development cohort). Time of attendance is split into quarter years. Dashed lines indicate lines of perfect calibration-in-the-large (0) and slope (1), respectively. Black squares indicate point estimates; bars indicate 95% confidence intervals; diamonds indicate pooled random-effects meta-analysis estimates. I2 values for c-statistic, calibration-in-the-large and calibration slope are shown in the figure footer.

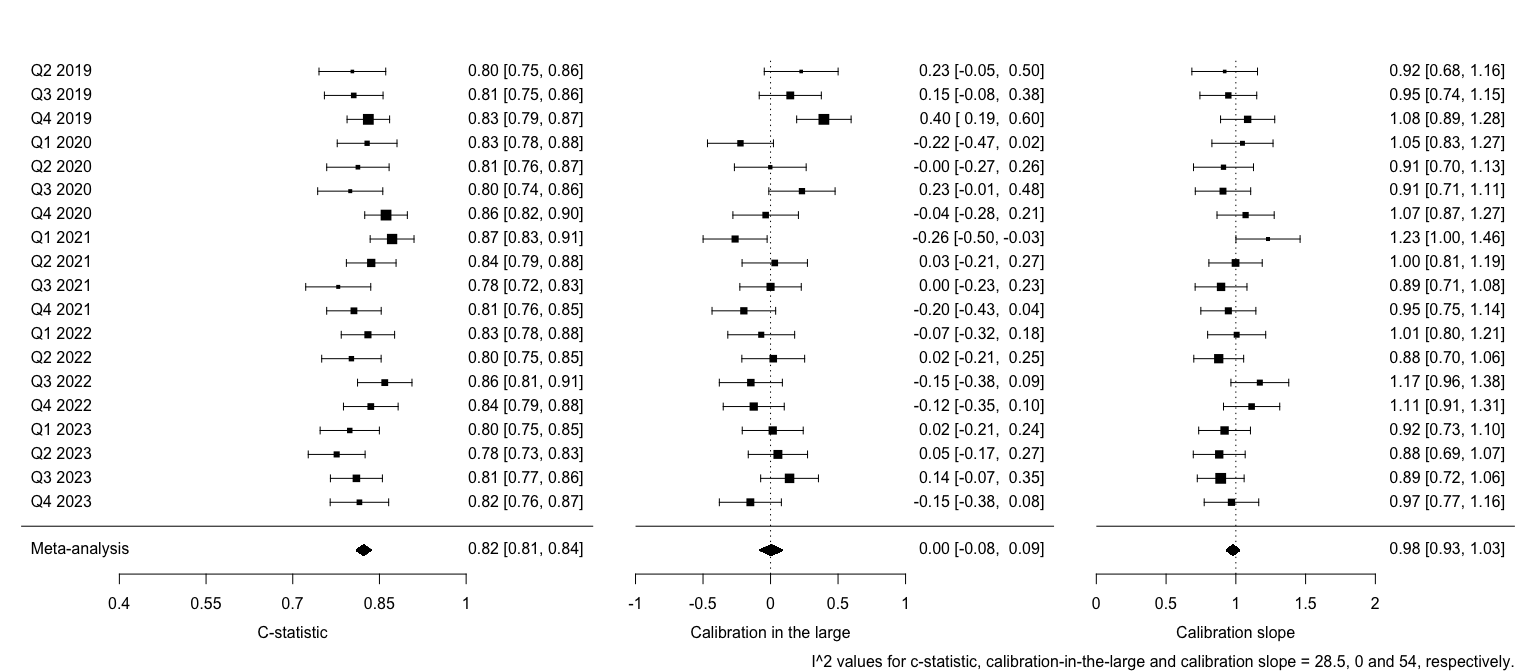

###### Supplementary Figure S5: IECV calibration plot

Calibration is plotted using a loess smoother, by IECV time period. Rug plots, shown on the x-axis, plot the distribution of predicted risk.

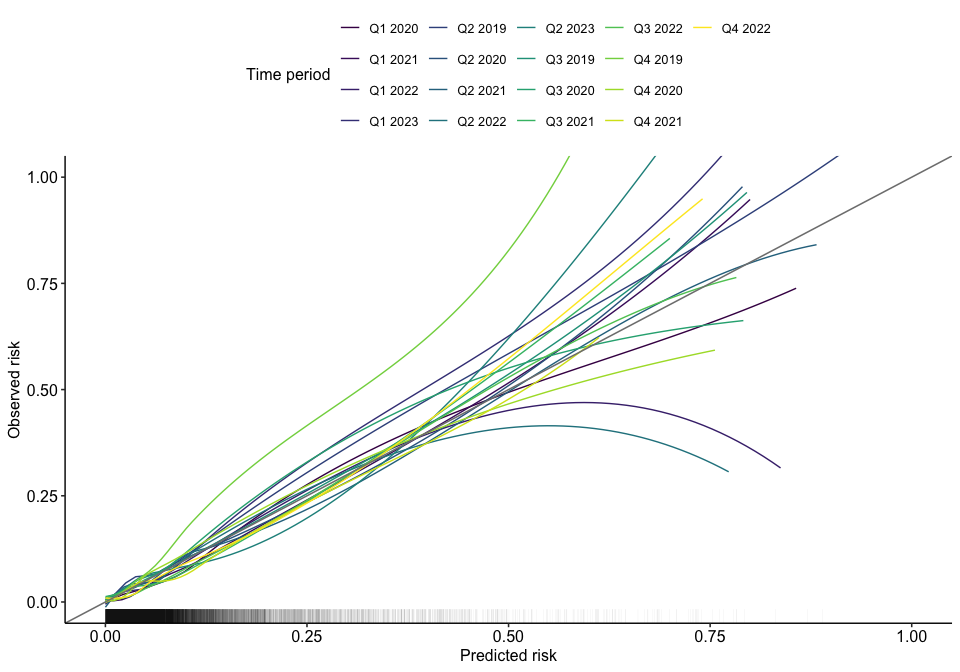

###### Supplementary Figure S6: Model prediction instability

Prediction instability plot showing model predictions plotted against bootstrapped ‘alternative’ model predictions. In the development data, 1000 bootstrapped datasets equal in sample size to the original data were sampled with replacement. The model development process (variable selection and model training) was repeated on each of these bootstrapped datasets, creating 1000 bootstrapped models. These bootstrapped models were then used to create 1000 sets of alternative bootstrapped predictions on the original dataset. The black line shows a line of best fit calculated by a generalised additive model with smoothing using random effects. The back dashed lines are lines of best fit, calculated using the same method, against the 5th and 95th centile of the bootstrapped predictions. Mean average prediction error (MAPE) was 0.009 (inter-quartile range 0.001 to 0.009).

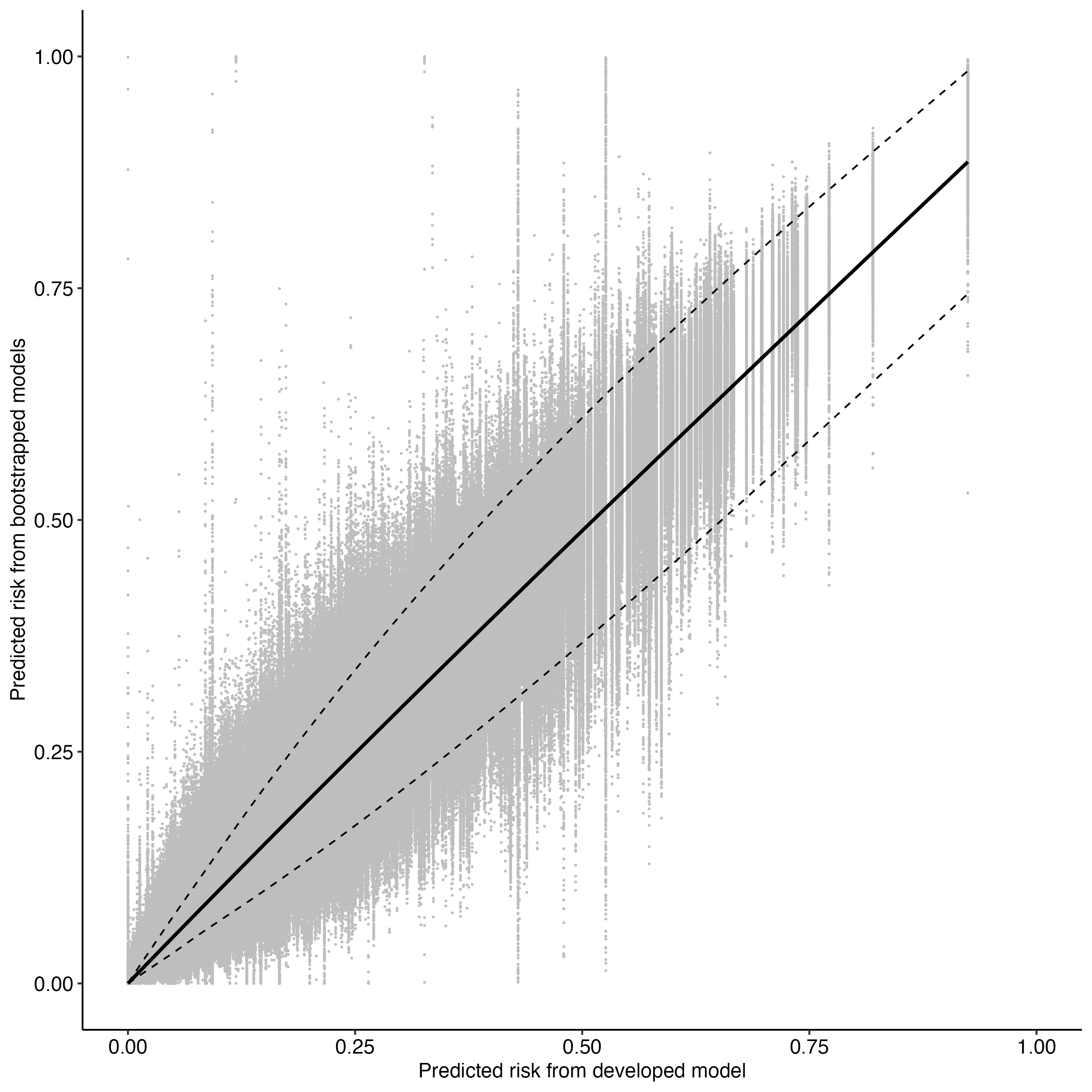

###### Supplementary Figure S6b: Individual-patient prediction instability

Individual-patient prediction instability plots adapted from Riley et al. (2025). Twelve patients were selected at risk levels across the predicted risk distribution. For each patient, the distribution of bootstrap model predictions is shown as a half-violin plot; the IQR box and 95% interval line summarise the spread. The horizontal coloured line extends to the right from each patient’s main-model predicted risk, indicating its position within the bootstrapped distribution. Panel (A) shows the full 0–1 prediction range. Panel (B) zooms into the 0–0.1 range, where the majority of the development data and clinically relevant predictions lie.

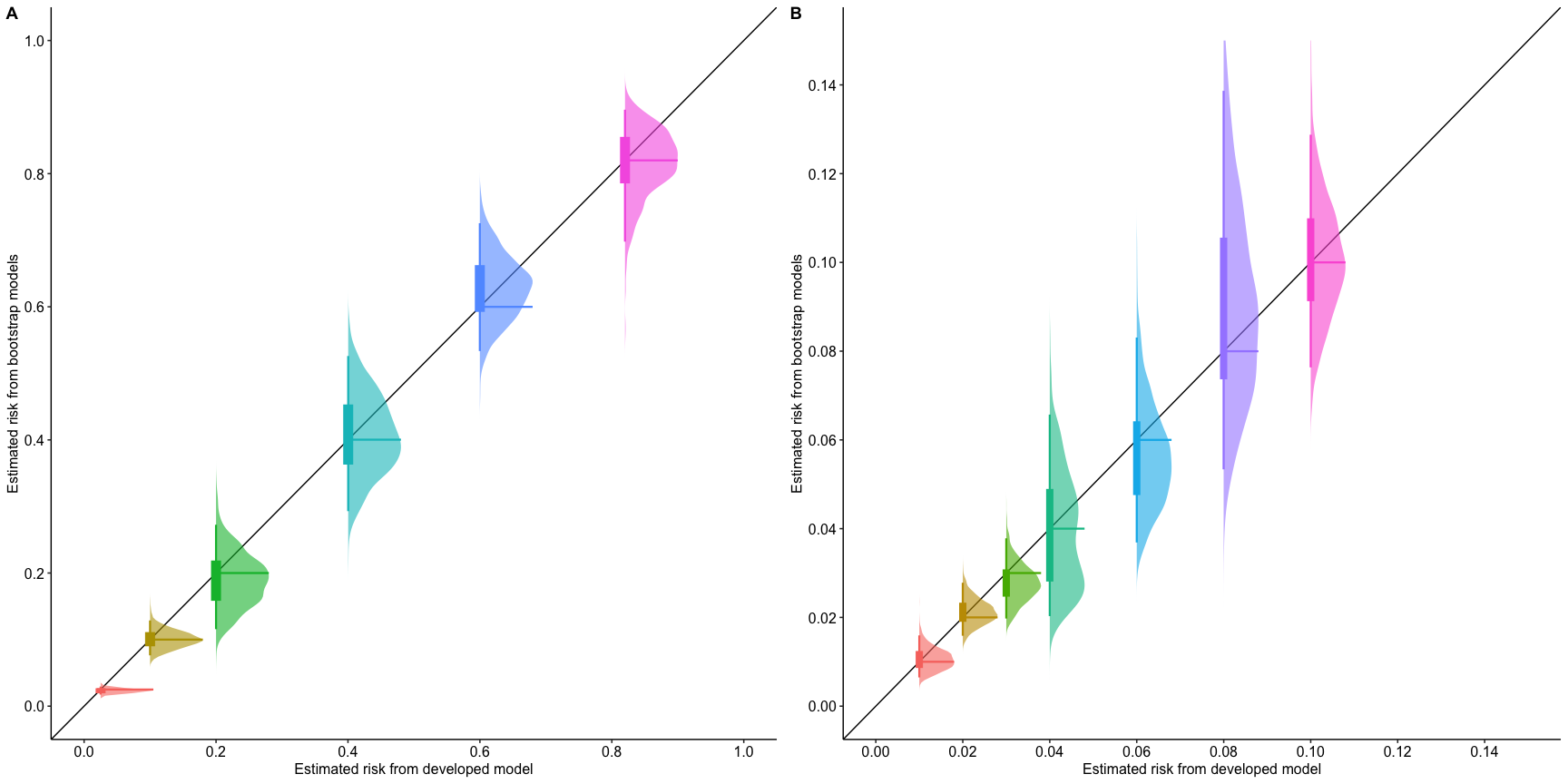

#### S4: Additional temporal validation analyses

###### Supplementary Table S9: Model performance by subgroup — UCLH held-out temporal validation cohort

Age was categorised by quartile in the whole UCLH dataset. CITL = calibration-in-the-large; N = number of participants.

| Variable | C-statistic | Calibration Slope | CITL | Observed | Expected | N |
| --- | --- | --- | --- | --- | --- | --- |
| Age category | | | | | | |
| 18-36 | 0.72 (0.58 - 0.85) | 0.75 (0.26 - 1.23) | -0.11 (-0.62 - 0.4) | 16 | 18 | 651 |
| 37-56 | 0.83 (0.76 - 0.9) | 1.07 (0.72 - 1.42) | 0.21 (-0.18 - 0.59) | 31 | 26 | 628 |
| 57-71 | 0.87 (0.81 - 0.93) | 1.21 (0.86 - 1.56) | -0.23 (-0.59 - 0.13) | 36 | 43 | 614 |
| 72+ | 0.81 (0.74 - 0.88) | 1.05 (0.77 - 1.34) | 0.19 (-0.11 - 0.48) | 56 | 48 | 602 |
| Ethnicity | | | | | | |
| Asian or Asian British | 0.94 (0.9 - 0.98) | 1.85 (1.07 - 2.63) | 0.49 (-0.06 - 1.05) | 16 | 11 | 204 |
| Black or Black British | 0.8 (0.56 - 1.03) | 1.11 (0.45 - 1.78) | -0.37 (-1.16 - 0.43) | 7 | 10 | 207 |
| Other ethnic group | 0.83 (0.72 - 0.94) | 1.11 (0.64 - 1.58) | 0.2 (-0.33 - 0.72) | 17 | 15 | 281 |
| Unknown | 0.74 (0.63 - 0.84) | 0.75 (0.45 - 1.05) | -0.04 (-0.43 - 0.34) | 30 | 31 | 654 |
| White | 0.84 (0.79 - 0.89) | 1.09 (0.85 - 1.32) | -0.01 (-0.27 - 0.25) | 69 | 69 | 1149 |
| Sex | | | | | | |
| Female | 0.86 (0.81 - 0.91) | 1.24 (0.97 - 1.51) | -0.07 (-0.34 - 0.21) | 59 | 62 | 1319 |
| Male | 0.8 (0.74 - 0.85) | 0.91 (0.7 - 1.11) | 0.11 (-0.13 - 0.36) | 80 | 73 | 1176 |
| IMD quintile | | | | | | |
| Q1 - Most deprived | 0.85 (0.76 - 0.93) | 1.2 (0.76 - 1.63) | -0.13 (-0.59 - 0.32) | 22 | 25 | 419 |
| Q2 | 0.79 (0.71 - 0.87) | 0.96 (0.68 - 1.25) | 0.14 (-0.2 - 0.47) | 43 | 38 | 660 |
| Q3 | 0.83 (0.75 - 0.91) | 0.99 (0.62 - 1.35) | -0.11 (-0.53 - 0.31) | 26 | 28 | 440 |
| Q4 | 0.87 (0.79 - 0.95) | 1.45 (0.82 - 2.08) | 0.44 (-0.06 - 0.95) | 19 | 13 | 224 |
| Q5 - Least deprived | 0.84 (0.66 - 1.02) | 0.97 (0.34 - 1.6) | -0.12 (-0.94 - 0.7) | 7 | 8 | 142 |

###### Supplementary Table S10: Univariable predictor performance

C-statistic for each model predictor fitted as a univariable logistic regression model with restricted cubic splines for continuous variables, trained and evaluated in multiply imputed UCLH development data.

| Variable | C-statistic (95% CI) |
| --- | --- |
| C-reactive protein | 0.7 (0.65 to 0.74) |
| Lymphocyte count | 0.69 (0.65 to 0.73) |
| Eosinophil count | 0.68 (0.63 to 0.72) |
| Urea | 0.67 (0.62 to 0.72) |
| Albumin | 0.67 (0.63 to 0.72) |
| Lactate | 0.65 (0.6 to 0.7) |
| Age | 0.63 (0.58 to 0.67) |
| Neutrophil count | 0.62 (0.57 to 0.67) |
| Chronic cardiac comorbidity | 0.62 (0.57 to 0.66) |
| Temperature | 0.61 (0.56 to 0.67) |
| Platelet count | 0.61 (0.56 to 0.66) |
| Alkaline phosphatase | 0.61 (0.56 to 0.66) |
| Potassium | 0.58 (0.52 to 0.63) |
| Diastolic blood pressure | 0.57 (0.52 to 0.63) |
| Systolic blood pressure | 0.55 (0.5 to 0.61) |
| Diabetes mellitus | 0.55 (0.51 to 0.59) |
| Heart rate | 0.55 (0.5 to 0.6) |
| On oxygen? (at time of Spo2 measurement) | 0.53 (0.5 to 0.56) |
| Peripheral oxygen saturation | 0.52 (0.48 to 0.57) |
| Chronic liver comorbidity | 0.51 (0.49 to 0.53) |

###### Supplementary Table S11: Alternative predictor and risk score performance in UCLH held-out temporal validation cohort

C-statistic for alternative predictors (C-reactive protein) and clinical risk scores (NEWS2, qSOFA, CURB65) evaluated in multiply imputed UCLH held-out temporal validation data.

| Variable | C-statistic (95% CI) |
| --- | --- |
| C-reactive protein | 0.7 (0.65 to 0.74) |
| CURB65 score | 0.65 (0.61 to 0.7) |
| NEWS2 score | 0.62 (0.57 to 0.67) |
| qSOFA score | 0.59 (0.55 to 0.64) |

###### Supplementary Table S12: Model performance — WHO priority pathogens

Pathogens included in this analysis are those represented in the UCLH dataset that correspond to species listed in the 2024 WHO Bacterial Priority Pathogens List. The analysis compares model performance in culture-negative patients against those with a positive blood culture growing a WHO-listed organism, as used in Ming et al. 2025 in the Lancet Digital Health. CITL = calibration-in-the-large

Organism

**Gram-negative**

*Escherichia coli*

*Klebsiella spp.*

*Pseudomonas aeruginosa / Pseudomonas spp.*

*Proteus spp.*

*Serratia spp.*

*Citrobacter spp.*

**Gram-positive**

*Enterococcus spp.*

*Staphylococcus aureus*

*Streptococcus agalactiae*

*Streptococcus dysgalactiae*

*Streptococcus pneumoniae*

*Streptococcus pyogenes*

*Streptococcus gallolyticus*

*Anginosus group Streptococci*

| Validation data | C-statistic | Calibration Slope | CITL |
| --- | --- | --- | --- |
| UCLH temporal validation - WHO pathogens only | 0.85 (0.81 - 0.89) | 1.14 (0.96 - 1.32) | -0.1 (-0.3 - 0.09) |

###### Supplementary Table S13: Comparison with Lee et al. model

For the probability reconstruction method, the original variable coefficients were extracted from Lee et al’s model, a model intercept was derived from our validation data, and a regression model constructed to allow the calculation of predicted risk. This allowed for calculation of calibration metrics. Given Lee et al’s model was developed to predict bacteraemia in respiratory infections only, we limited performance analysis to respiratory infections in our validation dataset, defined as previously by ICD10 codes. The performance of the IDEAS-Bacteraemia model (IDEAS-Bact) is shown for comparison. CITL = calibration-in-the-large.

| Model | Cohort | Reconstruction method | C-statistic | Calibration slope | CITL |
| --- | --- | --- | --- | --- | --- |
| IDEAS-Bact | UCH temporal validation - All | Probability | 0.83 (0.79 - 0.87) | 1.05 (0.89 - 1.21) | 0.03 (-0.15 - 0.21) |
| IDEAS-Bact | UCH temporal validation - Respiratory Infections only | Probability | 0.82 (0.71 - 0.92) | 0.9 (0.28 - 1.51) | -0.78 (-1.46 - -0.1) |
| Lee | UCH temporal validation - Respiratory Infections only | Point Score | 0.75 (0.64 - 0.86) | - | - |
| Lee | UCH temporal validation - Respiratory Infections only | Probability | 0.75 (0.63 - 0.86) | 0.67 (0.03 - 1.31) | 0.3 (-0.38 - 0.98) |

###### Supplementary Figure S7: Decision curve analysis in held-out temporal validation data at UCLH, respiratory patients only

Net benefit is shown with a loess smoother for the IDEAS-Bacteraemia model (IDEAS-Bact) and for Lee et al.’s model in participants from the held-out temporal validation cohort at UCLH with respiratory infection only, compared to test-all (All) and test-none (None). Vertical dashed line represents the number needed to test below which Lee et al.’s model becomes a superior strategy to the IDEAS-Bacteraemia model, for respiratory infection.

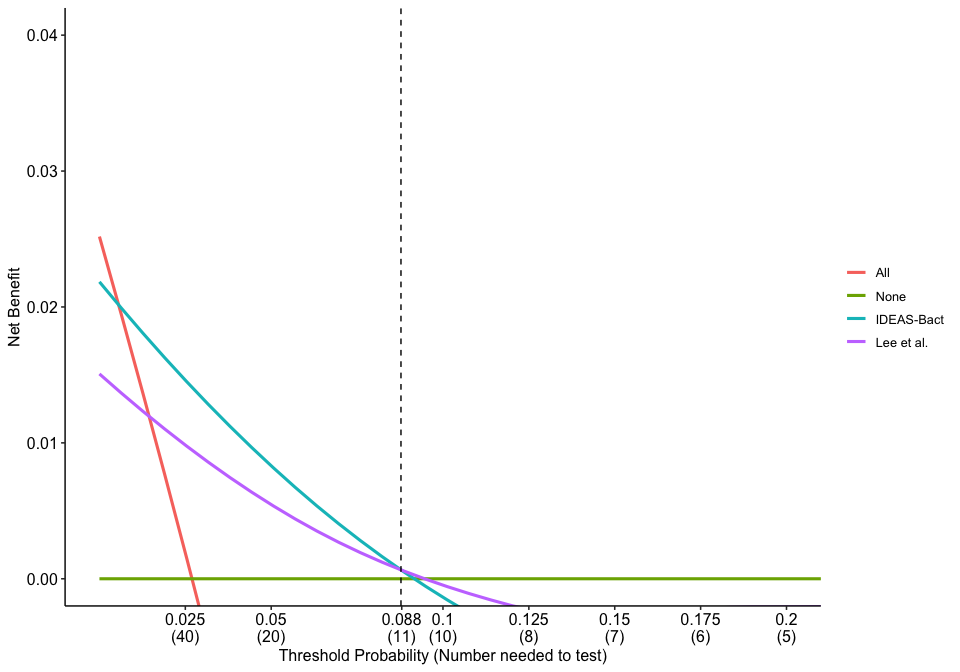

#### S5: External validation

###### Supplementary Table S14: External validation participant characteristics

|  | **Bacteraemia** | **No bacteraemia** | **Overall** |
| --- | --- | --- | --- |
| Variable | n = 4,765; (8.9%) | n = 48,904; (91.1%) | n = 53,669 |
| Age (years) | 77 (65 to 85) | 71 (50 to 82) | 71 (52 to 83) |
| Sex |  |  |  |
| Female | 2065 (7.9%) | 24046 (92.1%) | 26111 |
| Male | 2700 (9.8%) | 24858 (90.2%) | 27558 |
| Ethnicity |  |  |  |
| Asian or Asian British | 80 (5.6%) | 1358 (94.4%) | 1438 |
| Black or Black British | 38 (6.6%) | 536 (93.4%) | 574 |
| Chinese or other ethnic group | 52 (8.8%) | 540 (91.2%) | 592 |
| Mixed | 24 (5.5%) | 411 (94.5%) | 435 |
| Unknown | 730 (9.6%) | 6898 (90.4%) | 7628 |
| White | 3841 (8.9%) | 39161 (91.1%) | 43002 |
| IMD quintile |  |  |  |
| Q1 (Most deprived) | 290 (8.1%) | 3280 (91.9%) | 3570 |
| Q2 | 411 (7.9%) | 4766 (92.1%) | 5177 |
| Q3 | 843 (8.8%) | 8762 (91.2%) | 9605 |
| Q4 | 1381 (9.1%) | 13820 (90.9%) | 15201 |
| Q5 (Least deprived) | 1805 (9.2%) | 17783 (90.8%) | 19588 |
| Cardiac comorbidity | 1484 (10.5%) | 12636 (89.5%) | 14120 |
| Diabetes mellitus | 937 (11.2%) | 7436 (88.8%) | 8373 |
| Liver comorbidity | 131 (11.3%) | 1031 (88.7%) | 1162 |
| Systolic blood pressure (mmHg) | 121 (102 to 141) | 130 (113 to 147) | 129 (112 to 147) |
| Diastolic blood pressure (mmHg) | 65 (54 to 77) | 72 (61 to 83) | 71 (60 to 83) |
| Heart rate (bpm) | 102 (88 to 117) | 96 (82 to 111) | 96 (82 to 112) |
| SpO2 (%) | 96 (94 to 98) | 96 (94 to 98) | 96 (94 to 98) |
| On oxygen? | 1470 (10.9%) | 12045 (89.1%) | 13515 |
| Temperature (°C) | 37.7 (36.8 to 38.4) | 37.2 (36.4 to 38) | 37.2 (36.4 to 38) |
| Albumin (g/L) | 30 (26 to 34) | 34 (29 to 37) | 33 (29 to 37) |
| Alkaline phosphatase (U/L) | 111 (81 to 171) | 90 (71 to 120) | 91 (71 to 124) |
| C-reactive protein (mg/L) | 130.55 (48.88 to 240.02) | 53.8 (15.3 to 129.5) | 58.4 (16.7 to 140) |
| Eosinophils (×10⁹/L) | 0.01 (0 to 0.03) | 0.03 (0.01 to 0.11) | 0.03 (0.01 to 0.1) |
| Lactate (mmol/L) | 2.1 (1.4 to 3.5) | 1.4 (1 to 2.1) | 1.5 (1 to 2.2) |
| Lymphocytes (×10⁹/L) | 0.53 (0.31 to 0.9) | 1.05 (0.66 to 1.62) | 1 (0.61 to 1.57) |
| Neutrophils (×10⁹/L) | 11.79 (7.95 to 16.54) | 8.95 (5.96 to 12.83) | 9.16 (6.06 to 13.14) |
| Platelets (×10⁹/L) | 209 (158 to 280) | 246 (191 to 315) | 243.5 (187 to 313) |
| Potassium (mmol/L) | 4 (3.6 to 4.5) | 4.1 (3.7 to 4.5) | 4.1 (3.7 to 4.5) |
| Urea (mmol/L) | 8.7 (6 to 13.2) | 6.2 (4.4 to 9.4) | 6.4 (4.5 to 9.7) |

###### Supplementary Table S15: Comparison of model predictions and predictor distributions between UCLH and Oxford cohorts

Model predictions are shown as median (interquartile range) of predicted probability; UCLH predictions are derived from the whole UCLH cohort. Continuous predictors are shown as median (interquartile range). Binary predictors are shown as n (%). Bacteraemia prevalence is shown separately.

|  | **UCLH** | **Oxford external validation** |
| --- | --- | --- |
| Variable | n = 33,874 | n = 53,669 |
| **Model predictions** | | |
| Median predicted probability, bacteraemia (IQR) | 0.12 (0.05 to 0.24) | 0.19 (0.1 to 0.32) |
| Median predicted probability, no bacteraemia (IQR) | 0.02 (0.01 to 0.05) | 0.04 (0.02 to 0.1) |
| **Participant characteristics** | | |
| Bacteraemia, n (%) | 1,760 (5.2%) | 4,765 (8.9%) |
| Age (years) | 56 (35 to 72) | 71 (52 to 83) |
| Cardiac comorbidity | 6692 (23.3%) | 14120 (26.3%) |
| Diabetes mellitus | 4084 (14.2%) | 8373 (15.6%) |
| Liver comorbidity | 465 (1.6%) | 1162 (2.2%) |
| Systolic blood pressure (mmHg) | 129 (116 to 144) | 129 (112 to 147) |
| Diastolic blood pressure (mmHg) | 74 (65 to 83) | 71 (60 to 83) |
| Heart rate (bpm) | 99 (85 to 115) | 96 (82 to 112) |
| SpO2 (%) | 97 (96 to 99) | 96 (94 to 98) |
| On oxygen? | 3154 (9.4%) | 13515 (25.2%) |
| Temperature (°C) | 37.1 (36.6 to 38) | 37.2 (36.4 to 38) |
| Albumin (g/L) | 41 (37 to 45) | 33 (29 to 37) |
| Alkaline phosphatase (U/L) | 89 (69 to 122) | 91 (71 to 124) |
| C-reactive protein (mg/L) | 41 (10.4 to 109.2) | 58.4 (16.7 to 140) |
| Eosinophils (×10⁹/L) | 0.04 (0.01 to 0.12) | 0.03 (0.01 to 0.1) |
| Lactate (mmol/L) | 1.24 (0.9 to 1.8) | 1.5 (1 to 2.2) |
| Lymphocytes (×10⁹/L) | 1.14 (0.7 to 1.75) | 1 (0.61 to 1.57) |
| Neutrophils (×10⁹/L) | 6.96 (4.27 to 10.67) | 9.16 (6.06 to 13.14) |
| Platelets (×10⁹/L) | 239 (177 to 311) | 243.5 (187 to 313) |
| Potassium (mmol/L) | 4.2 (3.9 to 4.5) | 4.1 (3.7 to 4.5) |
| Urea (mmol/L) | 5.1 (3.6 to 7.4) | 6.4 (4.5 to 9.7) |

###### Supplementary Table S16: External validation — missingness in model variables

Model variables are shown in descending order of missingness

| Variable | Missing (%) |
| --- | --- |
| Lactate (mmol/L) | 11.1 |
| C-reactive protein (mg/L) | 9.9 |
| Alkaline phosphatase (U/L) | 8.0 |
| Albumin (g/L) | 7.7 |
| Potassium (mmol/L) | 7.2 |
| Temperature (°C) | 6.3 |
| Diastolic blood pressure (mmHg) | 6.1 |
| Systolic blood pressure (mmHg) | 5.9 |
| SpO2 (%) | 5.8 |
| Heart rate (bpm) | 5.7 |
| Eosinophils (×10⁹/L) | 4.5 |
| Lymphocytes (×10⁹/L) | 4.5 |
| Neutrophils (×10⁹/L) | 4.5 |
| Platelets (×10⁹/L) | 3.8 |
| Urea (mmol/L) | 3.6 |
| Age (years) | 0.0 |
| Cardiac comorbidity | 0.0 |
| Liver comorbidity | 0.0 |
| Diabetes mellitus | 0.0 |
| On oxygen? | 0.0 |

###### Supplementary Table S17: External validation — subgroup analysis

Age categories defined by quartile in UCLH data. CITL = calibration-in-the-large

| Category | C-statistic | Calibration slope | CITL | Observed events | Expected events | Sample size |
| --- | --- | --- | --- | --- | --- | --- |
| Age | | | | | | |
| 18-36 | 0.82 (0.79 - 0.85) | 1.12 (0.99 - 1.25) | -0.34 (-0.48 - -0.2) | 219 | 295 | 7312 |
| 37-56 | 0.83 (0.82 - 0.85) | 1.07 (0.99 - 1.16) | -0.1 (-0.2 - -0.01) | 547 | 594 | 8895 |
| 57-71 | 0.83 (0.82 - 0.84) | 1.09 (1.02 - 1.16) | -0.01 (-0.08 - 0.06) | 999 | 1009 | 10711 |
| 72+ | 0.81 (0.8 - 0.82) | 1.05 (1.01 - 1.09) | 0.07 (0.03 - 0.11) | 3000 | 2836 | 26751 |
| Ethnicity | | | | | | |
| Asian or Asian British | 0.82 (0.77 - 0.86) | 1.06 (0.83 - 1.28) | -0.19 (-0.43 - 0.05) | 80 | 93 | 1438 |
| Black or Black British | 0.82 (0.74 - 0.89) | 1.12 (0.77 - 1.47) | -0.01 (-0.36 - 0.34) | 38 | 38 | 574 |
| Other ethnic group | 0.85 (0.8 - 0.9) | 1.22 (0.96 - 1.47) | 0.12 (-0.13 - 0.38) | 76 | 69 | 1027 |
| Unknown | 0.84 (0.82 - 0.85) | 1.11 (1.03 - 1.19) | 0.13 (0.04 - 0.21) | 730 | 661 | 7628 |
| White | 0.83 (0.82 - 0.83) | 1.08 (1.04 - 1.11) | -0.01 (-0.05 - 0.02) | 3841 | 3874 | 43002 |
| IMD quintile | | | | | | |
| Q1 (Most deprived) | 0.84 (0.82 - 0.87) | 1.13 (1 - 1.25) | 0.01 (-0.12 - 0.14) | 290 | 288 | 3570 |
| Q2 | 0.82 (0.8 - 0.84) | 1.02 (0.92 - 1.12) | -0.05 (-0.15 - 0.06) | 411 | 426 | 5177 |
| Q3 | 0.83 (0.81 - 0.84) | 1.08 (1.01 - 1.16) | 0.04 (-0.04 - 0.11) | 843 | 820 | 9605 |
| Q4 | 0.82 (0.81 - 0.84) | 1.06 (1.01 - 1.12) | 0.03 (-0.03 - 0.09) | 1381 | 1351 | 15201 |
| Q5 (Least deprived) | 0.83 (0.82 - 0.84) | 1.1 (1.05 - 1.15) | 0 (-0.06 - 0.05) | 1805 | 1811 | 19588 |
| Sex | | | | | | |
| Female | 0.84 (0.83 - 0.85) | 1.12 (1.07 - 1.17) | 0.01 (-0.04 - 0.06) | 2065 | 2049 | 26111 |
| Male | 0.82 (0.81 - 0.83) | 1.05 (1.01 - 1.09) | 0.01 (-0.04 - 0.05) | 2700 | 2687 | 27558 |

### ICD-10 Code Sets and Organism Definitions

#### Site of Infection

ICD-10 codes used to classify the primary site of infection. Codes are grouped by infection system. Where an infection type sub-classification is available, this is shown.

##### Bone and Joint infections

| **ICD-10 code** | **Description** | **Infection type** |
| --- | --- | --- |
| A18.0 |  | Infection of bones and joints |
| A54.4 |  | Infection of bones and joints |
| B45.3 |  | Infection of bones and joints |
| B67.2 |  | Infection of bones and joints |
| B90.2 |  | Infection of bones and joints |
| M00 |  | Infection of bones and joints |
| M01 |  | Infection of bones and joints |
| M46.2 |  | Infection of bones and joints |
| M46.3 | Infection of intervertebral disc (pyogenic) | Infection of bones and joints |
| M46.4 | Discitis, unspecified | Infection of bones and joints |
| M46.5 | Other infective spondylopathies | Infection of bones and joints |
| M49.0 |  | Infection of bones and joints |
| M49.1 | Brucella spondylitis | Infection of bones and joints |
| M49.2 | Enterobacterial spondylitis | Infection of bones and joints |
| M49.3 | Spondylopathy in other infectious and parasitic diseases classified elsewhere | Infection of bones and joints |
| M86 | Osteomyelitis | Infection of bones and joints |
| M90.0 |  | Infection of bones and joints |
| M90.1 | Periostitis in other infectious diseases classified elsewhere | Infection of bones and joints |
| M90.2 | Osteopathy in other infectious diseases classified elsewhere | Infection of bones and joints |

##### Cardiovascular infections

| **ICD-10 code** | **Description** | **Infection type** |
| --- | --- | --- |
| A39.5 |  | Infections of the Heart |
| A52.0 |  | Infections of the Heart |
| B33.2 |  | Infections of the Heart |
| B37.6 |  | Infections of the Heart |
| I30.1 | Infective pericarditis | Infections of the Heart |
| I32.0 | Pericarditis in bacterial diseases classified elsewhere | Infections of the Heart |
| I32.1 | Pericarditis in other infectious and parasitic diseases classified elsewhere | Infections of the Heart |
| I33.0 | Acute and subacute infective endocarditis | Infections of the Heart |
| I40.0 | Infective myocarditis | Infections of the Heart |
| I41.0 | Myocarditis in bacterial diseases classified elsewhere | Infections of the Heart |
| I41.1 | Myocarditis in viral diseases classified elsewhere | Infections of the Heart |
| I41.2 | Myocarditis in other infectious and parasitic diseases classified elsewhere | Infections of the Heart |
| I43.0 | Cardiomyopathy in infectious and parasitic diseases classified elsewhere | Infections of the Heart |
| I98.0 | Cardiovascular syphilis | Infections of the Heart |
| I98.1 | Cardiovascular disorders in other infectious and parasitic diseases classified elsewhere | Infections of the Heart |

##### Gastrointestinal infections

| **ICD-10 code** | **Description** | **Infection type** |
| --- | --- | --- |
| A00 | Cholera | Infections of the digestive system |
| A01 | Typhoid and paratyphoid fevers | Infections of the digestive system |
| A02.0 |  | Infections of the digestive system |
| A03 | Shigellosis | Infections of the digestive system |
| A04 | Other bacterial intestinal infections | Infections of the digestive system |
| A05 | Other bacterial foodborne intoxications, not elsewhere classified | Infections of the digestive system |
| A06.0 |  | Infections of the digestive system |
| A06.1 |  | Infections of the digestive system |
| A06.2 |  | Infections of the digestive system |
| A06.3 |  | Infections of the digestive system |
| A07 | Other protozoal intestinal diseases | Infections of the digestive system |
| A08 |  | Infections of the digestive system |
| A09 | Other gastroenteritis and colitis of infectious and unspecified origin | Infections of the digestive system |
| A18.3 |  | Infections of the digestive system |
| A21.3 |  | Infections of the digestive system |
| A22.2 |  | Infections of the digestive system |
| A51.1 |  | Infection of anal and rectal regions |
| A54.6 |  | Infection of anal and rectal regions |
| A56.3 |  | Infection of anal and rectal regions |
| A60.1 |  | Infection of anal and rectal regions |
| A60.9 |  | Infection of anal and rectal regions |
| A63.0 | Anogenital (venereal) warts | Infection of anal and rectal regions |
| B05.4 |  | Infections of the digestive system |
| B46.2 |  | Infections of the digestive system |
| B78.0 |  | Infections of the digestive system |
| B81 | Other intestinal helminthiases, not elsewhere classified | Infections of the digestive system |
| B82 | Unspecified intestinal parasitism | Infections of the digestive system |
| B98.0 | Helicobacter pylori [H.pylori] as the cause of diseases classified to other chapters | Infections of the digestive system |
| K23.0 |  | Infections of the digestive system |
| K23.1 | Megaoesophagus in Chagas' disease | Infections of the digestive system |
| K25.1 |  | Peritonitis |
| K25.2 |  | Peritonitis |
| K25.5 |  | Peritonitis |
| K25.6 |  | Peritonitis |
| K26.1 |  | Peritonitis |
| K26.2 |  | Peritonitis |
| K26.5 |  | Peritonitis |
| K26.6 |  | Peritonitis |
| K27.1 |  | Peritonitis |
| K27.2 |  | Peritonitis |
| K27.5 |  | Peritonitis |
| K27.6 |  | Peritonitis |
| K28.1 |  | Peritonitis |
| K28.2 |  | Peritonitis |
| K28.5 |  | Peritonitis |
| K28.6 |  | Peritonitis |
| K35 |  | Appendicitis |
| K35.2 |  | Peritonitis |
| K35.3 |  | Peritonitis |
| K36 |  | Appendicitis |
| K37 |  | Appendicitis |
| K57.0 |  | Peritonitis |
| K57.2 |  | Peritonitis |
| K57.4 |  | Peritonitis |
| K57.8 |  | Peritonitis |
| K61 | Abscess of anal and rectal regions | Infection of anal and rectal regions |
| K63.0 | Abscess of intestine | Infections of the digestive system |
| K63.1 |  | Peritonitis |
| K65 |  | Peritonitis |
| K67 |  | Peritonitis |
| K93.0 |  | Infections of the digestive system |

##### Genital infections

| **ICD-10 code** | **Description** | **Infection type** |
| --- | --- | --- |
| A18.1 |  | Infection of other or unspecified genitourinary system |
| A51.0 |  | Infection of other or unspecified genitourinary system |
| A54.0 |  | Infection of other or unspecified genitourinary system |
| A54.1 |  | Infection of other or unspecified genitourinary system |
| A54.2 |  | Infection of other or unspecified genitourinary system |
| A56.0 |  | Infection of other or unspecified genitourinary system |
| A56.1 |  | Infection of other or unspecified genitourinary system |
| A56.2 |  | Infection of other or unspecified genitourinary system |
| A57 | Chancroid | Infection of other or unspecified genitourinary system |
| A58 | Granuloma inguinale | Infection of other or unspecified genitourinary system |
| A59.0 |  | Infection of other or unspecified genitourinary system |
| A60.0 |  | Infection of other or unspecified genitourinary system |
| B26.0 |  | Infection of male genital system |
| B37.3 |  | Infection of other or unspecified genitourinary system |
| B37.4 |  | Infection of other or unspecified genitourinary system |
| B90.1 |  | Infection of other or unspecified genitourinary system |
| N41.0 | Acute prostatitis | Infection of male genital system |
| N41.2 | Abscess of prostate | Infection of male genital system |
| N41.3 | Prostatocystitis | Infection of male genital system |
| N43.1 | Infected hydrocele | Infection of male genital system |
| N45 | Orchitis and epididymitis | Infection of male genital system |
| N48.1 | Balanoposthitis | Infection of male genital system |
| N70 | Salpingitis and oophoritis | Female pelvic inflammatory disease |
| N71 | Inflammatory disease of uterus, except cervix | Female pelvic inflammatory disease |
| N72 | Inflammatory disease of cervix uteri | Female pelvic inflammatory disease |
| N73 | Other female pelvic inflammatory diseases | Female pelvic inflammatory disease |
| N74 |  | Female pelvic inflammatory disease |
| N75.1 | Abscess of Bartholin's gland | Infection of other or unspecified genitourinary system |
| N77.0 | Ulceration of vulva in infectious and parasitic diseases classified elsewhere | Infection of other or unspecified genitourinary system |
| N77.1 | Vaginitis, vulvitis and vulvovaginitis in infectious and parasitic diseases classified elsewhere | Infection of other or unspecified genitourinary system |

##### Hepatobiliary infections

| **ICD-10 code** | **Description** | **Infection type** |
| --- | --- | --- |
| A06.4 |  | Infection of liver |
| B15 | Acute hepatitis A | Infection of liver |
| B16 | Acute hepatitis B | Infection of liver |
| B17 | Other acute viral hepatitis | Infection of liver |
| B18 |  | Infection of liver |
| B19 | Unspecified viral hepatitis | Infection of liver |
| B25.1 |  | Infection of liver |
| B58.1 |  | Infection of liver |
| B67.0 |  | Infection of liver |
| B67.5 |  | Infection of liver |
| B67.8 |  | Infection of liver |
| B94.2 | Sequelae of viral hepatitis | Infection of liver |
| K75.0 | Abscess of liver | Infection of liver |
| K77.0 | Liver disorders in infectious and parasitic diseases classified elsewhere | Infection of liver |
| K80.0 |  | Cholecystitis |
| K80.1 |  | Cholecystitis |
| K80.3 |  | Cholangitis |
| K80.4 |  | Cholecystitis |
| K81 |  | Cholecystitis |
| K83.0 |  | Cholangitis |

##### Lower respiratory infections

| **ICD-10 code** | **Description** | **Infection type** |
| --- | --- | --- |
| A06.5 |  | Lower Respiratory Tract Infections |
| A15 | Respiratory tuberculosis, bacteriologically and histologically confirmed | Lower Respiratory Tract Infections |
| A15 | Respiratory tuberculosis, bacteriologically and histologically confirmed | Respiratory Tract Infection |
| A15.0 | Tuberculosis of lung, confirmed by sputum microscopy with or without culture | Respiratory Tract Infection |
| A15.1 | Tuberculosis of lung, confirmed by culture only | Respiratory Tract Infection |
| A15.2 | Tuberculosis of lung, confirmed histologically | Respiratory Tract Infection |
| A15.3 | Tuberculosis of lung, confirmed by unspecified means | Respiratory Tract Infection |
| A15.4 | Tuberculosis of intrathoracic lymph nodes, confirmed bacteriologically and histologically | Respiratory Tract Infection |
| A15.5 | Tuberculosis of larynx, trachea and bronchus, confirmed bacteriologically and histologically | Respiratory Tract Infection |
| A15.6 | Tuberculous pleurisy, confirmed bacteriologically and histologically | Respiratory Tract Infection |
| A15.7 | Primary respiratory tuberculosis, confirmed bacteriologically and histologically | Respiratory Tract Infection |
| A15.8 | Other respiratory tuberculosis, confirmed bacteriologically and histologically | Respiratory Tract Infection |
| A15.9 | Respiratory tuberculosis unspecified, confirmed bacteriologically and histologically | Respiratory Tract Infection |
| A16 | Respiratory tuberculosis, not confirmed bacteriologically or histologically | Lower Respiratory Tract Infections |
| A16 | Respiratory tuberculosis, not confirmed bacteriologically or histologically | Respiratory Tract Infection |
| A16.0 | Tuberculosis of lung, bacteriologically and histologically negative | Respiratory Tract Infection |
| A16.1 | Tuberculosis of lung, bacteriological and histological examination not done | Respiratory Tract Infection |
| A16.2 | Tuberculosis of lung, without mention of bacteriological or histological confirmation | Respiratory Tract Infection |
| A16.3 | Tuberculosis of intrathoracic lymph nodes, without mention of bacteriological or histological confirmation | Respiratory Tract Infection |
| A16.4 | Tuberculosis of larynx, trachea and bronchus, without mention of bacteriological or histological confirmation | Respiratory Tract Infection |
| A16.5 | Tuberculous pleurisy, without mention of bacteriological or histological confirmation | Respiratory Tract Infection |
| A16.7 | Primary respiratory tuberculosis without mention of bacteriological or histological confirmation | Respiratory Tract Infection |
| A16.8 | Other respiratory tuberculosis, without mention of bacteriological or histological confirmation | Respiratory Tract Infection |
| A16.9 | Respiratory tuberculosis unspecified, without mention of bacteriological or histological confirmation | Respiratory Tract Infection |
| A20.2 |  | Lower Respiratory Tract Infections |
| A21.2 |  | Lower Respiratory Tract Infections |
| A22.1 |  | Lower Respiratory Tract Infections |
| A31.0 |  | Lower Respiratory Tract Infections |
| A37 | Whooping cough | Lower Respiratory Tract Infections |
| A42.0 |  | Lower Respiratory Tract Infections |
| A43.0 |  | Lower Respiratory Tract Infections |
| A48.1 |  | Respiratory Tract Infection |
| B01.2 |  | Lower Respiratory Tract Infections |
| B05.2 |  | Lower Respiratory Tract Infections |
| B25.0 |  | Lower Respiratory Tract Infections |
| B37.1 |  | Lower Respiratory Tract Infections |
| B38.0 |  | Lower Respiratory Tract Infections |
| B38.1 |  | Lower Respiratory Tract Infections |
| B38.2 |  | Lower Respiratory Tract Infections |
| B39.0 |  | Lower Respiratory Tract Infections |
| B39.1 |  | Lower Respiratory Tract Infections |
| B39.2 |  | Lower Respiratory Tract Infections |
| B40.0 |  | Lower Respiratory Tract Infections |
| B40.1 |  | Lower Respiratory Tract Infections |
| B40.2 |  | Lower Respiratory Tract Infections |
| B41.0 |  | Lower Respiratory Tract Infections |
| B42.0 |  | Lower Respiratory Tract Infections |
| B44.0 | Invasive pulmonary aspergillosis | Lower Respiratory Tract Infections |
| B44.1 | Other pulmonary aspergillosis | Lower Respiratory Tract Infections |
| B45.0 |  | Lower Respiratory Tract Infections |
| B46.0 |  | Lower Respiratory Tract Infections |
| B58.3 |  | Lower Respiratory Tract Infections |
| B59 | Pneumocystosis | Lower Respiratory Tract Infections |
| B67.1 |  | Lower Respiratory Tract Infections |
| J10.0 |  | Lower Respiratory Tract Infections |
| J11.0 |  | Lower Respiratory Tract Infections |
| J12 | Viral pneumonia, not elsewhere classified | Lower Respiratory Tract Infections |
| J12 | Viral pneumonia, not elsewhere classified | Respiratory Tract Infection |
| J12.0 |  | Respiratory Tract Infection |
| J12.1 |  | Respiratory Tract Infection |
| J12.2 |  | Respiratory Tract Infection |
| J12.3 |  | Respiratory Tract Infection |
| J12.8 |  | Respiratory Tract Infection |
| J12.9 |  | Respiratory Tract Infection |
| J13 | Pneumonia due to Streptococcus pneumoniae | Lower Respiratory Tract Infections |
| J13 | Pneumonia due to Streptococcus pneumoniae | Respiratory Tract Infection |
| J14 | Pneumonia due to Haemophilus influenzae | Lower Respiratory Tract Infections |
| J14 | Pneumonia due to Haemophilus influenzae | Respiratory Tract Infection |
| J15 | Bacterial pneumonia, not elsewhere classified | Lower Respiratory Tract Infections |
| J15 | Bacterial pneumonia, not elsewhere classified | Respiratory Tract Infection |
| J15.0 |  | Respiratory Tract Infection |
| J15.1 |  | Respiratory Tract Infection |
| J15.2 |  | Respiratory Tract Infection |
| J15.3 |  | Respiratory Tract Infection |
| J15.4 |  | Respiratory Tract Infection |
| J15.5 |  | Respiratory Tract Infection |
| J15.6 |  | Respiratory Tract Infection |
| J15.7 |  | Respiratory Tract Infection |
| J15.8 |  | Respiratory Tract Infection |
| J15.9 |  | Respiratory Tract Infection |
| J16 |  | Lower Respiratory Tract Infections |
| J16 |  | Respiratory Tract Infection |
| J16.0 | Chlamydial pneumonia | Respiratory Tract Infection |
| J16.8 | Pneumonia due to other specified infectious organisms | Respiratory Tract Infection |
| J17 |  | Lower Respiratory Tract Infections |
| J17 |  | Respiratory Tract Infection |
| J17.0 | Pneumonia in bacterial diseases classified elsewhere | Respiratory Tract Infection |
| J17.1 | Pneumonia in viral diseases classified elsewhere | Respiratory Tract Infection |
| J17.2 | Pneumonia in mycoses | Respiratory Tract Infection |
| J17.3 | Pneumonia in parasitic diseases | Respiratory Tract Infection |
| J17.8 | Pneumonia in other diseases classified elsewhere | Respiratory Tract Infection |
| J18 | Pneumonia, organism unspecified | Lower Respiratory Tract Infections |
| J18 | Pneumonia, organism unspecified | Respiratory Tract Infection |
| J18.0 |  | Respiratory Tract Infection |
| J18.1 |  | Respiratory Tract Infection |
| J18.2 |  | Respiratory Tract Infection |
| J18.8 |  | Respiratory Tract Infection |
| J18.9 |  | Respiratory Tract Infection |
| J20 |  | Lower Respiratory Tract Infections |
| J20 |  | Respiratory Tract Infection |
| J20.0 | Acute bronchitis due to Mycoplasma pneumoniae | Respiratory Tract Infection |
| J20.1 | Acute bronchitis due to Haemophilus influenzae | Respiratory Tract Infection |
| J20.2 | Acute bronchitis due to streptococcus | Respiratory Tract Infection |
| J20.3 | Acute bronchitis due to coxsackievirus | Respiratory Tract Infection |
| J20.4 | Acute bronchitis due to parainfluenza virus | Respiratory Tract Infection |
| J20.5 | Acute bronchitis due to respiratory syncytial virus | Respiratory Tract Infection |
| J20.6 | Acute bronchitis due to rhinovirus | Respiratory Tract Infection |
| J20.7 | Acute bronchitis due to echovirus | Respiratory Tract Infection |
| J20.8 | Acute bronchitis due to other specified organisms | Respiratory Tract Infection |
| J20.9 | Acute bronchitis, unspecified | Respiratory Tract Infection |
| J21 |  | Lower Respiratory Tract Infections |
| J21 |  | Respiratory Tract Infection |
| J21.0 | Acute bronchiolitis due to respiratory syncytial virus | Respiratory Tract Infection |
| J21.1 | Acute bronchiolitis due to human metapneumovirus | Respiratory Tract Infection |
| J21.8 | Acute bronchiolitis due to other specified organisms | Respiratory Tract Infection |
| J21.9 | Acute bronchiolitis, unspecified | Respiratory Tract Infection |
| J22 | Unspecified acute lower respiratory infection | Lower Respiratory Tract Infections |
| J22 | Unspecified acute lower respiratory infection | Respiratory Tract Infection |
| J40 |  | Respiratory Tract Infection |
| J44.0 | Chronic obstructive pulmonary disease with acute lower respiratory infection | Lower Respiratory Tract Infections |
| J44.0 | Chronic obstructive pulmonary disease with acute lower respiratory infection | Respiratory Tract Infection |
| J44.1 | Chronic obstructive pulmonary disease with acute exacerbation, unspecified | Lower Respiratory Tract Infections |
| J44.1 | Chronic obstructive pulmonary disease with acute exacerbation, unspecified | Respiratory Tract Infection |
| J65 | Pneumoconiosis associated with tuberculosis | Lower Respiratory Tract Infections |
| J69.0 |  | Aspiration pneumonitis |
| J69.1 |  | Aspiration pneumonitis |
| J69.8 |  | Aspiration pneumonitis |
| J85 | Abscess of lung and mediastinum | Respiratory Tract Infection |
| J85.0 |  | Lower Respiratory Tract Infections |
| J85.0 |  | Respiratory Tract Infection |
| J85.1 |  | Lower Respiratory Tract Infections |
| J85.1 |  | Respiratory Tract Infection |
| J85.2 |  | Lower Respiratory Tract Infections |
| J85.2 |  | Respiratory Tract Infection |
| J85.3 | Abscess of mediastinum | Respiratory Tract Infection |
| J86 | Pyothorax | Lower Respiratory Tract Infections |
| J86 | Pyothorax | Respiratory Tract Infection |
| J86.0 |  | Respiratory Tract Infection |
| J86.9 |  | Respiratory Tract Infection |
| P23 |  | Lower Respiratory Tract Infections |
| U071 |  | COVID-19 infection |

##### Neurological infections

| **ICD-10 code** | **Description** | **Infection type** |
| --- | --- | --- |
| A06.6 |  | Other nervous system infections |
| A17.0 |  | Meningitis |
| A17.1 |  | Meningitis |
| A17.8 |  | Other nervous system infections |
| A17.9 |  | Other nervous system infections |
| A20.3 |  | Meningitis |
| A32.1 |  | Meningitis |
| A39.0 |  | Meningitis |
| A52.1 |  | Other nervous system infections |
| A52.2 |  | Other nervous system infections |
| A52.3 |  | Other nervous system infections |
| A80 | Acute poliomyelitis | Other nervous system infections |
| A81 |  | Other nervous system infections |
| A82 | Rabies | Other nervous system infections |
| A83 | Mosquito-borne viral encephalitis | Encephalitis |
| A84 | Tick-borne viral encephalitis | Encephalitis |
| A85 | Other viral encephalitis, not elsewhere classified | Encephalitis |
| A86 | Unspecified viral encephalitis | Encephalitis |
| A87 | Viral meningitis | Meningitis |
| A88 | Other viral infections of central nervous system, not elsewhere classified | Other nervous system infections |
| A89 | Unspecified viral infection of central nervous system | Other nervous system infections |
| B00.3 |  | Meningitis |
| B00.4 |  | Encephalitis |
| B01.0 |  | Meningitis |
| B01.1 |  | Encephalitis |
| B02.0 |  | Encephalitis |
| B02.1 |  | Meningitis |
| B02.2 |  | Other nervous system infections |
| B05.0 |  | Encephalitis |
| B05.1 |  | Meningitis |
| B06.0 |  | Other nervous system infections |
| B26.1 |  | Meningitis |
| B26.2 |  | Encephalitis |
| B37.5 |  | Meningitis |
| B38.4 |  | Meningitis |
| B43.1 |  | Other nervous system infections |
| B45.1 |  | Other nervous system infections |
| B50.0 |  | Other nervous system infections |
| B58.2 |  | Other nervous system infections |
| B69.0 |  | Other nervous system infections |
| B90.0 |  | Other nervous system infections |
| B91 | Sequelae of poliomyelitis | Other nervous system infections |
| B94.1 | Sequelae of viral encephalitis | Encephalitis |
| G00 | Bacterial meningitis, not elsewhere classified | Meningitis |
| G01 | Meningitis in bacterial diseases classified elsewhere | Meningitis |
| G02.0 | Meningitis in viral diseases classified elsewhere | Meningitis |
| G02.1 | Meningitis in mycoses | Meningitis |
| G02.8 | Meningitis in other specified infectious and parasitic diseases classified elsewhere | Meningitis |
| G04.2 | Bacterial meningoencephalitis and meningomyelitis, not elsewhere classified | Other nervous system infections |
| G04.8 | Other encephalitis, myelitis and encephalomyelitis | Other nervous system infections |
| G04.9 | Encephalitis, myelitis and encephalomyelitis, unspecified | Other nervous system infections |
| G05.0 | Encephalitis, myelitis and encephalomyelitis in bacterial diseases classified elsewhere | Other nervous system infections |
| G05.1 | Encephalitis, myelitis and encephalomyelitis in viral diseases classified elsewhere | Other nervous system infections |
| G05.2 | Encephalitis, myelitis and encephalomyelitis in other infectious and parasitic diseases classified elsewhere | Other nervous system infections |
| G06 | Intracranial and intraspinal abscess and granuloma | Other nervous system infections |
| G07 | Intracranial and intraspinal abscess and granuloma in diseases classified elsewhere | Other nervous system infections |
| G08 | Intracranial and intraspinal phlebitis and thrombophlebitis | Other nervous system infections |
| M89.6 | Osteopathy after poliomyelitis | Other nervous system infections |

##### Ocular infections

| **ICD-10 code** | **Description** | **Infection type** |
| --- | --- | --- |
| A18.5 |  | Eye infections |
| A21.1 |  | Eye infections |
| A54.3 |  | Eye infections |
| A71 | Trachoma | Eye infections |
| A74.0 |  | Eye infections |
| B00.5 |  | Eye infections |
| B02.3 |  | Eye infections |
| B30 | Viral conjunctivitis | Eye infections |
| B58.0 |  | Eye infections |
| B69.1 |  | Eye infections |
| B87.2 |  | Eye infections |
| B94.0 | Sequelae of trachoma | Eye infections |
| H00.0 |  | Eye infections |
| H10 | Conjunctivitis | Eye infections |
| H13.1 | Conjunctivitis in infectious and parasitic diseases classified elsewhere | Eye infections |
| H19.1 | Herpesviral keratitis and keratoconjunctivitis | Eye infections |
| H19.2 | Keratitis and keratoconjunctivitis in other infectious and parasitic diseases classified elsewhere | Eye infections |
| P39.1 |  | Eye infections |

##### Sepsis

| **ICD-10 code** | **Description** | **Infection type** |
| --- | --- | --- |
| A02.1 |  | Septicaemia |
| A20.7 |  | Septicaemia |
| A22.7 |  | Septicaemia |
| A26.7 |  | Septicaemia |
| A32.7 |  | Septicaemia |
| A39.1 |  | Septicaemia |
| A39.2 |  | Septicaemia |
| A39.3 |  | Septicaemia |
| A39.4 |  | Septicaemia |
| A40 | Streptococcal sepsis | Septicaemia |
| A41 |  | Septicaemia |
| A42.7 |  | Septicaemia |
| B37.7 |  | Septicaemia |
| P36 | Bacterial sepsis of newborn | Septicaemia |

##### Skin and soft tissue infections

| **ICD-10 code** | **Description** | **Infection type** |
| --- | --- | --- |
| A06.7 |  | Infection of skin and subcutaneous tissues |
| A18.4 |  | Infection of skin and subcutaneous tissues |
| A20.1 |  | Infection of skin and subcutaneous tissues |
| A21.0 |  | Infection of skin and subcutaneous tissues |
| A22.0 |  | Infection of skin and subcutaneous tissues |
| A26.0 |  | Infection of skin and subcutaneous tissues |
| A31.1 |  | Infection of skin and subcutaneous tissues |
| A32.0 |  | Infection of skin and subcutaneous tissues |
| A36.3 |  | Infection of skin and subcutaneous tissues |
| A43.1 |  | Infection of skin and subcutaneous tissues |
| A46 | Erysipelas | Infection of skin and subcutaneous tissues |
| A51.3 |  | Infection of skin and subcutaneous tissues |
| B00.0 |  | Infection of skin and subcutaneous tissues |
| B00.1 |  | Infection of skin and subcutaneous tissues |
| B07 | Viral warts | Infection of skin and subcutaneous tissues |
| B08 | Other viral infections characterized by skin and mucous membrane lesions, not elsewhere classified | Infection of skin and subcutaneous tissues |
| B09 | Unspecified viral infection characterized by skin and mucous membrane lesions | Infection of skin and subcutaneous tissues |
| B35 | Dermatophytosis | Infection of skin and subcutaneous tissues |
| B36 | Other superficial mycoses | Infection of skin and subcutaneous tissues |
| B37.2 |  | Infection of skin and subcutaneous tissues |
| B38.3 |  | Infection of skin and subcutaneous tissues |
| B40.3 |  | Infection of skin and subcutaneous tissues |
| B42.1 |  | Infection of skin and subcutaneous tissues |
| B43.0 |  | Infection of skin and subcutaneous tissues |
| B43.2 |  | Infection of skin and subcutaneous tissues |
| B45.2 |  | Infection of skin and subcutaneous tissues |
| B46.3 |  | Infection of skin and subcutaneous tissues |
| B55.1 |  | Infection of skin and subcutaneous tissues |
| B78.1 |  | Infection of skin and subcutaneous tissues |
| B85 | Pediculosis and phthiriasis | Infection of skin and subcutaneous tissues |
| B86 | Scabies | Infection of skin and subcutaneous tissues |
| B87.0 |  | Infection of skin and subcutaneous tissues |
| B87.1 |  | Infection of skin and subcutaneous tissues |
| B88 | Other infestations | Infection of skin and subcutaneous tissues |
| L00 | Staphylococcal scalded skin syndrome | Infection of skin and subcutaneous tissues |
| L01 | Impetigo | Infection of skin and subcutaneous tissues |
| L02 | Cutaneous abscess, furuncle and carbuncle | Infection of skin and subcutaneous tissues |
| L03 | Cellulitis | Infection of skin and subcutaneous tissues |
| L05.0 | Pilonidal cyst with abscess | Infection of skin and subcutaneous tissues |
| L08 |  | Infection of skin and subcutaneous tissues |
| L30.3 | Infective dermatitis | Infection of skin and subcutaneous tissues |
| P38 |  | Infection of skin and subcutaneous tissues |
| P39.4 |  | Infection of skin and subcutaneous tissues |

##### Upper respiratory and ENT infections

| **ICD-10 code** | **Description** | **Infection type** |
| --- | --- | --- |
| A18.6 |  | Ear and Upper Respiratory Tract Infections |
| A36.0 |  | Ear and Upper Respiratory Tract Infections |
| A36.1 |  | Ear and Upper Respiratory Tract Infections |
| A36.2 |  | Ear and Upper Respiratory Tract Infections |
| A54.5 |  | Ear and Upper Respiratory Tract Infections |
| A56.4 |  | Ear and Upper Respiratory Tract Infections |
| B05.3 |  | Ear and Upper Respiratory Tract Infections |
| B27 | Infectious mononucleosis | Ear and Upper Respiratory Tract Infections |
| B44.2 | Tonsillar aspergillosis | Ear and Upper Respiratory Tract Infections |
| B87.3 |  | Ear and Upper Respiratory Tract Infections |
| B87.4 |  | Ear and Upper Respiratory Tract Infections |
| H60 | Otitis externa | Ear and Upper Respiratory Tract Infections |
| H62.0 | Otitis externa in bacterial diseases classified elsewhere | Ear and Upper Respiratory Tract Infections |
| H62.1 | Otitis externa in viral diseases classified elsewhere | Ear and Upper Respiratory Tract Infections |
| H62.2 | Otitis externa in mycoses | Ear and Upper Respiratory Tract Infections |
| H62.3 | Otitis externa in other infectious and parasitic diseases classified elsewhere | Ear and Upper Respiratory Tract Infections |
| H62.4 | Otitis externa in other diseases classified elsewhere | Ear and Upper Respiratory Tract Infections |
| H65 | Nonsuppurative otitis media | Ear and Upper Respiratory Tract Infections |
| H66 | Suppurative and unspecified otitis media | Ear and Upper Respiratory Tract Infections |
| H67 |  | Ear and Upper Respiratory Tract Infections |
| H70 | Mastoiditis and related conditions | Ear and Upper Respiratory Tract Infections |
| H73.0 | Acute myringitis | Ear and Upper Respiratory Tract Infections |
| H73.1 | Chronic myringitis | Ear and Upper Respiratory Tract Infections |
| H75.0 | Mastoiditis in infectious and parasitic diseases classified elsewhere | Ear and Upper Respiratory Tract Infections |
| J00 | Acute nasopharyngitis [common cold] | Ear and Upper Respiratory Tract Infections |
| J01 | Acute sinusitis | Ear and Upper Respiratory Tract Infections |
| J02 |  | Ear and Upper Respiratory Tract Infections |
| J03 |  | Ear and Upper Respiratory Tract Infections |
| J04 | Acute laryngitis and tracheitis | Ear and Upper Respiratory Tract Infections |
| J05 | Acute obstructive laryngitis [croup] and epiglottitis | Ear and Upper Respiratory Tract Infections |
| J06 | Acute upper respiratory infections of multiple and unspecified sites | Ear and Upper Respiratory Tract Infections |
| J34.0 | Abscess, furuncle and carbuncle of nose | Ear and Upper Respiratory Tract Infections |
| J36 | Peritonsillar abscess | Ear and Upper Respiratory Tract Infections |
| J37 | Chronic laryngitis and laryngotracheitis | Ear and Upper Respiratory Tract Infections |
| J39.0 | Retropharyngeal and parapharyngeal abscess | Ear and Upper Respiratory Tract Infections |
| J39.1 | Other abscess of pharynx | Ear and Upper Respiratory Tract Infections |

##### Urinary infections

| **ICD-10 code** | **Description** | **Infection type** |
| --- | --- | --- |
| N08.0 | Glomerular disorders in infectious and parasitic diseases classified elsewhere | Urinary Tract Infections |
| N10 | Acute tubulo-interstitial nephritis | Urinary Tract Infections |
| N13.6 | Pyonephrosis | Urinary Tract Infections |
| N15.1 | Renal and perinephric abscess | Urinary Tract Infections |
| N16.0 | Renal tubulo-interstitial disorders in infectious and parasitic diseases classified elsewhere | Urinary Tract Infections |
| N29.0 | Late syphilis of kidney | Urinary Tract Infections |
| N29.1 | Other disorders of kidney and ureter in infectious and parasitic diseases classified elsewhere | Urinary Tract Infections |
| N30.0 | Acute cystitis | Urinary Tract Infections |
| N30.9 | Cystitis, unspecified | Urinary Tract Infections |
| N33.0 |  | Urinary Tract Infections |
| N34 | Urethritis and urethral syndrome | Urinary Tract Infections |
| N37.0 |  | Urinary Tract Infections |
| N39.0 | Urinary tract infection, site not specified | Urinary Tract Infections |

#### Comorbidities

ICD-10 codes used to define comorbidity categories in the study cohort. Each table corresponds to one comorbidity class; sub-categories within each class are shown in the Category column.

##### Cardiac

| **ICD-10 code** | **Category** | **Description** |
| --- | --- | --- |
| I05X | Chronic cardiac comorbidities | Rheumatic mitral valve diseases |
| I050 | Chronic cardiac comorbidities | Mitral stenosis |
| I051 | Chronic cardiac comorbidities | Rheumatic mitral insufficiency |
| I052 | Chronic cardiac comorbidities | Mitral stenosis with insufficiency |
| I058 | Chronic cardiac comorbidities | Other mitral valve diseases |
| I059 | Chronic cardiac comorbidities | Mitral valve disease, unspecified |
| I06X | Chronic cardiac comorbidities | Rheumatic aortic valve diseases |
| I060 | Chronic cardiac comorbidities | Rheumatic aortic stenosis |
| I061 | Chronic cardiac comorbidities | Rheumatic aortic insufficiency |
| I062 | Chronic cardiac comorbidities | Rheumatic aortic stenosis with insufficiency |
| I068 | Chronic cardiac comorbidities | Other rheumatic aortic valve diseases |
| I069 | Chronic cardiac comorbidities | Rheumatic aortic valve disease, unspecified |
| I07X | Chronic cardiac comorbidities | Rheumatic tricuspid valve diseases |
| I070 | Chronic cardiac comorbidities | Tricuspid stenosis |
| I071 | Chronic cardiac comorbidities | Tricuspid insufficiency |
| I072 | Chronic cardiac comorbidities | Tricuspid stenosis with insufficiency |
| I078 | Chronic cardiac comorbidities | Other tricuspid valve diseases |
| I079 | Chronic cardiac comorbidities | Tricuspid valve disease, unspecified |
| I08X | Chronic cardiac comorbidities | Multiple valve diseases |
| I080 | Chronic cardiac comorbidities | Disorders of both mitral and aortic valves |
| I081 | Chronic cardiac comorbidities | Disorders of both mitral and tricuspid valves |
| I082 | Chronic cardiac comorbidities | Disorders of both aortic and tricuspid valves |
| I083 | Chronic cardiac comorbidities | Combined disorders of mitral, aortic and tricuspid valves |
| I088 | Chronic cardiac comorbidities | Other multiple valve diseases |
| I089 | Chronic cardiac comorbidities | Multiple valve disease, unspecified |
| I09X | Chronic cardiac comorbidities | Other rheumatic heart diseases |
| I090 | Chronic cardiac comorbidities | Rheumatic myocarditis |
| I091 | Chronic cardiac comorbidities | Rheumatic diseases of endocardium, valve unspecified |
| I092 | Chronic cardiac comorbidities | Chronic rheumatic pericarditis |
| I098 | Chronic cardiac comorbidities | Other specified rheumatic heart diseases |
| I099 | Chronic cardiac comorbidities | Rheumatic heart disease, unspecified |
| I11X | Chronic cardiac comorbidities | Hypertensive heart disease |
| I110 | Chronic cardiac comorbidities | Hypertensive heart disease with (congestive) heart failure |
| I119 | Chronic cardiac comorbidities | Hypertensive heart disease without (congestive) heart failure |
| I13X | Chronic cardiac comorbidities | Hypertensive heart and renal disease |
| I130 | Chronic cardiac comorbidities | Hypertensive heart and renal disease with (congestive) heart failure |
| I131 | Chronic cardiac comorbidities | Hypertensive heart and renal disease with renal failure |
| I132 | Chronic cardiac comorbidities | Hypertensive heart and renal disease with both (congestive) heart failure and renal failure |
| I139 | Chronic cardiac comorbidities | Hypertensive heart and renal disease, unspecified |
| I25X | Chronic cardiac comorbidities | Chronic ischaemic heart disease |
| I250 | Chronic cardiac comorbidities | Atherosclerotic cardiovascular disease, so described |
| I251 | Chronic cardiac comorbidities | Atherosclerotic heart disease |
| I252 | Chronic cardiac comorbidities | Old myocardial infarction |
| I253 | Chronic cardiac comorbidities | Aneurysm of heart |
| I254 | Chronic cardiac comorbidities | Coronary artery aneurysm and dissection |
| I255 | Chronic cardiac comorbidities | Ischaemic cardiomyopathy |
| I256 | Chronic cardiac comorbidities | Silent myocardial ischaemia |
| I258 | Chronic cardiac comorbidities | Other forms of chronic ischaemic heart disease |
| I259 | Chronic cardiac comorbidities | Chronic ischaemic heart disease, unspecified |
| I34X | Chronic cardiac comorbidities | Nonrheumatic mitral valve disorders |
| I340 | Chronic cardiac comorbidities | Mitral (valve) insufficiency |
| I341 | Chronic cardiac comorbidities | Mitral (valve) prolapse |
| I342 | Chronic cardiac comorbidities | Nonrheumatic mitral (valve) stenosis |
| I348 | Chronic cardiac comorbidities | Other nonrheumatic mitral valve disorders |
| I349 | Chronic cardiac comorbidities | Nonrheumatic mitral valve disorder, unspecified |
| I35X | Chronic cardiac comorbidities | Nonrheumatic aortic valve disorders |
| I350 | Chronic cardiac comorbidities | Aortic (valve) stenosis |
| I351 | Chronic cardiac comorbidities | Aortic (valve) insufficiency |
| I352 | Chronic cardiac comorbidities | Aortic (valve) stenosis with insufficiency |
| I358 | Chronic cardiac comorbidities | Other aortic valve disorders |
| I359 | Chronic cardiac comorbidities | Aortic valve disorder, unspecified |
| I36X | Chronic cardiac comorbidities | Nonrheumatic tricuspid valve disorders |
| I360 | Chronic cardiac comorbidities | Nonrheumatic tricuspid (valve) stenosis |
| I361 | Chronic cardiac comorbidities | Nonrheumatic tricuspid (valve) insufficiency |
| I362 | Chronic cardiac comorbidities | Nonrheumatic tricuspid (valve) stenosis with insufficiency |
| I368 | Chronic cardiac comorbidities | Other nonrheumatic tricuspid valve disorders |
| I369 | Chronic cardiac comorbidities | Nonrheumatic tricuspid valve disorder, unspecified |
| I37X | Chronic cardiac comorbidities | Pulmonary valve disorders |
| I370 | Chronic cardiac comorbidities | Pulmonary valve stenosis |
| I371 | Chronic cardiac comorbidities | Pulmonary valve insufficiency |
| I372 | Chronic cardiac comorbidities | Pulmonary valve stenosis with insufficiency |
| I378 | Chronic cardiac comorbidities | Other pulmonary valve disorders |
| I379 | Chronic cardiac comorbidities | Pulmonary valve disorder, unspecified |
| I42X | Chronic cardiac comorbidities | Cardiomyopathy |
| I420 | Chronic cardiac comorbidities | Dilated cardiomyopathy |
| I421 | Chronic cardiac comorbidities | Obstructive hypertrophic cardiomyopathy |
| I422 | Chronic cardiac comorbidities | Other hypertrophic cardiomyopathy |
| I423 | Chronic cardiac comorbidities | Endomyocardial (eosinophilic) disease |
| I424 | Chronic cardiac comorbidities | Endocardial fibroelastosis |
| I425 | Chronic cardiac comorbidities | Other restrictive cardiomyopathy |
| I426 | Chronic cardiac comorbidities | Alcoholic cardiomyopathy |
| I427 | Chronic cardiac comorbidities | Cardiomyopathy due to drugs and other external agents |
| I428 | Chronic cardiac comorbidities | Other cardiomyopathies |
| I429 | Chronic cardiac comorbidities | Cardiomyopathy, unspecified |
| I43X | Chronic cardiac comorbidities | Cardiomyopathy in diseases classified elsewhere |
| I430 | Chronic cardiac comorbidities | Cardiomyopathy in infectious and parasitic diseases classified elsewhere |
| I431 | Chronic cardiac comorbidities | Cardiomyopathy in metabolic diseases |
| I432 | Chronic cardiac comorbidities | Cardiomyopathy in nutritional diseases |
| I438 | Chronic cardiac comorbidities | Cardiomyopathy in other diseases classified elsewhere |
| I44X | Chronic cardiac comorbidities | Atrioventricular and left bundle-branch block |
| I440 | Chronic cardiac comorbidities | Atrioventricular block, first degree |
| I441 | Chronic cardiac comorbidities | Atrioventricular block, second degree |
| I442 | Chronic cardiac comorbidities | Atrioventricular block, complete |
| I443 | Chronic cardiac comorbidities | Other and unspecified atrioventricular block |
| I444 | Chronic cardiac comorbidities | Left anterior fascicular block |
| I445 | Chronic cardiac comorbidities | Left posterior fascicular block |
| I446 | Chronic cardiac comorbidities | Other and unspecified fascicular block |
| I447 | Chronic cardiac comorbidities | Left bundle-branch block, unspecified |
| I45X | Chronic cardiac comorbidities | Other conduction disorders |
| I450 | Chronic cardiac comorbidities | Right fascicular block |
| I451 | Chronic cardiac comorbidities | Other and unspecified right bundle-branch block |
| I452 | Chronic cardiac comorbidities | Bifascicular block |
| I453 | Chronic cardiac comorbidities | Trifascicular block |
| I454 | Chronic cardiac comorbidities | Nonspecific intraventricular block |
| I455 | Chronic cardiac comorbidities | Other specified heart block |
| I456 | Chronic cardiac comorbidities | Pre-excitation syndrome |
| I458 | Chronic cardiac comorbidities | Other specified conduction disorders |
| I459 | Chronic cardiac comorbidities | Conduction disorder, unspecified |
| I47X | Chronic cardiac comorbidities | Paroxysmal tachycardia |
| I470 | Chronic cardiac comorbidities | Re-entry ventricular arrhythmia |
| I471 | Chronic cardiac comorbidities | Supraventricular tachycardia |
| I472 | Chronic cardiac comorbidities | Ventricular tachycardia |
| I479 | Chronic cardiac comorbidities | Paroxysmal tachycardia, unspecified |
| I48X | Chronic cardiac comorbidities | Atrial fibrillation and flutter |
| I48X | Chronic cardiac comorbidities | Atrial fibrillation and flutter |
| I480 | Chronic cardiac comorbidities | Paroxysmal atrial fibrillation |
| I481 | Chronic cardiac comorbidities | Persistent atrial fibrillation |
| I482 | Chronic cardiac comorbidities | Chronic atrial fibrillation |
| I483 | Chronic cardiac comorbidities | Typical atrial flutter |
| I484 | Chronic cardiac comorbidities | Atypical atrial flutter |
| I489 | Chronic cardiac comorbidities | Atrial fibrillation and atrial flutter, unspecified |
| I50X | Chronic cardiac comorbidities | Heart failure |
| I500 | Chronic cardiac comorbidities | Congestive heart failure |
| I501 | Chronic cardiac comorbidities | Left ventricular failure |
| I509 | Chronic cardiac comorbidities | Heart failure, unspecified |
| I51X | Chronic cardiac comorbidities | Complications and ill-defined descriptions of heart disease |
| I510 | Chronic cardiac comorbidities | Cardiac septal defect, acquired |
| I511 | Chronic cardiac comorbidities | Rupture of chordae tendineae, not elsewhere classified |
| I512 | Chronic cardiac comorbidities | Rupture of papillary muscle, not elsewhere classified |
| I513 | Chronic cardiac comorbidities | Intracardiac thrombosis, not elsewhere classified |
| I514 | Chronic cardiac comorbidities | Myocarditis, unspecified |
| I515 | Chronic cardiac comorbidities | Myocardial degeneration |
| I516 | Chronic cardiac comorbidities | Cardiovascular disease, unspecified |
| I517 | Chronic cardiac comorbidities | Cardiomegaly |
| I518 | Chronic cardiac comorbidities | Other ill-defined heart diseases |
| I519 | Chronic cardiac comorbidities | Heart disease, unspecified |

##### Respiratory

| **ICD-10 code** | **Category** | **Description** |
| --- | --- | --- |
| I270 | Chronic respiratory comorbidities | Primary pulmonary hypertension |
| J43X | Chronic respiratory comorbidities | Emphysema |
| J430 | Chronic respiratory comorbidities | MacLeod syndrome |
| J431 | Chronic respiratory comorbidities | Panlobular emphysema |
| J432 | Chronic respiratory comorbidities | Centrilobular emphysema |
| J438 | Chronic respiratory comorbidities | Other emphysema |
| J439 | Chronic respiratory comorbidities | Emphysema, unspecified |
| J44X | Chronic respiratory comorbidities | Other chronic obstructive pulmonary disease |
| J440 | Chronic respiratory comorbidities | Chronic obstructive pulmonary disease with acute lower respiratory infection |
| J441 | Chronic respiratory comorbidities | Chronic obstructive pulmonary disease with acute exacerbation, unspecified |
| J448 | Chronic respiratory comorbidities | Other specified chronic obstructive pulmonary disease |
| J449 | Chronic respiratory comorbidities | Chronic obstructive pulmonary disease, unspecified |
| J45X | Chronic respiratory comorbidities | Asthma |
| J450 | Chronic respiratory comorbidities | Predominantly allergic asthma |
| J451 | Chronic respiratory comorbidities | Nonallergic asthma |
| J458 | Chronic respiratory comorbidities | Mixed asthma |
| J459 | Chronic respiratory comorbidities | Asthma, unspecified |
| J47X | Chronic respiratory comorbidities | Bronchiectasis |
| J60X | Chronic respiratory comorbidities | Coalworker pneumoconiosis |
| J61X | Chronic respiratory comorbidities | Pneumoconiosis due to asbestos and other mineral fibres |
| J62X | Chronic respiratory comorbidities | Pneumoconiosis due to dust containing silica |
| J620 | Chronic respiratory comorbidities | Pneumoconiosis due to talc dust |
| J628 | Chronic respiratory comorbidities | Pneumoconiosis due to other dust containing silica |
| J63X | Chronic respiratory comorbidities | Pneumoconiosis due to other inorganic dusts |
| J630 | Chronic respiratory comorbidities | Aluminosis (of lung) |
| J631 | Chronic respiratory comorbidities | Bauxite fibrosis (of lung) |
| J632 | Chronic respiratory comorbidities | Berylliosis |
| J633 | Chronic respiratory comorbidities | Graphite fibrosis (of lung) |
| J634 | Chronic respiratory comorbidities | Siderosis |
| J635 | Chronic respiratory comorbidities | Stannosis |
| J638 | Chronic respiratory comorbidities | Pneumoconiosis due to other specified inorganic dusts |
| J64X | Chronic respiratory comorbidities | Unspecified pneumoconiosis |
| J65X | Chronic respiratory comorbidities | Pneumoconiosis associated with tuberculosis |
| J66X | Chronic respiratory comorbidities | Airway disease due to specific organic dust |
| J660 | Chronic respiratory comorbidities | Byssinosis |
| J661 | Chronic respiratory comorbidities | Flax-dresser disease |
| J662 | Chronic respiratory comorbidities | Cannabinosis |
| J668 | Chronic respiratory comorbidities | Airway disease due to other specific organic dusts |
| J67X | Chronic respiratory comorbidities | Hypersensitivity pneumonitis due to organic dust |
| J670 | Chronic respiratory comorbidities | Farmer lung |
| J671 | Chronic respiratory comorbidities | Bagassosis |
| J672 | Chronic respiratory comorbidities | Bird fancier lung |
| J673 | Chronic respiratory comorbidities | Suberosis |
| J674 | Chronic respiratory comorbidities | Maltworker lung |
| J675 | Chronic respiratory comorbidities | Mushroom-worker lung |
| J676 | Chronic respiratory comorbidities | Maple-bark-stripper lung |
| J677 | Chronic respiratory comorbidities | Air-conditioner and humidifier lung |
| J678 | Chronic respiratory comorbidities | Hypersensitivity pneumonitis due to other organic dusts |
| J679 | Chronic respiratory comorbidities | Hypersensitivity pneumonitis due to unspecified organic dust |
| J684 | Chronic respiratory comorbidities | Chronic respiratory conditions due to chemicals, gases, fumes and vapours |
| J701 | Chronic respiratory comorbidities | Chronic and other pulmonary manifestations due to radiation |
| J703 | Chronic respiratory comorbidities | Chronic drug-induced interstitial lung disorders |
| J84X | Chronic respiratory comorbidities | Other interstitial pulmonary diseases |
| J840 | Chronic respiratory comorbidities | Alveolar and parietoalveolar conditions |
| J841 | Chronic respiratory comorbidities | Other interstitial pulmonary diseases with fibrosis |
| J848 | Chronic respiratory comorbidities | Other specified interstitial pulmonary diseases |
| J849 | Chronic respiratory comorbidities | Interstitial pulmonary disease, unspecified |
| J961 | Chronic respiratory comorbidities | Chronic respiratory failure |
| J99X | Chronic respiratory comorbidities | Respiratory disorders in diseases classified elsewhere |
| J990 | Chronic respiratory comorbidities | Rheumatoid lung disease (M05.1+) |
| J991 | Chronic respiratory comorbidities | Respiratory disorders in other diffuse connective tissue disorders |
| J998 | Chronic respiratory comorbidities | Respiratory disorders in other diseases classified elsewhere |

##### Liver

| **ICD-10 code** | **Category** | **Description** |
| --- | --- | --- |
| K702 | Chronic liver comorbidities | Alcoholic fibrosis and sclerosis of liver |
| K703 | Chronic liver comorbidities | Alcoholic cirrhosis of liver |
| K704 | Chronic liver comorbidities | Alcoholic hepatic failure |
| K717 | Chronic liver comorbidities | Toxic liver disease with fibrosis and cirrhosis of liver |
| K721 | Chronic liver comorbidities | Chronic hepatic failure |
| K74X | Chronic liver comorbidities | Fibrosis and cirrhosis of liver |
| K740 | Chronic liver comorbidities | Hepatic fibrosis |
| K741 | Chronic liver comorbidities | Hepatic sclerosis |
| K742 | Chronic liver comorbidities | Hepatic fibrosis with hepatic sclerosis |
| K743 | Chronic liver comorbidities | Primary biliary cirrhosis |
| K744 | Chronic liver comorbidities | Secondary biliary cirrhosis |
| K745 | Chronic liver comorbidities | Biliary cirrhosis, unspecified |
| K746 | Chronic liver comorbidities | Other and unspecified cirrhosis of liver |
| K766 | Chronic liver comorbidities | Portal hypertension |
| K767 | Chronic liver comorbidities | Hepatorenal syndrome |

##### Renal

| **ICD-10 code** | **Category** | **Description** |
| --- | --- | --- |
| I12X | Chronic renal comorbidities | Hypertensive renal disease |
| I120 | Chronic renal comorbidities | Hypertensive renal disease with renal failure |
| I129 | Chronic renal comorbidities | Hypertensive renal disease without renal failure |
| I13X | Chronic renal comorbidities | Hypertensive heart and renal disease |
| I130 | Chronic renal comorbidities | Hypertensive heart and renal disease with (congestive) heart failure |
| I131 | Chronic renal comorbidities | Hypertensive heart and renal disease with renal failure |
| I132 | Chronic renal comorbidities | Hypertensive heart and renal disease with both (congestive) heart failure and renal failure |
| I139 | Chronic renal comorbidities | Hypertensive heart and renal disease, unspecified |
| I151 | Chronic renal comorbidities | Hypertension secondary to other renal disorders |
| N183 | Chronic renal comorbidities | Chronic kidney disease, stage 3 |
| N184 | Chronic renal comorbidities | Chronic kidney disease, stage 4 |
| N185 | Chronic renal comorbidities | Chronic kidney disease, stage 5 |

##### Neurological

| **ICD-10 code** | **Category** | **Description** |
| --- | --- | --- |
| I69X | Chronic neurological comorbidities | Sequelae of cerebrovascular disease |
| I690 | Chronic neurological comorbidities | Sequelae of subarachnoid haemorrhage |
| I691 | Chronic neurological comorbidities | Sequelae of intracerebral haemorrhage |
| I692 | Chronic neurological comorbidities | Sequelae of other nontraumatic intracranial haemorrhage |
| I693 | Chronic neurological comorbidities | Sequelae of cerebral infarction |
| I694 | Chronic neurological comorbidities | Sequelae of stroke, not specified as haemorrhage or infarction |
| I698 | Chronic neurological comorbidities | Sequelae of other and unspecified cerebrovascular diseases |
| G10X | Chronic neurological comorbidities | Huntington disease |
| G11X | Chronic neurological comorbidities | Hereditary ataxia |
| G110 | Chronic neurological comorbidities | Congenital nonprogressive ataxia |
| G111 | Chronic neurological comorbidities | Early-onset cerebellar ataxia |
| G112 | Chronic neurological comorbidities | Late-onset cerebellar ataxia |
| G113 | Chronic neurological comorbidities | Cerebellar ataxia with defective DNA repair |
| G114 | Chronic neurological comorbidities | Hereditary spastic paraplegia |
| G118 | Chronic neurological comorbidities | Other hereditary ataxias |
| G119 | Chronic neurological comorbidities | Hereditary ataxia, unspecified |
| G12X | Chronic neurological comorbidities | Spinal muscular atrophy and related syndromes |
| G120 | Chronic neurological comorbidities | Infantile spinal muscular atrophy, type i (werdnig-hoffman) |
| G121 | Chronic neurological comorbidities | Other inherited spinal muscular atrophy |
| G122 | Chronic neurological comorbidities | Motor neuron disease |
| G128 | Chronic neurological comorbidities | Other spinal muscular atrophies and related syndromes |
| G129 | Chronic neurological comorbidities | Spinal muscular atrophy, unspecified |
| G13X | Chronic neurological comorbidities | Systemic atrophies primarily affecting central nervous system in diseases classified elsewhere |
| G130 | Chronic neurological comorbidities | Paraneoplastic neuromyopathy and neuropathy |
| G131 | Chronic neurological comorbidities | Other systemic atrophy primarily affecting central nervous system in neoplastic disease |
| G132 | Chronic neurological comorbidities | Systemic atrophy primarily affecting central nervous system in myxoedema |
| G138 | Chronic neurological comorbidities | Systemic atrophy primarily affecting central nervous system in other diseases classified elsewhere |
| G20X | Chronic neurological comorbidities | Parkinson disease |
| G213 | Chronic neurological comorbidities | Postencephalitic parkinsonism |
| G214 | Chronic neurological comorbidities | Vascular parkinsonism |
| G23X | Chronic neurological comorbidities | Other degenerative diseases of basal ganglia |
| G230 | Chronic neurological comorbidities | Hallervorden-Spatz disease |
| G231 | Chronic neurological comorbidities | Progressive supranuclear ophthalmoplegia (steele-Richardson-olszewski) |
| G232 | Chronic neurological comorbidities | Multiple system atrophy, parkinsonian type (msa-p) |
| G233 | Chronic neurological comorbidities | Multiple system atrophy, cerebellar type (msa-c) |
| G238 | Chronic neurological comorbidities | Other specified degenerative diseases of basal ganglia |
| G239 | Chronic neurological comorbidities | Degenerative disease of basal ganglia, unspecified |
| G35X | Chronic neurological comorbidities | Multiple sclerosis |
| G360 | Chronic neurological comorbidities | Neuromyelitis optica (devic) |
| G37X | Chronic neurological comorbidities | Other demyelinating diseases of central nervous system |
| G370 | Chronic neurological comorbidities | Diffuse sclerosis |
| G371 | Chronic neurological comorbidities | Central demyelination of corpus callosum |
| G372 | Chronic neurological comorbidities | Central pontine myelinolysis |
| G373 | Chronic neurological comorbidities | Acute transverse myelitis in demyelinating disease of central nervous system |
| G374 | Chronic neurological comorbidities | Subacute necrotizing myelitis |
| G375 | Chronic neurological comorbidities | Concentric sclerosis (baló) |
| G378 | Chronic neurological comorbidities | Other specified demyelinating diseases of central nervous system |
| G379 | Chronic neurological comorbidities | Demyelinating disease of central nervous system, unspecified |
| G40X | Chronic neurological comorbidities | Epilepsy |
| G400 | Chronic neurological comorbidities | Localization-related (focal)(partial) idiopathic epilepsy and epileptic syndromes with seizures of localized onset |
| G401 | Chronic neurological comorbidities | Localization-related (focal)(partial) symptomatic epilepsy and epileptic syndromes with simple partial seizures |
| G402 | Chronic neurological comorbidities | Localization-related (focal)(partial) symptomatic epilepsy and epileptic syndromes with complex partial seizures |
| G403 | Chronic neurological comorbidities | Generalized idiopathic epilepsy and epileptic syndromes |
| G404 | Chronic neurological comorbidities | Other generalized epilepsy and epileptic syndromes |
| G405 | Chronic neurological comorbidities | Special epileptic syndromes |
| G406 | Chronic neurological comorbidities | Grand mal seizures, unspecified (with or without petit mal) |
| G407 | Chronic neurological comorbidities | Petit mal, unspecified, without grand mal seizures |
| G408 | Chronic neurological comorbidities | Other epilepsy |
| G409 | Chronic neurological comorbidities | Epilepsy, unspecified |
| G60X | Chronic neurological comorbidities | Hereditary and idiopathic neuropathy |
| G600 | Chronic neurological comorbidities | Hereditary motor and sensory neuropathy |
| G601 | Chronic neurological comorbidities | Refsum disease |
| G602 | Chronic neurological comorbidities | Neuropathy in association with hereditary ataxia |
| G603 | Chronic neurological comorbidities | Idiopathic progressive neuropathy |
| G608 | Chronic neurological comorbidities | Other hereditary and idiopathic neuropathies |
| G609 | Chronic neurological comorbidities | Hereditary and idiopathic neuropathy, unspecified |
| G70X | Chronic neurological comorbidities | Myasthenia gravis and other myoneural disorders |
| G700 | Chronic neurological comorbidities | Myasthenia gravis |
| G701 | Chronic neurological comorbidities | Toxic myoneural disorders |
| G702 | Chronic neurological comorbidities | Congenital and developmental myasthenia |
| G708 | Chronic neurological comorbidities | Other specified myoneural disorders |
| G709 | Chronic neurological comorbidities | Myoneural disorder, unspecified |
| G71X | Chronic neurological comorbidities | Primary disorders of muscles |
| G710 | Chronic neurological comorbidities | Muscular dystrophy |
| G711 | Chronic neurological comorbidities | Myotonic disorders |
| G712 | Chronic neurological comorbidities | Congenital myopathies |
| G713 | Chronic neurological comorbidities | Mitochondrial myopathy, not elsewhere classified |
| G718 | Chronic neurological comorbidities | Other primary disorders of muscles |
| G719 | Chronic neurological comorbidities | Primary disorder of muscle, unspecified |
| G80X | Chronic neurological comorbidities | Cerebral palsy |
| G800 | Chronic neurological comorbidities | Spastic quadriplegic cerebral palsy |
| G801 | Chronic neurological comorbidities | Spastic diplegic cerebral palsy |
| G802 | Chronic neurological comorbidities | Spastic hemiplegic cerebral palsy |
| G803 | Chronic neurological comorbidities | Dyskinetic cerebral palsy |
| G804 | Chronic neurological comorbidities | Ataxic cerebral palsy |
| G808 | Chronic neurological comorbidities | Other cerebral palsy |
| G809 | Chronic neurological comorbidities | Cerebral palsy, unspecified |
| G81X | Chronic neurological comorbidities | Hemiplegia |
| G810 | Chronic neurological comorbidities | Flaccid hemiplegia |
| G811 | Chronic neurological comorbidities | Spastic hemiplegia |
| G819 | Chronic neurological comorbidities | Hemiplegia, unspecified |
| G82X | Chronic neurological comorbidities | Paraplegia and tetraplegia |
| G820 | Chronic neurological comorbidities | Flaccid paraplegia |
| G821 | Chronic neurological comorbidities | Spastic paraplegia |
| G822 | Chronic neurological comorbidities | Paraplegia, unspecified |
| G823 | Chronic neurological comorbidities | Flaccid tetraplegia |
| G824 | Chronic neurological comorbidities | Spastic tetraplegia |
| G825 | Chronic neurological comorbidities | Tetraplegia, unspecified |
| G83X | Chronic neurological comorbidities | Other paralytic syndromes |
| G830 | Chronic neurological comorbidities | Diplegia of upper limbs |
| G831 | Chronic neurological comorbidities | Monoplegia of lower limb |
| G832 | Chronic neurological comorbidities | Monoplegia of upper limb |
| G833 | Chronic neurological comorbidities | Monoplegia, unspecified |
| G834 | Chronic neurological comorbidities | Cauda equina syndrome |
| G835 | Chronic neurological comorbidities | Locked-in syndrome |
| G838 | Chronic neurological comorbidities | Other specified paralytic syndromes |
| G839 | Chronic neurological comorbidities | Paralytic syndrome, unspecified |

##### Dementia

| **ICD-10 code** | **Category** | **Description** |
| --- | --- | --- |
| F00X | Dementia | Dementia in Alzheimer disease |
| F000 | Dementia | Dementia in Alzheimer disease with early onset |
| F001 | Dementia | Dementia in Alzheimer disease with late onset |
| F002 | Dementia | Dementia in Alzheimer disease, atypical or mixed type |
| F009 | Dementia | Dementia in Alzheimer disease, unspecified |
| F01X | Dementia | Vascular dementia |
| F010 | Dementia | Vascular dementia of acute onset |
| F011 | Dementia | Multi-infarct dementia |
| F012 | Dementia | Subcortical vascular dementia |
| F013 | Dementia | Mixed cortical and subcortical vascular dementia |
| F018 | Dementia | Other vascular dementia |
| F019 | Dementia | Vascular dementia, unspecified |
| F02X | Dementia | Dementia in other diseases classified elsewhere |
| F020 | Dementia | Dementia in pick disease |
| F021 | Dementia | Dementia in Creutzfeldt-Jakob disease |
| F022 | Dementia | Dementia in Huntington disease |
| F023 | Dementia | Dementia in Parkinson disease |
| F024 | Dementia | Dementia in human immunodeficiency virus (hiv) disease |
| F028 | Dementia | Dementia in other specified diseases classified elsewhere |
| F03X | Dementia | Unspecified dementia |
| G30X | Dementia | Alzheimer disease |
| G300 | Dementia | Alzheimer disease with early onset |
| G301 | Dementia | Alzheimer disease with late onset |
| G308 | Dementia | Other Alzheimer disease |
| G309 | Dementia | Alzheimer disease, unspecified |

##### CTD

| **ICD-10 code** | **Category** | **Description** |
| --- | --- | --- |
| M050 | Chronic connective tissue disease comorbidities | Felty syndrome |
| M0500 | Chronic connective tissue disease comorbidities | Felty syndrome: multiple sites |
| M0501 | Chronic connective tissue disease comorbidities | Felty syndrome: shoulder region |
| M0502 | Chronic connective tissue disease comorbidities | Felty syndrome: upper arm |
| M0503 | Chronic connective tissue disease comorbidities | Felty syndrome: forearm |
| M0504 | Chronic connective tissue disease comorbidities | Felty syndrome: hand |
| M0505 | Chronic connective tissue disease comorbidities | Felty syndrome: pelvic region and thigh |
| M0506 | Chronic connective tissue disease comorbidities | Felty syndrome: lower leg |
| M0507 | Chronic connective tissue disease comorbidities | Felty syndrome: ankle and foot |
| M0508 | Chronic connective tissue disease comorbidities | Felty syndrome: other |
| M0509 | Chronic connective tissue disease comorbidities | Felty syndrome: site unspecified |
| M051 | Chronic connective tissue disease comorbidities | Rheumatoid lung disease |
| M0510 | Chronic connective tissue disease comorbidities | Rheumatoid lung disease: multiple sites |
| M0511 | Chronic connective tissue disease comorbidities | Rheumatoid lung disease: shoulder region |
| M0512 | Chronic connective tissue disease comorbidities | Rheumatoid lung disease: upper arm |
| M0513 | Chronic connective tissue disease comorbidities | Rheumatoid lung disease: forearm |
| M0514 | Chronic connective tissue disease comorbidities | Rheumatoid lung disease: hand |
| M0515 | Chronic connective tissue disease comorbidities | Rheumatoid lung disease: pelvic region and thigh |
| M0516 | Chronic connective tissue disease comorbidities | Rheumatoid lung disease: lower leg |
| M0517 | Chronic connective tissue disease comorbidities | Rheumatoid lung disease: ankle and foot |
| M0518 | Chronic connective tissue disease comorbidities | Rheumatoid lung disease: other |
| M0519 | Chronic connective tissue disease comorbidities | Rheumatoid lung disease: site unspecified |
| M052 | Chronic connective tissue disease comorbidities | Rheumatoid vasculitis |
| M0520 | Chronic connective tissue disease comorbidities | Rheumatoid vasculitis: multiple sites |
| M0521 | Chronic connective tissue disease comorbidities | Rheumatoid vasculitis: shoulder region |
| M0522 | Chronic connective tissue disease comorbidities | Rheumatoid vasculitis: upper arm |
| M0523 | Chronic connective tissue disease comorbidities | Rheumatoid vasculitis: forearm |
| M0524 | Chronic connective tissue disease comorbidities | Rheumatoid vasculitis: hand |
| M0525 | Chronic connective tissue disease comorbidities | Rheumatoid vasculitis: pelvic region and thigh |
| M0526 | Chronic connective tissue disease comorbidities | Rheumatoid vasculitis: lower leg |
| M0527 | Chronic connective tissue disease comorbidities | Rheumatoid vasculitis: ankle and foot |
| M0528 | Chronic connective tissue disease comorbidities | Rheumatoid vasculitis: other |
| M0529 | Chronic connective tissue disease comorbidities | Rheumatoid vasculitis: site unspecified |
| M053 | Chronic connective tissue disease comorbidities | Rheumatoid arthritis with involvement of other organs and systems |
| M0530 | Chronic connective tissue disease comorbidities | Rheumatoid arthritis with involvement of other organs and systems: multiple sites |
| M0531 | Chronic connective tissue disease comorbidities | Rheumatoid arthritis with involvement of other organs and systems: shoulder region |
| M0532 | Chronic connective tissue disease comorbidities | Rheumatoid arthritis with involvement of other organs and systems: upper arm |
| M0533 | Chronic connective tissue disease comorbidities | Rheumatoid arthritis with involvement of other organs and systems: forearm |
| M0534 | Chronic connective tissue disease comorbidities | Rheumatoid arthritis with involvement of other organs and systems: hand |
| M0535 | Chronic connective tissue disease comorbidities | Rheumatoid arthritis with involvement of other organs and systems: pelvic region and thigh |
| M0536 | Chronic connective tissue disease comorbidities | Rheumatoid arthritis with involvement of other organs and systems: lower leg |
| M0537 | Chronic connective tissue disease comorbidities | Rheumatoid arthritis with involvement of other organs and systems: ankle and foot |
| M0538 | Chronic connective tissue disease comorbidities | Rheumatoid arthritis with involvement of other organs and systems: other |
| M0539 | Chronic connective tissue disease comorbidities | Rheumatoid arthritis with involvement of other organs and systems: site unspecified |
| M058 | Chronic connective tissue disease comorbidities | Other seropositive rheumatoid arthritis |
| M0580 | Chronic connective tissue disease comorbidities | Other seropositive rheumatoid arthritis: multiple sites |
| M0581 | Chronic connective tissue disease comorbidities | Other seropositive rheumatoid arthritis: shoulder region |
| M0582 | Chronic connective tissue disease comorbidities | Other seropositive rheumatoid arthritis: upper arm |
| M0583 | Chronic connective tissue disease comorbidities | Other seropositive rheumatoid arthritis: forearm |
| M0584 | Chronic connective tissue disease comorbidities | Other seropositive rheumatoid arthritis: hand |
| M0585 | Chronic connective tissue disease comorbidities | Other seropositive rheumatoid arthritis: pelvic region and thigh |
| M0586 | Chronic connective tissue disease comorbidities | Other seropositive rheumatoid arthritis: lower leg |
| M0587 | Chronic connective tissue disease comorbidities | Other seropositive rheumatoid arthritis: ankle and foot |
| M0588 | Chronic connective tissue disease comorbidities | Other seropositive rheumatoid arthritis: other |
| M0589 | Chronic connective tissue disease comorbidities | Other seropositive rheumatoid arthritis: site unspecified |
| M059 | Chronic connective tissue disease comorbidities | Seropositive rheumatoid arthritis, unspecified |
| M0590 | Chronic connective tissue disease comorbidities | Seropositive rheumatoid arthritis, unspecified: multiple sites |
| M0591 | Chronic connective tissue disease comorbidities | Seropositive rheumatoid arthritis, unspecified: shoulder region |
| M0592 | Chronic connective tissue disease comorbidities | Seropositive rheumatoid arthritis, unspecified: upper arm |
| M0593 | Chronic connective tissue disease comorbidities | Seropositive rheumatoid arthritis, unspecified: forearm |
| M0594 | Chronic connective tissue disease comorbidities | Seropositive rheumatoid arthritis, unspecified: hand |
| M0595 | Chronic connective tissue disease comorbidities | Seropositive rheumatoid arthritis, unspecified: pelvic region and thigh |
| M0596 | Chronic connective tissue disease comorbidities | Seropositive rheumatoid arthritis, unspecified: lower leg |
| M0597 | Chronic connective tissue disease comorbidities | Seropositive rheumatoid arthritis, unspecified: ankle and foot |
| M0598 | Chronic connective tissue disease comorbidities | Seropositive rheumatoid arthritis, unspecified: other |
| M0599 | Chronic connective tissue disease comorbidities | Seropositive rheumatoid arthritis, unspecified: site unspecified |
| M06X | Chronic connective tissue disease comorbidities | Other rheumatoid arthritis |
| M060 | Chronic connective tissue disease comorbidities | Seronegative rheumatoid arthritis |
| M0600 | Chronic connective tissue disease comorbidities | Seronegative rheumatoid arthritis: multiple sites |
| M0601 | Chronic connective tissue disease comorbidities | Seronegative rheumatoid arthritis: shoulder region |
| M0602 | Chronic connective tissue disease comorbidities | Seronegative rheumatoid arthritis: upper arm |
| M0603 | Chronic connective tissue disease comorbidities | Seronegative rheumatoid arthritis: forearm |
| M0604 | Chronic connective tissue disease comorbidities | Seronegative rheumatoid arthritis: hand |
| M0605 | Chronic connective tissue disease comorbidities | Seronegative rheumatoid arthritis: pelvic region and thigh |
| M0606 | Chronic connective tissue disease comorbidities | Seronegative rheumatoid arthritis: lower leg |
| M0607 | Chronic connective tissue disease comorbidities | Seronegative rheumatoid arthritis: ankle and foot |
| M0608 | Chronic connective tissue disease comorbidities | Seronegative rheumatoid arthritis: other |
| M0609 | Chronic connective tissue disease comorbidities | Seronegative rheumatoid arthritis: site unspecified |
| M061 | Chronic connective tissue disease comorbidities | Adult-onset still disease |
| M0610 | Chronic connective tissue disease comorbidities | Adult-onset still disease: multiple sites |
| M0611 | Chronic connective tissue disease comorbidities | Adult-onset still disease: shoulder region |
| M0612 | Chronic connective tissue disease comorbidities | Adult-onset still disease: upper arm |
| M0613 | Chronic connective tissue disease comorbidities | Adult-onset still disease: forearm |
| M0614 | Chronic connective tissue disease comorbidities | Adult-onset still disease: hand |
| M0615 | Chronic connective tissue disease comorbidities | Adult-onset still disease: pelvic region and thigh |
| M0616 | Chronic connective tissue disease comorbidities | Adult-onset still disease: lower leg |
| M0617 | Chronic connective tissue disease comorbidities | Adult-onset still disease: ankle and foot |
| M0618 | Chronic connective tissue disease comorbidities | Adult-onset still disease: other |
| M0619 | Chronic connective tissue disease comorbidities | Adult-onset still disease: site unspecified |
| M062 | Chronic connective tissue disease comorbidities | Rheumatoid bursitis |
| M0620 | Chronic connective tissue disease comorbidities | Rheumatoid bursitis: multiple sites |
| M0621 | Chronic connective tissue disease comorbidities | Rheumatoid bursitis: shoulder region |
| M0622 | Chronic connective tissue disease comorbidities | Rheumatoid bursitis: upper arm |
| M0623 | Chronic connective tissue disease comorbidities | Rheumatoid bursitis: forearm |
| M0624 | Chronic connective tissue disease comorbidities | Rheumatoid bursitis: hand |
| M0625 | Chronic connective tissue disease comorbidities | Rheumatoid bursitis: pelvic region and thigh |
| M0626 | Chronic connective tissue disease comorbidities | Rheumatoid bursitis: lower leg |
| M0627 | Chronic connective tissue disease comorbidities | Rheumatoid bursitis: ankle and foot |
| M0628 | Chronic connective tissue disease comorbidities | Rheumatoid bursitis: other |
| M0629 | Chronic connective tissue disease comorbidities | Rheumatoid bursitis: site unspecified |
| M063 | Chronic connective tissue disease comorbidities | Rheumatoid nodule |
| M0630 | Chronic connective tissue disease comorbidities | Rheumatoid nodule: multiple sites |
| M0631 | Chronic connective tissue disease comorbidities | Rheumatoid nodule: shoulder region |
| M0632 | Chronic connective tissue disease comorbidities | Rheumatoid nodule: upper arm |
| M0633 | Chronic connective tissue disease comorbidities | Rheumatoid nodule: forearm |
| M0634 | Chronic connective tissue disease comorbidities | Rheumatoid nodule: hand |
| M0635 | Chronic connective tissue disease comorbidities | Rheumatoid nodule: pelvic region and thigh |
| M0636 | Chronic connective tissue disease comorbidities | Rheumatoid nodule: lower leg |
| M0637 | Chronic connective tissue disease comorbidities | Rheumatoid nodule: ankle and foot |
| M0638 | Chronic connective tissue disease comorbidities | Rheumatoid nodule: other |
| M0639 | Chronic connective tissue disease comorbidities | Rheumatoid nodule: site unspecified |
| M064 | Chronic connective tissue disease comorbidities | Inflammatory polyarthropathy |
| M0640 | Chronic connective tissue disease comorbidities | Inflammatory polyarthropathy: multiple sites |
| M0641 | Chronic connective tissue disease comorbidities | Inflammatory polyarthropathy: shoulder region |
| M0642 | Chronic connective tissue disease comorbidities | Inflammatory polyarthropathy: upper arm |
| M0643 | Chronic connective tissue disease comorbidities | Inflammatory polyarthropathy: forearm |
| M0644 | Chronic connective tissue disease comorbidities | Inflammatory polyarthropathy: hand |
| M0645 | Chronic connective tissue disease comorbidities | Inflammatory polyarthropathy: pelvic region and thigh |
| M0646 | Chronic connective tissue disease comorbidities | Inflammatory polyarthropathy: lower leg |
| M0647 | Chronic connective tissue disease comorbidities | Inflammatory polyarthropathy: ankle and foot |
| M0648 | Chronic connective tissue disease comorbidities | Inflammatory polyarthropathy: other |
| M0649 | Chronic connective tissue disease comorbidities | Inflammatory polyarthropathy: site unspecified |
| M068 | Chronic connective tissue disease comorbidities | Other specified rheumatoid arthritis |
| M0680 | Chronic connective tissue disease comorbidities | Other specified rheumatoid arthritis: multiple sites |
| M0681 | Chronic connective tissue disease comorbidities | Other specified rheumatoid arthritis: shoulder region |
| M0682 | Chronic connective tissue disease comorbidities | Other specified rheumatoid arthritis: upper arm |
| M0683 | Chronic connective tissue disease comorbidities | Other specified rheumatoid arthritis: forearm |
| M0684 | Chronic connective tissue disease comorbidities | Other specified rheumatoid arthritis: hand |
| M0685 | Chronic connective tissue disease comorbidities | Other specified rheumatoid arthritis: pelvic region and thigh |
| M0686 | Chronic connective tissue disease comorbidities | Other specified rheumatoid arthritis: lower leg |
| M0687 | Chronic connective tissue disease comorbidities | Other specified rheumatoid arthritis: ankle and foot |
| M0688 | Chronic connective tissue disease comorbidities | Other specified rheumatoid arthritis: other |
| M0689 | Chronic connective tissue disease comorbidities | Other specified rheumatoid arthritis: site unspecified |
| M069 | Chronic connective tissue disease comorbidities | Rheumatoid arthritis, unspecified |
| M0690 | Chronic connective tissue disease comorbidities | Rheumatoid arthritis, unspecified: multiple sites |
| M0691 | Chronic connective tissue disease comorbidities | Rheumatoid arthritis, unspecified: shoulder region |
| M0692 | Chronic connective tissue disease comorbidities | Rheumatoid arthritis, unspecified: upper arm |
| M0693 | Chronic connective tissue disease comorbidities | Rheumatoid arthritis, unspecified: forearm |
| M0694 | Chronic connective tissue disease comorbidities | Rheumatoid arthritis, unspecified: hand |
| M0695 | Chronic connective tissue disease comorbidities | Rheumatoid arthritis, unspecified: pelvic region and thigh |
| M0696 | Chronic connective tissue disease comorbidities | Rheumatoid arthritis, unspecified: lower leg |
| M0697 | Chronic connective tissue disease comorbidities | Rheumatoid arthritis, unspecified: ankle and foot |
| M0698 | Chronic connective tissue disease comorbidities | Rheumatoid arthritis, unspecified: other |
| M0699 | Chronic connective tissue disease comorbidities | Rheumatoid arthritis, unspecified: site unspecified |
| M07X | Chronic connective tissue disease comorbidities | Psoriatic and enteropathic arthropathies |
| M070 | Chronic connective tissue disease comorbidities | Distal interphalangeal psoriatic arthropathy |
| M0700 | Chronic connective tissue disease comorbidities | Distal interphalangeal psoriatic arthropathy: multiple sites |
| M0704 | Chronic connective tissue disease comorbidities | Distal interphalangeal psoriatic arthropathy: hand |
| M0707 | Chronic connective tissue disease comorbidities | Distal interphalangeal psoriatic arthropathy: ankle and foot |
| M0709 | Chronic connective tissue disease comorbidities | Distal interphalangeal psoriatic arthropathy: site unspecified |
| M071 | Chronic connective tissue disease comorbidities | Arthritis mutilans |
| M0710 | Chronic connective tissue disease comorbidities | Arthritis mutilans: multiple sites |
| M0711 | Chronic connective tissue disease comorbidities | Arthritis mutilans: shoulder region |
| M0712 | Chronic connective tissue disease comorbidities | Arthritis mutilans: upper arm |
| M0713 | Chronic connective tissue disease comorbidities | Arthritis mutilans: forearm |
| M0714 | Chronic connective tissue disease comorbidities | Arthritis mutilans: hand |
| M0715 | Chronic connective tissue disease comorbidities | Arthritis mutilans: pelvic region and thigh |
| M0716 | Chronic connective tissue disease comorbidities | Arthritis mutilans: lower leg |
| M0717 | Chronic connective tissue disease comorbidities | Arthritis mutilans: ankle and foot |
| M0718 | Chronic connective tissue disease comorbidities | Arthritis mutilans: other |
| M0719 | Chronic connective tissue disease comorbidities | Arthritis mutilans: site unspecified |
| M072 | Chronic connective tissue disease comorbidities | Psoriatic spondylitis |
| M0720 | Chronic connective tissue disease comorbidities | Psoriatic spondylitis: multiple sites |
| M0721 | Chronic connective tissue disease comorbidities | Psoriatic spondylitis: shoulder region |
| M0722 | Chronic connective tissue disease comorbidities | Psoriatic spondylitis: upper arm |
| M0723 | Chronic connective tissue disease comorbidities | Psoriatic spondylitis: forearm |
| M0724 | Chronic connective tissue disease comorbidities | Psoriatic spondylitis: hand |
| M0725 | Chronic connective tissue disease comorbidities | Psoriatic spondylitis: pelvic region and thigh |
| M0726 | Chronic connective tissue disease comorbidities | Psoriatic spondylitis: lower leg |
| M0727 | Chronic connective tissue disease comorbidities | Psoriatic spondylitis: ankle and foot |
| M0728 | Chronic connective tissue disease comorbidities | Psoriatic spondylitis: other |
| M0729 | Chronic connective tissue disease comorbidities | Psoriatic spondylitis: site unspecified |
| M073 | Chronic connective tissue disease comorbidities | Other psoriatic arthropathies |
| M0730 | Chronic connective tissue disease comorbidities | Other psoriatic arthropathies: multiple sites |
| M0731 | Chronic connective tissue disease comorbidities | Other psoriatic arthropathies: shoulder region |
| M0732 | Chronic connective tissue disease comorbidities | Other psoriatic arthropathies: upper arm |
| M0733 | Chronic connective tissue disease comorbidities | Other psoriatic arthropathies: forearm |
| M0734 | Chronic connective tissue disease comorbidities | Other psoriatic arthropathies: hand |
| M0735 | Chronic connective tissue disease comorbidities | Other psoriatic arthropathies: pelvic region and thigh |
| M0736 | Chronic connective tissue disease comorbidities | Other psoriatic arthropathies: lower leg |
| M0737 | Chronic connective tissue disease comorbidities | Other psoriatic arthropathies: ankle and foot |
| M0738 | Chronic connective tissue disease comorbidities | Other psoriatic arthropathies: other |
| M0739 | Chronic connective tissue disease comorbidities | Other psoriatic arthropathies: site unspecified |
| M074 | Chronic connective tissue disease comorbidities | Arthropathy in Crohn disease (regional enteritis) |
| M0740 | Chronic connective tissue disease comorbidities | Arthropathy in Crohn disease (regional enteritis): multiple sites |
| M0741 | Chronic connective tissue disease comorbidities | Arthropathy in Crohn disease (regional enteritis): shoulder region |
| M0742 | Chronic connective tissue disease comorbidities | Arthropathy in Crohn disease (regional enteritis): upper arm |
| M0743 | Chronic connective tissue disease comorbidities | Arthropathy in Crohn disease (regional enteritis): forearm |
| M0744 | Chronic connective tissue disease comorbidities | Arthropathy in Crohn disease (regional enteritis): hand |
| M0745 | Chronic connective tissue disease comorbidities | Arthropathy in Crohn disease (regional enteritis): pelvic region and thigh |
| M0746 | Chronic connective tissue disease comorbidities | Arthropathy in Crohn disease (regional enteritis): lower leg |
| M0747 | Chronic connective tissue disease comorbidities | Arthropathy in Crohn disease (regional enteritis): ankle and foot |
| M0748 | Chronic connective tissue disease comorbidities | Arthropathy in Crohn disease (regional enteritis): other |
| M0749 | Chronic connective tissue disease comorbidities | Arthropathy in Crohn disease (regional enteritis): site unspecified |
| M075 | Chronic connective tissue disease comorbidities | Arthropathy in ulcerative colitis |
| M0750 | Chronic connective tissue disease comorbidities | Arthropathy in ulcerative colitis: multiple sites |
| M0751 | Chronic connective tissue disease comorbidities | Arthropathy in ulcerative colitis: shoulder region |
| M0752 | Chronic connective tissue disease comorbidities | Arthropathy in ulcerative colitis: upper arm |
| M0753 | Chronic connective tissue disease comorbidities | Arthropathy in ulcerative colitis: forearm |
| M0754 | Chronic connective tissue disease comorbidities | Arthropathy in ulcerative colitis: hand |
| M0755 | Chronic connective tissue disease comorbidities | Arthropathy in ulcerative colitis: pelvic region and thigh |
| M0756 | Chronic connective tissue disease comorbidities | Arthropathy in ulcerative colitis: lower leg |
| M0757 | Chronic connective tissue disease comorbidities | Arthropathy in ulcerative colitis: ankle and foot |
| M0758 | Chronic connective tissue disease comorbidities | Arthropathy in ulcerative colitis: other |
| M0759 | Chronic connective tissue disease comorbidities | Arthropathy in ulcerative colitis: site unspecified |
| M076 | Chronic connective tissue disease comorbidities | Other enteropathic arthropathies |
| M0760 | Chronic connective tissue disease comorbidities | Other enteropathic arthropathies: multiple sites |
| M0761 | Chronic connective tissue disease comorbidities | Other enteropathic arthropathies: shoulder region |
| M0762 | Chronic connective tissue disease comorbidities | Other enteropathic arthropathies: upper arm |
| M0763 | Chronic connective tissue disease comorbidities | Other enteropathic arthropathies: forearm |
| M0764 | Chronic connective tissue disease comorbidities | Other enteropathic arthropathies: hand |
| M0765 | Chronic connective tissue disease comorbidities | Other enteropathic arthropathies: pelvic region and thigh |
| M0766 | Chronic connective tissue disease comorbidities | Other enteropathic arthropathies: lower leg |
| M0767 | Chronic connective tissue disease comorbidities | Other enteropathic arthropathies: ankle and foot |
| M0768 | Chronic connective tissue disease comorbidities | Other enteropathic arthropathies: other |
| M0769 | Chronic connective tissue disease comorbidities | Other enteropathic arthropathies: site unspecified |
| M30X | Chronic connective tissue disease comorbidities | Polyarteritis nodosa and related conditions |
| M300 | Chronic connective tissue disease comorbidities | Polyarteritis nodosa |
| M301 | Chronic connective tissue disease comorbidities | Polyarteritis with lung involvement (churg-strauss) |
| M302 | Chronic connective tissue disease comorbidities | Juvenile polyarteritis |
| M303 | Chronic connective tissue disease comorbidities | Mucocutaneous lymph node syndrome (kawasaki) |
| M308 | Chronic connective tissue disease comorbidities | Other conditions related to polyarteritis nodosa |
| M31X | Chronic connective tissue disease comorbidities | Other necrotizing vasculopathies |
| M310 | Chronic connective tissue disease comorbidities | Hypersensitivity angiitis |
| M311 | Chronic connective tissue disease comorbidities | Thrombotic microangiopathy |
| M312 | Chronic connective tissue disease comorbidities | Lethal midline granuloma |
| M313 | Chronic connective tissue disease comorbidities | Wegener granulomatosis |
| M314 | Chronic connective tissue disease comorbidities | Aortic arch syndrome (takayasu) |
| M315 | Chronic connective tissue disease comorbidities | Giant cell arteritis with polymyalgia rheumatica |
| M316 | Chronic connective tissue disease comorbidities | Other giant cell arteritis |
| M317 | Chronic connective tissue disease comorbidities | Microscopic polyangiitis |
| M318 | Chronic connective tissue disease comorbidities | Other specified necrotizing vasculopathies |
| M319 | Chronic connective tissue disease comorbidities | Necrotizing vasculopathy, unspecified |
| M32X | Chronic connective tissue disease comorbidities | Systemic lupus erythematosus |
| M320 | Chronic connective tissue disease comorbidities | Drug-induced systemic lupus erythematosus |
| M321 | Chronic connective tissue disease comorbidities | Systemic lupus erythematosus with organ or system involvement |
| M328 | Chronic connective tissue disease comorbidities | Other forms of systemic lupus erythematosus |
| M329 | Chronic connective tissue disease comorbidities | Systemic lupus erythematosus, unspecified |
| M33X | Chronic connective tissue disease comorbidities | Dermatopolymyositis |
| M330 | Chronic connective tissue disease comorbidities | Juvenile dermatomyositis |
| M331 | Chronic connective tissue disease comorbidities | Other dermatomyositis |
| M332 | Chronic connective tissue disease comorbidities | Polymyositis |
| M339 | Chronic connective tissue disease comorbidities | Dermatopolymyositis, unspecified |
| M34X | Chronic connective tissue disease comorbidities | Systemic sclerosis |
| M340 | Chronic connective tissue disease comorbidities | Progressive systemic sclerosis |
| M341 | Chronic connective tissue disease comorbidities | Cr(e)st syndrome |
| M342 | Chronic connective tissue disease comorbidities | Systemic sclerosis induced by drugs and chemicals |
| M348 | Chronic connective tissue disease comorbidities | Other forms of systemic sclerosis |
| M349 | Chronic connective tissue disease comorbidities | Systemic sclerosis, unspecified |
| M35X | Chronic connective tissue disease comorbidities | Other systemic involvement of connective tissue |
| M350 | Chronic connective tissue disease comorbidities | Sicca syndrome (sjögren) |
| M351 | Chronic connective tissue disease comorbidities | Other overlap syndromes |
| M352 | Chronic connective tissue disease comorbidities | Behçet disease |
| M353 | Chronic connective tissue disease comorbidities | Polymyalgia rheumatica |
| M354 | Chronic connective tissue disease comorbidities | Diffuse (eosinophilic) fasciitis |
| M355 | Chronic connective tissue disease comorbidities | Multifocal fibrosclerosis |
| M356 | Chronic connective tissue disease comorbidities | Relapsing panniculitis (weber-christian) |
| M357 | Chronic connective tissue disease comorbidities | Hypermobility syndrome |
| M358 | Chronic connective tissue disease comorbidities | Other specified systemic involvement of connective tissue |
| M359 | Chronic connective tissue disease comorbidities | Systemic involvement of connective tissue, unspecified |
| M36X | Chronic connective tissue disease comorbidities | Systemic disorders of connective tissue in diseases classified elsewhere |
| M360 | Chronic connective tissue disease comorbidities | Dermato(poly)myositis in neoplastic disease |
| M361 | Chronic connective tissue disease comorbidities | Arthropathy in neoplastic disease |
| M362 | Chronic connective tissue disease comorbidities | Haemophilic arthropathy |
| M363 | Chronic connective tissue disease comorbidities | Arthropathy in other blood disorders |
| M364 | Chronic connective tissue disease comorbidities | Arthropathy in hypersensitivity reactions classified elsewhere |
| M368 | Chronic connective tissue disease comorbidities | Systemic disorders of connective tissue in other diseases classified elsewhere |
| M45X | Chronic connective tissue disease comorbidities | Ankylosing spondylitis |
| M45X0 | Chronic connective tissue disease comorbidities | Ankylosing spondylitis: multiple sites in spine |
| M45X1 | Chronic connective tissue disease comorbidities | Ankylosing spondylitis: occipito-atlanto-axial region |
| M45X2 | Chronic connective tissue disease comorbidities | Ankylosing spondylitis: cervical region |
| M45X3 | Chronic connective tissue disease comorbidities | Ankylosing spondylitis: cervicothoracic region |
| M45X4 | Chronic connective tissue disease comorbidities | Ankylosing spondylitis: thoracic region |
| M45X5 | Chronic connective tissue disease comorbidities | Ankylosing spondylitis: thoracolumbar region |
| M45X6 | Chronic connective tissue disease comorbidities | Ankylosing spondylitis: lumbar region |
| M45X7 | Chronic connective tissue disease comorbidities | Ankylosing spondylitis: lumbosacral region |
| M45X8 | Chronic connective tissue disease comorbidities | Ankylosing spondylitis: sacral and sacrococcygeal region |
| M45X9 | Chronic connective tissue disease comorbidities | Ankylosing spondylitis: site unspecified |

##### HTN

| **ICD-10 code** | **Category** | **Description** |
| --- | --- | --- |
| I10X | Hypertension | Essential (primary) hypertension |
| I15X | Hypertension | Secondary hypertension |
| I150 | Hypertension | Renovascular hypertension |
| I151 | Hypertension | Hypertension secondary to other renal disorders |
| I152 | Hypertension | Hypertension secondary to endocrine disorders |
| I158 | Hypertension | Other secondary hypertension |
| I159 | Hypertension | Secondary hypertension, unspecified |

##### Diabetes

| **ICD-10 code** | **Category** | **Description** |
| --- | --- | --- |
| E10X | Diabetes Mellitus | Type 1 diabetes mellitus |
| E100 | Diabetes Mellitus | Type 1 diabetes mellitus: with coma |
| E101 | Diabetes Mellitus | Type 1 diabetes mellitus: with ketoacidosis |
| E102 | Diabetes Mellitus | Type 1 diabetes mellitus: with renal complications |
| E103 | Diabetes Mellitus | Type 1 diabetes mellitus: with ophthalmic complications |
| E104 | Diabetes Mellitus | Type 1 diabetes mellitus: with neurological complications |
| E105 | Diabetes Mellitus | Type 1 diabetes mellitus: with peripheral circulatory complications |
| E106 | Diabetes Mellitus | Type 1 diabetes mellitus: with other specified complications |
| E107 | Diabetes Mellitus | Type 1 diabetes mellitus: with multiple complications |
| E108 | Diabetes Mellitus | Type 1 diabetes mellitus: with unspecified complications |
| E109 | Diabetes Mellitus | Type 1 diabetes mellitus: without complications |
| E11X | Diabetes Mellitus | Type 2 diabetes mellitus |
| E110 | Diabetes Mellitus | Type 2 diabetes mellitus: with coma |
| E111 | Diabetes Mellitus | Type 2 diabetes mellitus: with ketoacidosis |
| E112 | Diabetes Mellitus | Type 2 diabetes mellitus: with renal complications |
| E113 | Diabetes Mellitus | Type 2 diabetes mellitus: with ophthalmic complications |
| E114 | Diabetes Mellitus | Type 2 diabetes mellitus: with neurological complications |
| E115 | Diabetes Mellitus | Type 2 diabetes mellitus: with peripheral circulatory complications |
| E116 | Diabetes Mellitus | Type 2 diabetes mellitus: with other specified complications |
| E117 | Diabetes Mellitus | Type 2 diabetes mellitus: with multiple complications |
| E118 | Diabetes Mellitus | Type 2 diabetes mellitus: with unspecified complications |
| E119 | Diabetes Mellitus | Type 2 diabetes mellitus: without complications |
| E12X | Diabetes Mellitus | Malnutrition-related diabetes mellitus |
| E120 | Diabetes Mellitus | Malnutrition-related diabetes mellitus: with coma |
| E121 | Diabetes Mellitus | Malnutrition-related diabetes mellitus: with ketoacidosis |
| E122 | Diabetes Mellitus | Malnutrition-related diabetes mellitus: with renal complications |
| E123 | Diabetes Mellitus | Malnutrition-related diabetes mellitus: with ophthalmic complications |
| E124 | Diabetes Mellitus | Malnutrition-related diabetes mellitus: with neurological complications |
| E125 | Diabetes Mellitus | Malnutrition-related diabetes mellitus: with peripheral circulatory complications |
| E126 | Diabetes Mellitus | Malnutrition-related diabetes mellitus: with other specified complications |
| E127 | Diabetes Mellitus | Malnutrition-related diabetes mellitus: with multiple complications |
| E128 | Diabetes Mellitus | Malnutrition-related diabetes mellitus: with unspecified complications |
| E129 | Diabetes Mellitus | Malnutrition-related diabetes mellitus: without complications |
| E13X | Diabetes Mellitus | Other specified diabetes mellitus |
| E130 | Diabetes Mellitus | Other specified diabetes mellitus: with coma |
| E131 | Diabetes Mellitus | Other specified diabetes mellitus: with ketoacidosis |
| E132 | Diabetes Mellitus | Other specified diabetes mellitus: with renal complications |
| E133 | Diabetes Mellitus | Other specified diabetes mellitus: with ophthalmic complications |
| E134 | Diabetes Mellitus | Other specified diabetes mellitus: with neurological complications |
| E135 | Diabetes Mellitus | Other specified diabetes mellitus: with peripheral circulatory complications |
| E136 | Diabetes Mellitus | Other specified diabetes mellitus: with other specified complications |
| E137 | Diabetes Mellitus | Other specified diabetes mellitus: with multiple complications |
| E138 | Diabetes Mellitus | Other specified diabetes mellitus: with unspecified complications |
| E139 | Diabetes Mellitus | Other specified diabetes mellitus: without complications |
| E14X | Diabetes Mellitus | Unspecified diabetes mellitus |
| E140 | Diabetes Mellitus | Unspecified diabetes mellitus: with coma |
| E141 | Diabetes Mellitus | Unspecified diabetes mellitus: with ketoacidosis |
| E142 | Diabetes Mellitus | Unspecified diabetes mellitus: with renal complications |
| E143 | Diabetes Mellitus | Unspecified diabetes mellitus: with ophthalmic complications |
| E144 | Diabetes Mellitus | Unspecified diabetes mellitus: with neurological complications |
| E145 | Diabetes Mellitus | Unspecified diabetes mellitus: with peripheral circulatory complications |
| E146 | Diabetes Mellitus | Unspecified diabetes mellitus: with other specified complications |
| E147 | Diabetes Mellitus | Unspecified diabetes mellitus: with multiple complications |
| E148 | Diabetes Mellitus | Unspecified diabetes mellitus: with unspecified complications |
| E149 | Diabetes Mellitus | Unspecified diabetes mellitus: without complications |

##### HIV

| **ICD-10 code** | **Category** | **Description** |
| --- | --- | --- |
| B20X | HIV | Human immunodeficiency virus (hiv) disease resulting in infectious and parasitic diseases |
| B200 | HIV | HIV disease resulting in mycobacterial infection |
| B201 | HIV | HIV disease resulting in other bacterial infections |
| B202 | HIV | HIV disease resulting in cytomegaloviral disease |
| B203 | HIV | HIV disease resulting in other viral infections |
| B204 | HIV | HIV disease resulting in candidiasis |
| B205 | HIV | HIV disease resulting in other mycoses |
| B206 | HIV | HIV disease resulting in Pneumocystis jirovecii pneumonia |
| B207 | HIV | HIV disease resulting in multiple infections |
| B208 | HIV | HIV disease resulting in other infectious and parasitic diseases |
| B209 | HIV | HIV disease resulting in unspecified infectious or parasitic disease |
| B21X | HIV | Human immunodeficiency virus (hiv) disease resulting in malignant neoplasms |
| B210 | HIV | HIV disease resulting in Kaposi sarcoma |
| B211 | HIV | HIV disease resulting in Burkitt lymphoma |
| B212 | HIV | HIV disease resulting in other types of non-Hodgkin lymphoma |
| B213 | HIV | HIV disease resulting in other malignant neoplasms of lymphoid, haematopoietic and related tissue |
| B217 | HIV | HIV disease resulting in multiple malignant neoplasms |
| B218 | HIV | HIV disease resulting in other malignant neoplasms |
| B219 | HIV | HIV disease resulting in unspecified malignant neoplasm |
| B22X | HIV | Human immunodeficiency virus (hiv) disease resulting in other specified diseases |
| B220 | HIV | HIV disease resulting in encephalopathy |
| B221 | HIV | HIV disease resulting in lymphoid interstitial pneumonitis |
| B222 | HIV | HIV disease resulting in wasting syndrome |
| B227 | HIV | HIV disease resulting in multiple diseases classified elsewhere |
| B23X | HIV | Human immunodeficiency virus (hiv) disease resulting in other conditions |
| B230 | HIV | Acute HIV infection syndrome |
| B231 | HIV | HIV disease resulting in (persistent) generalized lymphadenopathy |
| B232 | HIV | HIV disease resulting in haematological and immunological abnormalities, not elsewhere classified |
| B238 | HIV | HIV disease resulting in other specified conditions |
| B24X | HIV | Unspecified human immunodeficiency virus (hiv) disease |

##### Solid Malig

| **ICD-10 code** | **Category** | **Description** |
| --- | --- | --- |
| C00X | Solid organ malignancies | Malignant neoplasm of lip |
| C021 | Solid organ malignancies | Malignant neoplasm: border of tongue |
| C098 | Solid organ malignancies | Malignant neoplasm: overlapping lesion of tonsil |
| C009 | Solid organ malignancies | Malignant neoplasm: lip, unspecified |
| C10X | Solid organ malignancies | Malignant neoplasm of oropharynx |
| C110 | Solid organ malignancies | Malignant neoplasm: superior wall of nasopharynx |
| C132 | Solid organ malignancies | Malignant neoplasm: posterior wall of hypopharynx |
| C154 | Solid organ malignancies | Malignant neoplasm: middle third of oesophagus |
| C165 | Solid organ malignancies | Malignant neoplasm: lesser curvature of stomach, unspecified |
| C187 | Solid organ malignancies | Malignant neoplasm: sigmoid colon |
| C109 | Solid organ malignancies | Malignant neoplasm: oropharynx, unspecified |
| C20X | Solid organ malignancies | Malignant neoplasm of rectum |
| C210 | Solid organ malignancies | Malignant neoplasm: anus, unspecified |
| C221 | Solid organ malignancies | Malignant neoplasm: intrahepatic bile duct carcinoma |
| C254 | Solid organ malignancies | Malignant neoplasm: endocrine pancreas |
| C30X | Solid organ malignancies | Malignant neoplasm of nasal cavity and middle ear |
| C310 | Solid organ malignancies | Malignant neoplasm: maxillary sinus |
| C321 | Solid organ malignancies | Malignant neoplasm: supraglottis |
| C343 | Solid organ malignancies | Malignant neoplasm: lower lobe, bronchus or lung |
| C398 | Solid organ malignancies | Malignant neoplasm: overlapping lesion of respiratory and intrathoracic organs |
| C40X | Solid organ malignancies | Malignant neoplasm of bone and articular cartilage of limbs |
| C410 | Solid organ malignancies | Malignant neoplasm: bones of skull and face |
| C432 | Solid organ malignancies | Malignant neoplasm: malignant melanoma of ear and external auricular canal |
| C443 | Solid organ malignancies | Malignant neoplasm: skin of other and unspecified parts of face |
| C476 | Solid organ malignancies | Malignant neoplasm: peripheral nerves of trunk, unspecified |
| C498 | Solid organ malignancies | Malignant neoplasm: overlapping lesion of connective and soft tissue |
| C409 | Solid organ malignancies | Malignant neoplasm: bone and articular cartilage of limb, unspecified |
| C50X | Solid organ malignancies | Malignant neoplasm of breast |
| C510 | Solid organ malignancies | Malignant neoplasm: labium majus |
| C543 | Solid organ malignancies | Malignant neoplasm: fundus uteri |
| C508 | Solid organ malignancies | Malignant neoplasm: overlapping lesion of breast |
| C519 | Solid organ malignancies | Malignant neoplasm: vulva, unspecified |
| C60X | Solid organ malignancies | Malignant neoplasm of penis |
| C621 | Solid organ malignancies | Malignant neoplasm: descended testis |
| C632 | Solid organ malignancies | Malignant neoplasm: scrotum |
| C676 | Solid organ malignancies | Malignant neoplasm: ureteric orifice |
| C698 | Solid organ malignancies | Malignant neoplasm: overlapping lesion of eye and adnexa |
| C609 | Solid organ malignancies | Malignant neoplasm: penis, unspecified |
| C70X | Solid organ malignancies | Malignant neoplasm of meninges |
| C710 | Solid organ malignancies | Malignant neoplasm: cerebrum, except lobes and ventricles |
| C721 | Solid organ malignancies | Malignant neoplasm: cauda equina |
| C754 | Solid organ malignancies | Malignant neoplasm: carotid body |
| C765 | Solid organ malignancies | Malignant neoplasm of other and ill-defined sites: lower limb |
| C787 | Solid organ malignancies | Secondary malignant neoplasm of liver and intrahepatic bile duct |
| C798 | Solid organ malignancies | Secondary malignant neoplasm of other specified sites |
| C709 | Solid organ malignancies | Malignant neoplasm: meninges, unspecified |
| C80X | Solid organ malignancies | Malignant neoplasm without specification of site |
| C800 | Solid organ malignancies | Malignant neoplasm, primary site unknown, so stated |
| C809 | Solid organ malignancies | Malignant neoplasm, primary site unspecified |
| C97X | Solid organ malignancies | Malignant neoplasms of independent (primary) multiple sites |

##### Haem Malig

| **ICD-10 code** | **Category** | **Description** |
| --- | --- | --- |
| C80X | Haematological malignancies | Malignant neoplasm without specification of site |
| C810 | Haematological malignancies | Nodular lymphocyte predominant Hodgkin lymphoma |
| C821 | Haematological malignancies | Follicular lymphoma grade II |
| C865 | Haematological malignancies | Angioimmunoblastic T-cell lymphoma |
| C829 | Haematological malignancies | Follicular lymphoma, unspecified |
| C90X | Haematological malignancies | Multiple myeloma and malignant plasma cell neoplasms |
| C900 | Haematological malignancies | Multiple myeloma |
| C901 | Haematological malignancies | Plasma cell leukaemia |
| C902 | Haematological malignancies | Extramedullary plasmacytoma |
| C903 | Haematological malignancies | Solitary plasmacytoma |
| C91X | Haematological malignancies | Lymphoid leukaemia |
| C910 | Haematological malignancies | Acute lymphoblastic leukaemia (all) |
| C911 | Haematological malignancies | Chronic lymphocytic leukaemia of b-cell type |
| C913 | Haematological malignancies | Prolymphocytic leukaemia of b-cell type |
| C914 | Haematological malignancies | Hairy-cell leukaemia |
| C915 | Haematological malignancies | Adult t-cell lymphoma/leukaemia (htlv-1-associated) |
| C916 | Haematological malignancies | Prolymphocytic leukaemia of t-cell type |
| C917 | Haematological malignancies | Other lymphoid leukaemia |
| C918 | Haematological malignancies | Mature B-cell leukaemia Burkitt-type |
| C919 | Haematological malignancies | Lymphoid leukaemia, unspecified |
| C92X | Haematological malignancies | Myeloid leukaemia |
| C920 | Haematological malignancies | Acute myeloblastic leukaemia (aml) |
| C921 | Haematological malignancies | Chronic myeloid leukaemia (cml), BCR/abl-positive |
| C922 | Haematological malignancies | Atypical chronic myeloid leukaemia, BCR/ABL-negative |
| C923 | Haematological malignancies | Myeloid sarcoma |
| C924 | Haematological malignancies | Acute promyelocytic leukaemia (pml) |
| C925 | Haematological malignancies | Acute myelomonocytic leukaemia |
| C926 | Haematological malignancies | Acute myeloid leukaemia with 11q23-abnormality |
| C927 | Haematological malignancies | Other myeloid leukaemia |
| C928 | Haematological malignancies | Acute myeloid leukaemia with multilineage dysplasia |
| C929 | Haematological malignancies | Myeloid leukaemia, unspecified |
| C93X | Haematological malignancies | Monocytic leukaemia |
| C930 | Haematological malignancies | Acute monoblastic/monocytic leukaemia |
| C931 | Haematological malignancies | Chronic myelomonocytic leukaemia |
| C933 | Haematological malignancies | Juvenile myelomonocytic leukaemia |
| C937 | Haematological malignancies | Other monocytic leukaemia |
| C939 | Haematological malignancies | Monocytic leukaemia, unspecified |
| C94X | Haematological malignancies | Other leukaemias of specified cell type |
| C940 | Haematological malignancies | Acute erythroid leukaemia |
| C942 | Haematological malignancies | Acute megakaryoblastic leukaemia |
| C943 | Haematological malignancies | Mast cell leukaemia |
| C944 | Haematological malignancies | Acute panmyelosis with myelofibrosis |
| C946 | Haematological malignancies | Myelodysplastic and myeloproliferative disease, not elsewhere classified |
| C947 | Haematological malignancies | Other specified leukaemias |
| C95X | Haematological malignancies | Leukaemia of unspecified cell type |
| C950 | Haematological malignancies | Acute leukaemia of unspecified cell type |
| C951 | Haematological malignancies | Chronic leukaemia of unspecified cell type |
| C957 | Haematological malignancies | Other leukaemia of unspecified cell type |
| C959 | Haematological malignancies | Leukaemia, unspecified |
| C96X | Haematological malignancies | Other and unspecified malignant neoplasms of lymphoid, haematopoietic and related tissue |
| C960 | Haematological malignancies | Multifocal and multisystemic (disseminated) Langerhans-cell histiocytosis (letterer-Siwe disease) |
| C962 | Haematological malignancies | Malignant mast cell tumour |
| C964 | Haematological malignancies | Sarcoma of dendritic cells (accessory cells) |
| C965 | Haematological malignancies | Multifocal and unisystemic Langerhans-cell histiocytosis |
| C966 | Haematological malignancies | Unifocal Langerhans-cell histiocytosis |
| C967 | Haematological malignancies | Other specified malignant neoplasms of lymphoid, haematopoietic and related tissue |
| C968 | Haematological malignancies | Histiocytic sarcoma |
| C969 | Haematological malignancies | Malignant neoplasm of lymphoid, haematopoietic and related tissue, unspecified |

#### Pathogenic and Contaminant Organisms

Blood culture isolates classified as pathogenic or contaminant, as used to derive the any_bc_pos outcome and the organism lists exported for external validation.

##### Pathogens

**Organism**

Achromobacter species

Achromobacter xylosoxidans

Acinetobacter baumannii complex

Acinetobacter johnsonii

Acinetobacter lwoffii

Acinetobacter nosocomialis

Acinetobacter pittii

Acinetobacter species

Acinetobacter ursingii

Actinomyces neuii

Actinomyces odontolyticus

Actinomyces oris

Actinomyces species

Aerococcus urinae

Aeromonas caviae

Aeromonas veronii

Arcanobacterium haemolyticum

Bacillus cereus

Bacteroides caccae

Bacteroides fragilis

Bacteroides ovatus

Bacteroides sp.

Bacteroides thetaiotamicron

Brucella abortus

Brucella melitensis

Brucella species

Burkholderia cepacia

Burkholderia multivorans

Campylobacter jejuni

Candida albicans

Candida glabrata

Candida parapsilosis

Candida tropicalis

Capnocytophaga sputigena

Cardiobacterium hominis

Citrobacter freundii

Citrobacter koseri

Clostridium paraputrificum

Clostridium perfringens

Clostridium septicum

Clostridium species

Clostridium sporogenes

Enterobacter asburiae

Enterobacter cloacae

Enterobacter cloacae complex

Enterobacter sp.

Enterococcus avium

Enterococcus casseliflavus

Enterococcus durans

Enterococcus faecalis

Enterococcus faecium

Enterococcus gallinarum

Enterococcus raffinosus

Escherichia coli

Fusobacterium necrophorum

Fusobacterium nucleatum

Fusobacterium species

Haemophilus influenzae

Kingella kingae

Klebsiella aerogenes

Klebsiella oxytoca

Klebsiella pneumoniae

Klebsiella species

Klebsiella variicola

Listeria monocytogenes

Methicillin Resistant Staph. aureus (MRSA)

Morganella morganii

Mycobacterium chelonae

Mycobacterium fortuitum

Mycobacterium species

Neisseria meningitidis

Pantoea species

Pasteurella multocida

Prevotella bivia

Prevotella buccae

Prevotella intermedia

Prevotella melaninogenica

Prevotella species

Proteus mirabilis

Proteus penneri

Proteus vulgaris

Providencia rettgeri

Pseudomonas aeruginosa

Pseudomonas oryzihabitans

Pseudomonas putida

Pseudomonas species

Pseudomonas stutzeri

Salmonella Enteritidis

Salmonella Javiana

Salmonella Newport

Salmonella Paratyphi A

Salmonella Paratyphi B

Salmonella Typhi

Salmonella Typhimurium

Salmonella species

Serratia marcescens

Serratia species

Staphylococcus aureus

Staphylococcus lugdunensis

Stenotrophomonas maltophilia

Streptococcus Group G

Streptococcus agalactiae (Group B)

Streptococcus anginosus

Streptococcus constellatus

Streptococcus dysgalactiae

Streptococcus dysgalactiae (Group C/G)

Streptococcus gallolyticus

Streptococcus intermedius

Streptococcus pneumoniae

Streptococcus pyogenes (Group A)

##### Contaminants

**Organism**

Abiotrophia defectiva

Actinobacillus species

Actinotignum sanguinis

Aerococcus viridans

Alcaligenes faecalis

Anaerococcus octavius

Atopobium species

Bacillus licheniformis

Bacillus sp.

Bifidobacterium species

Bilophilia wadsworthia

Brevibacillus parabrevis

Brevibacterium casei

Brevibacterium species

Brevundimonas vesicularis

Cellulosimicrobium cellulans

Chryseobacterium species

Coagulase Negative Staphylococcus

Corynebacterium amycolatum

Corynebacterium jeikeium

Corynebacterium mucifaciens

Corynebacterium pseudodiphtheritcum

Corynebacterium species

Corynebacterium striatum

Cutibacterium acnes

Cutibacterium species

Dermabacter species

Dialister micraerophilus

Dialister pneumosintes ( prev Bacteroides pneumo.)

Eggerthella lenta

Escherichia vulneris

Eubacterium species

Facklamia hominis

Finegoldia magna

Gemella haemolysans

Gemella morbillorum

Gemella sanguinis

Gemella species

Gordonia species

Granulicatella adiacens

Haemophilus parainfluenzae

Kocuria kristinae

Kocuria rhizophila

Kocuria species

Lactobacillus gasseri

Lactobacillus paracasei

Lactobacillus species

Leptotrichia species

Leuconostoc species

Microbacterium paraoxydans

Microbacterium species

Micrococcus luteus

Micrococcus species

Moraxella osloensis

Moraxella species

Neisseria species

Ochrobactrum anthropi

Oligella urethralis

Paenibacillus sp

Parabacteroides distasonis

Paracoccus yeei

Parvimonas micra

Peptoniphilus harei

Peptoniphilus species

Propionibacterium species

Psychrobacter sanguinis

Raoultella ornithinolytica

Raoultella species

Rhizobium radiobacter

Roseomonas species

Rothia dentocariosa

Ruminococcus gnavus

Slakia exigua

Staphylococcus capitis

Staphylococcus caprae

Staphylococcus cohnii

Staphylococcus epidermidis

Staphylococcus haemolyticus

Staphylococcus hominis

Staphylococcus pasteuri

Staphylococcus petrasii

Staphylococcus pettenkoferi

Staphylococcus pseudintermedius

Staphylococcus saccharolyticus

Staphylococcus saprophyticus

Staphylococcus simulans

Staphylococcus species

Staphylococcus warneri

Streptococcus canis

Streptococcus gordonii

Streptococcus infantarius

Streptococcus lutetiensis

Streptococcus mitis

Streptococcus mutans

Streptococcus oralis

Streptococcus oralis/mitis

Streptococcus parasanguinis

Streptococcus salivarius

Streptococcus sanguinis

Streptococcus species

Streptococcus vestibularis

Streptococcus zooepidemicus

Terrisporobacter glycolicus

Trueperella bernardiae

Veillonella parvula

Veillonella species
